# Multimodal surveillance of SARS-CoV-2 at a university enables development of a robust outbreak response framework

**DOI:** 10.1101/2022.07.06.22277314

**Authors:** Brittany A. Petros, Jillian S. Paull, Christopher H. Tomkins-Tinch, Bryn C. Loftness, Katherine C. DeRuff, Parvathy Nair, Gabrielle L. Gionet, Aaron Benz, Taylor Brock-Fisher, Michael Hughes, Leonid Yurkovetskiy, Shandukani Mulaudzi, Emma Leenerman, Thomas Nyalile, Gage K. Moreno, Ivan Specht, Kian Sani, Gordon Adams, Simone V. Babet, Emily Baron, Jesse T. Blank, Chloe Boehm, Yolanda Botti-Lodovico, Jeremy Brown, Adam R. Buisker, Timothy Burcham, Lily Chylek, Paul Cronan, Valentine Desreumaux, Megan Doss, Belinda Flynn, Adrianne Gladden-Young, Olivia Glennon, Hunter D. Harmon, Thomas V. Hook, Anton Kary, Clay King, Christine Loreth, Libby Marrs, Kyle J. McQuade, Thorsen T. Milton, Jada M. Mulford, Kyle Oba, Leah Pearlman, Mark Schifferli, Madelyn J. Schmidt, Grace M. Tandus, Andy Tyler, Megan E. Vodzak, Kelly Krohn Bevill, Andres Colubri, Bronwyn L. MacInnis, A. Zeynep Ozsoy, Eric Parrie, Kari Sholtes, Katherine J. Siddle, Ben Fry, Jeremy Luban, Daniel J. Park, John Marshall, Amy Bronson, Stephen F. Schaffner, Pardis C. Sabeti

**Author notes:** These authors contributed equally (Brittany A. Petros, Jillian S. Paull, Christopher H. Tomkins-Tinch, Bryn C. Loftness). corresponding authors (lead contact); (wastewater); (wifi). These authors jointly supervised this work (Ben Fry, Jeremy Luban, Daniel J. Park, John Marshall, Amy Bronson, Stephen F. Schaffner, Pardis C. Sabeti).

## Abstract

Universities are particularly vulnerable to infectious disease outbreaks and are also ideal environments to study transmission dynamics and evaluate mitigation and surveillance measures when outbreaks occur. Here, we introduce a SARS-CoV-2 surveillance and response framework based on high-resolution, multimodal data collected during the 2020-2021 academic year at Colorado Mesa University. We analyzed epidemiological and sociobehavioral data (demographics, contact tracing, and wifi-based co-location data) alongside pathogen surveillance data (wastewater, random, and reflexive diagnostic testing; and viral genomic sequencing of wastewater and clinical specimens) to characterize outbreak dynamics and inform policy decisions. We quantified group attributes that increased disease risk, and highlighted parallels between traditional and wifi-based contact tracing. We additionally used clinical and environmental viral sequencing to identify cryptic transmission, cluster overdispersion, and novel lineages or mutations. Ultimately, we used distinct data types to identify information that may help shape institutional policy and to develop a model of pathogen surveillance suitable for the future of outbreak preparedness.

## Intro

Infectious disease outbreaks are existential threats to congregate communities; in universities, in particular, students are susceptible because of close-quarters housing^1, 2^, dense social networks^3–5^, and widespread involvement in sports teams and other student organizations^5, 6^. Students may also be individually vulnerable to infection due to sleep deprivation^7^ and poor hygiene^8^. In addition to their own susceptibility, universities have potential to drive transmission in surrounding communities^9–11^.

At the same time, residential universities are ideal environments for the study of pathogen transmission and the impact of interventions and surveillance thanks to their semi-insular character and their role as centers of innovation^12^. In response to SARS-CoV-2 they have widely employed high-cadence testing^13–15^, vaccination programs^16, 17^, strict quarantine of cases in dedicated facilities^18–21^, and social distancing measures^22–25^. In addition, universities are well-positioned to test and implement new surveillance methods that can subsequently be applied at greater scale. For example, they were among the first to implement wastewater surveillance for SARS-CoV-2^18, 26^, institution-wide viral sequencing^21, 27^, and contact tracing via wifi network co-location data^28, 29^.

In Fall 2020, Colorado Mesa University (CMU) committed to in-person instruction of ∼8,000 students for the 2020–2021 academic year, motivated by a desire to avoid amplifying resource disparities via remote learning. This decision necessitated a rigorous SARS-CoV-2 surveillance program, balancing public health goals with efficient use of limited resources. Given these considerations, CMU eschewed mandatory periodic testing of all university community members in favor of a surveillance program with randomized testing and robust reflexive testing – *i.e.*, strategic testing of students due to symptoms (reported through the *Scout* web-based tool^30^), contact with recently diagnosed individuals, or a positive wastewater signal in their residential dorm.

CMU piloted *Lookout*, a tool integrating multiple data types to identify, alert, and test individuals or groups at increased risk of infection (Figure 1; demo: https://sentinel.network/lookout-demo-campus). Lookout integrated numerous data types, including clinical diagnostic test results, student attributes such as residence hall and sports team affiliations, self-reported contacts of positive individuals, viral genome sequences from diagnostic specimens, and wastewater viral titers. The interactive dashboard allowed the administration to quickly identify students at risk of infection and to minimize opportunities for transmission. Here, we explore the utility of combining these and additional data types (including wifi co-location logs and genome sequences from wastewater effluent) to design effective disease surveillance systems.

**Figure 1.**
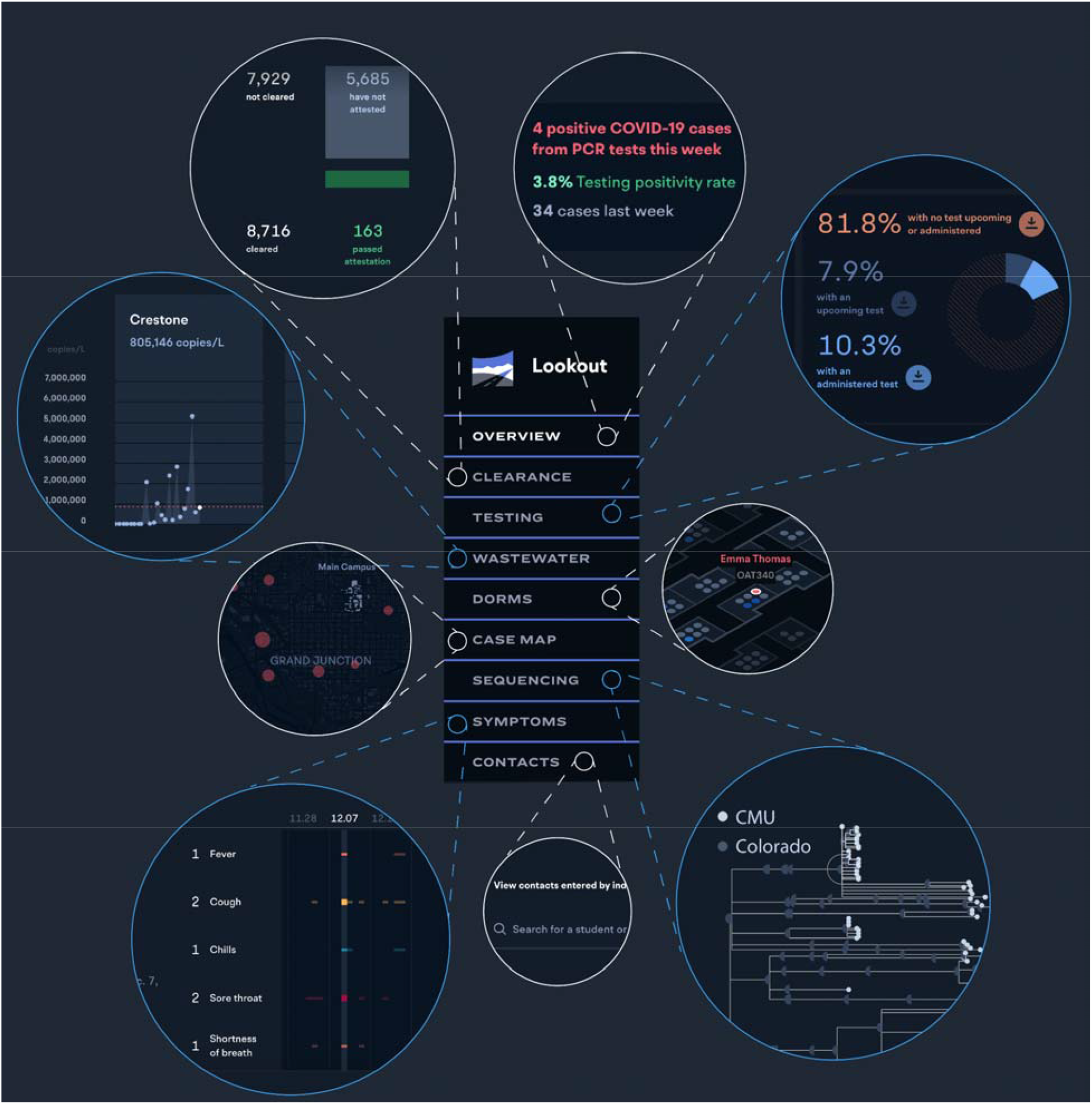
The *Lookout* system implements real-time monitoring of COVID-19 cases, including spatial, longitudinal, and social patterns of disease burden The *Lookout* tool integrated clinical diagnostic test results, student metadata, viral genome sequences from diagnostic specimens, and wastewater viral titers. A demo with representative synthetic data is available at https://sentinel.network/lookout-demo-campus/. **Overview**. Current data on community case burden, test volume, high-incidence groups, and symptom and exposure attestation. **Clearance**. Full-population counts of the individuals complying with the training and symptom attestation requirements for campus entry. **Testing**. Reports of positive tests in the past 7 days as well as the volume of tests scheduled, taken, and missed for the current week. **Testing - Baseline**. A view of “back to campus” testing measures tracking the number of tests administered over time against the amount needed to successfully test the entire population before a return to campus. **Wastewater**. Viral load over time, measured on a per-sewershed basis, aligned with individual test results from the same residence halls. **Dorms**. Views showing spatial position of residence hall cases on a per-floor basis. Individuals may be selected to view their associated membership groups (*i.e.*, potential close contacts), present attestation, test, and isolation status. Main view page depicts campus-wide case density. **Case Map**. Large-area view of case locations for members of the university community who live off-campus, with hot spots for locations of high case density. **Sequencing**. Phylogenetic tree showing the ancestry and clustering of viral genomes collected from university cases. Individual cases may be selected to view other individuals who are members of the same cluster, as well as specific attributes of the individual. Viral lineages are noted. **Symptoms**. Timelines depict reported symptoms for students or staff, including fever, cough, chills, sore throat, shortness of breath, loss of smell/taste, and runny nose. **Contacts**. Rapid search for the list of contacts reported by cases, and their associated contact information. **Lookup**. Review of information in *Lookout* for a given individual, including group affiliations, test result history, symptom history, attestation history, and contact history.

## Results

### 1. CMU deployed a comprehensive and effective surveillance program based on a multi-pronged testing approach

Over the 2020–2021 academic year, CMU’s surveillance program identified 1,113 COVID-19 cases (1,076 students, 37 faculty or staff) through randomized and targeted (*i.e.*, symptomatic or reflexive) testing. The test positivity rate was 5.1% in Fall 2020 (August 17–November 20) and 1.5% in Spring 2021 (January 18–April 30) (Figure 2A–C); individuals who tested positive were moved to an isolation dorm. CMU’s randomized testing strategy sampled students non-uniformly to test more frequently those at greater risk of onward transmission, *i.e.*, on-campus students and athletes. Consistent with this bias, on-campus students and athletes ultimately tested positive 1.30 and 2.45 times as frequently as expected given uniform sampling (Appendix Figure 1C).

**Figure 2.**
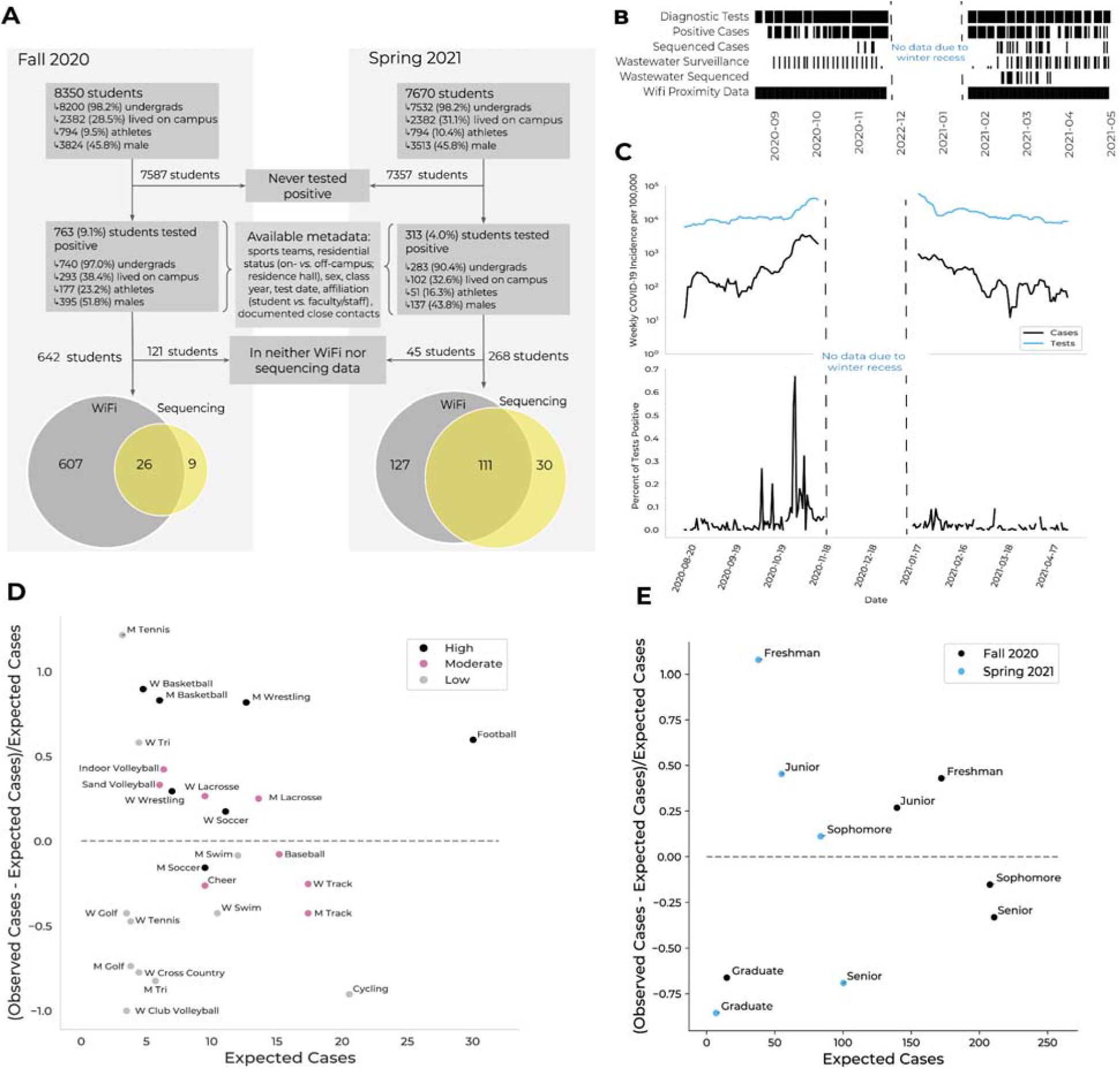
Data types, incidence rates, and epidemiological risk factors for SARS-CoV-2 positivity on Colorado Mesa University’s campus **A.** Cohort description. A subset of students at Colorado Mesa University (CMU) tested positive for COVID-19 *via* reflexive or random surveillance RT-qPCR testing. CMU provided demographic and behavioral metadata for each case. The majority of students who tested positive were enrolled in the wifi proximity program (gray). A subset of positive samples were available for viral genomic sequencing (yellow). **B.** Timeline showing data collection timepoints (black) by data type during the Fall and Spring semesters. Data not shown for November 21– January 18 due to winter recess. **C.** Upper: Weekly COVID-19 incidence (black) and number of tests conducted (blue) over the 2020–2021 academic year. Lower: Percent positivity rate. Data not shown for November 21–January 18 due to winter recess. **D.** The difference between the number of cases observed and the number of cases expected (based on sports team player count and scaled by the number of cases expected; *y* axis) *vs.* the number of cases expected (*x* axis), per sports team. The dashed line at y=0 separates teams with more (above) or fewer (below) cases observed than expected. Teams are colored by contact level (legend). M refers to men’s teams and W to women’s teams. **E.** The difference between the number of cases observed and the number of cases expected (based on class size and scaled by the number of cases expected; *y* axis) *vs.* the number of cases expected (*x* axis), per class year. The dashed line at *y* = 0 separates classes with more (above) or fewer (below) cases observed than expected. Classes are colored by semester (legend).

In addition, CMU’s reflexive testing program tested individuals identified by institutional contact tracing as close contacts. Of the identified positive individuals, 720 (65%) reported close contacts, enabling subsequent detection of 93 distinct cases (8.4% of the total cases) within a week of the sentinel case’s positive test. These efforts identified plausible transmission links; among pairs of sequenced cases identified via contact tracing, 79% had closely related genomes (with a genetic distance of at most two mutations), compared to 10% among sequenced pairs more generally (Appendix Figure 2A).

Frequent wastewater surveillance enhanced reflexive testing efforts, identifying SARS-CoV-2-positive residence halls whose residents were then randomly selected for follow-up testing. The effort captured effluent from ∼75% (Fall) and ∼85% (Spring) of the residential student population. In response to spikes in viral titer, contributing residence halls were oversampled for testing; when warranted, up to half of the residents of a hall were tested. The success of this program is reflected in the close correlation of wastewater viral titer with contemporaneous case (Spearman ρ=0.40, p<.001; Figure 5AB, Appendix Figure 3).

**Figure 3.**
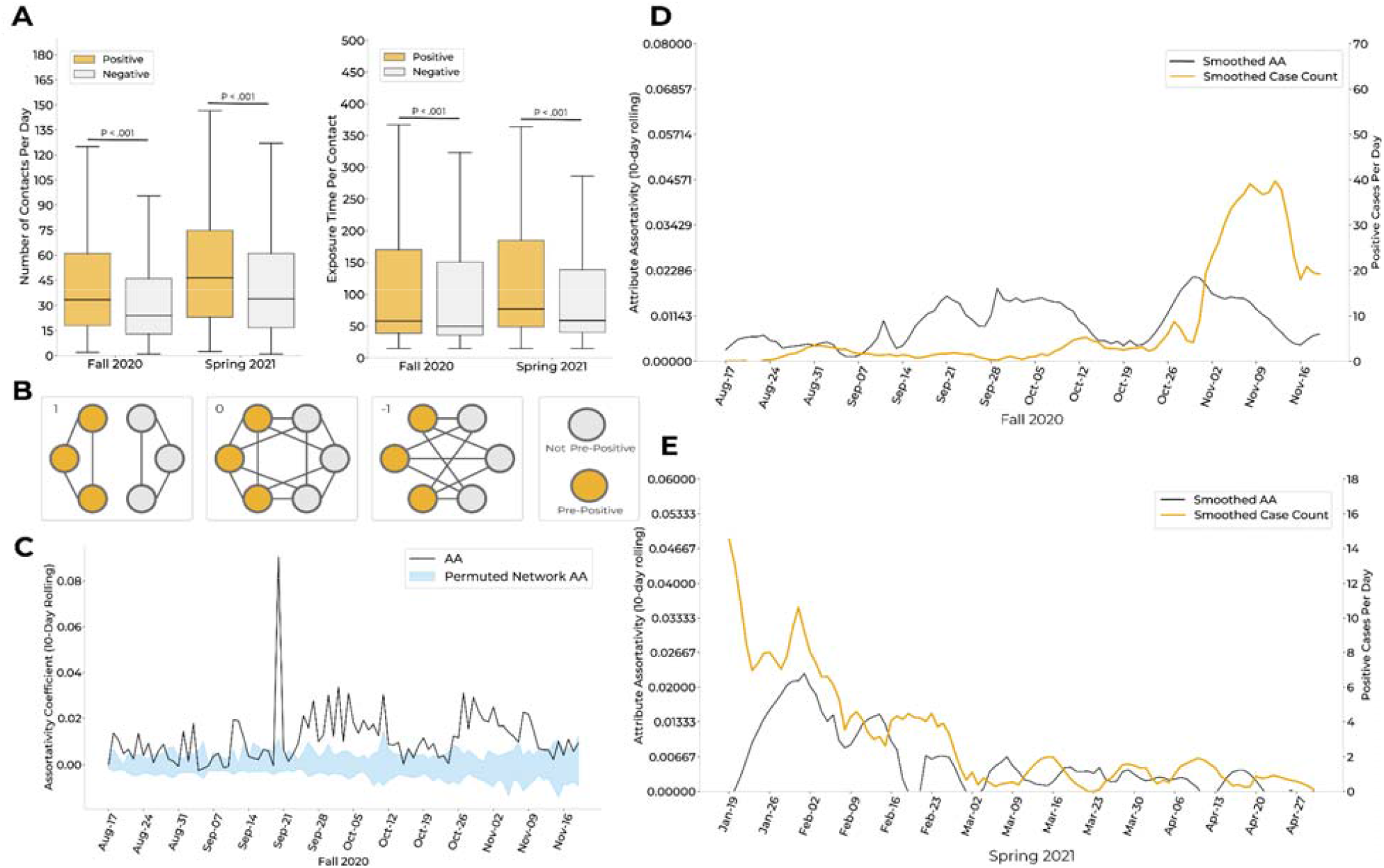
Social connectivity network from the wifi co-location data identifies behavioral trends that correlate with case counts **A.** Left: Distributions of daily contacts (Appendix Table 11) for students who tested positive over the course of each semester (orange) *vs.* those who remained negative (gray) over the course of each semester. Effect sizes, 9.5 (Fall; positive median = 33.5, negative median = 24), 12.5 (Spring; positive median = 46.5, negative median = 34). Right: Distributions of average exposure time per contact (Appendix Table 11), in minutes, for cases (orange) *vs.* those who remained negative (gray). Effect sizes, 8.2 (Fall; positive median = 58, negative median = 49.8), 11.8 (Spring; positive median = 77, negative median = 58.8). **B**. Visual representation of the network metric attribute assortativity (AA; described in Supplemental Methods). We depict network scenarios where the AA coefficient is equal to 1, 0, or −1. When AA = 1, pre-positive individuals, defined as those within 10 days prior to their positive test, have interactions onl with pre-positive individuals, and vice versa. When AA = 0: pre-positives individuals have an equal likelihood of interactions with pre-positive individuals and non-pre-positive individuals. When AA = −1, pre-positive individuals have interactions only with non-pre-positive individuals, and vice versa. Positive AA values indicate a higher propensity for within-group interactions, while negative values indicate a higher propensity for between-group interactions. **C**. Comparison of per-semester AA for individuals within a 10 day window of a positive test (*i.e.*, pre-positives) *vs.* those who never tested positive (*i.e.*, negatives). 95% confidence intervals (CI; blue) were calculated by permuting pre-positive and negative labels for individuals within the proximity network each day, 40 times. The AA of the proximity network (black) was above the upper bound of the CI for 69.4% (66/95 days) of the Fall 2020 semester, implying significance at p < 0.025. Only Fall 2020 is presented here; methodology and results were consistent for Spring 2021 (Appendix Figure 8). **D-E**. Comparison of smoothed case counts and smoothed AA for Fall 2020 (**D**) and Spring 2021 (**E**). Smoothing *via* the Savitzky-Golay filter (window length = 17, polynomial order = 4).

To assess the overall efficacy of CMU’s surveillance program, we compared CMU’s incidence rate to that of Mesa County, which had limited testing available at the time. CMU’s weekly incidence exceeded county incidence rates and predicted them with a lag time of 3 days (correlation = 0.73; Appendix Figure 1AB). This is consistent with reports that adequate university testing can foreshadow community outcomes^12^ and highlights the ability of well-designed university testing programs to serve as bellwethers. As the pandemic’s impact on the surrounding community became clearer, the university sponsored testing for external community members, both as a public benefit and to limit spread of SARS-CoV-2 into the campus^31^.

### 2. Epidemiological analyses identified student attributes associated with SARS-CoV-2 positivity and support a surveillance paradigm of targeted testing and risk mitigation

Here we evaluate risk factors among a wide range of institutionally captured attributes for individuals who tested positive: role (*i.e.*, student or faculty/staff), sex, class year^32^, test date, association with a residence hall, and membership in a sports team. Residence halls (Appendix Table 1) and sports teams (Appendix Table 2) were annotated with features, including perceived contact risk for sports teams (defined in Appendix Table 3). Our results support a two-pronged surveillance strategy, in which groups at increased risk are targeted for higher-cadence testing, while putatively causal factors are mitigated via institutional policies that reduce risk.

Besides athletes and on-campus students, whose higher risk of testing positive could reflect increased sampling, males, freshmen, and juniors also exhibited more cases than expected by chance (Appendix Figure 1C; Figure 2E; Appendix Table 4). These findings may underscore risk factors relevant for other universities: for example, a potential commonality between freshmen and juniors is that many students moved to new living arrangements (on-campus for freshmen, off-campus for juniors). Future surveillance efforts could target populations undergoing such transitions, though real-time epidemiology is essential to identify community-specific risk factors.

Looking more closely among sports teams, we identified specific attributes that further predict disease risk. High-contact sports teams had increased disease risk (Appendix Table 4), with 50% more cases than expected from the risk for athletes as a whole, while low-contact teams had 47% fewer. Interestingly, risk was not uniform across teams of comparable contact level; women’s basketball had 90% more cases than expected, while men’s soccer had 16% fewer, despite both being high-contact sports (Figure 2D). We found no association between sports location (*i.e.*, indoor *vs.* outdoor sports) and case counts, though sports played in both seasons had higher case incidence than fall or spring sports (Appendix Table 4). These findings are consistent with a model where individual athletes sporadically contract COVID-19, with an increased risk of further transmission and thus outbreaks on higher-contact teams or teams with longer seasons.

Because COVID-19 incidence rates varied approximately three-fold from 9.7% to 27% across residence halls (Appendix Table 1), we conducted linear regression with multiple possible predictors related to halls to characterize factors that influenced case rates (Appendix Figure 4AB). Two features were significant predictors: percent occupancy (*i.e.*, number of available beds filled) and private (*vs.* hallway) bathrooms (see Appendix Figure 4C-E for model validation). We included an indicator variable for fall wastewater tracking to assess whether reflexive testing biased hall incidence rates; it was dropped as a predictor and thus does not significantly bias rates.

**Figure 4.**
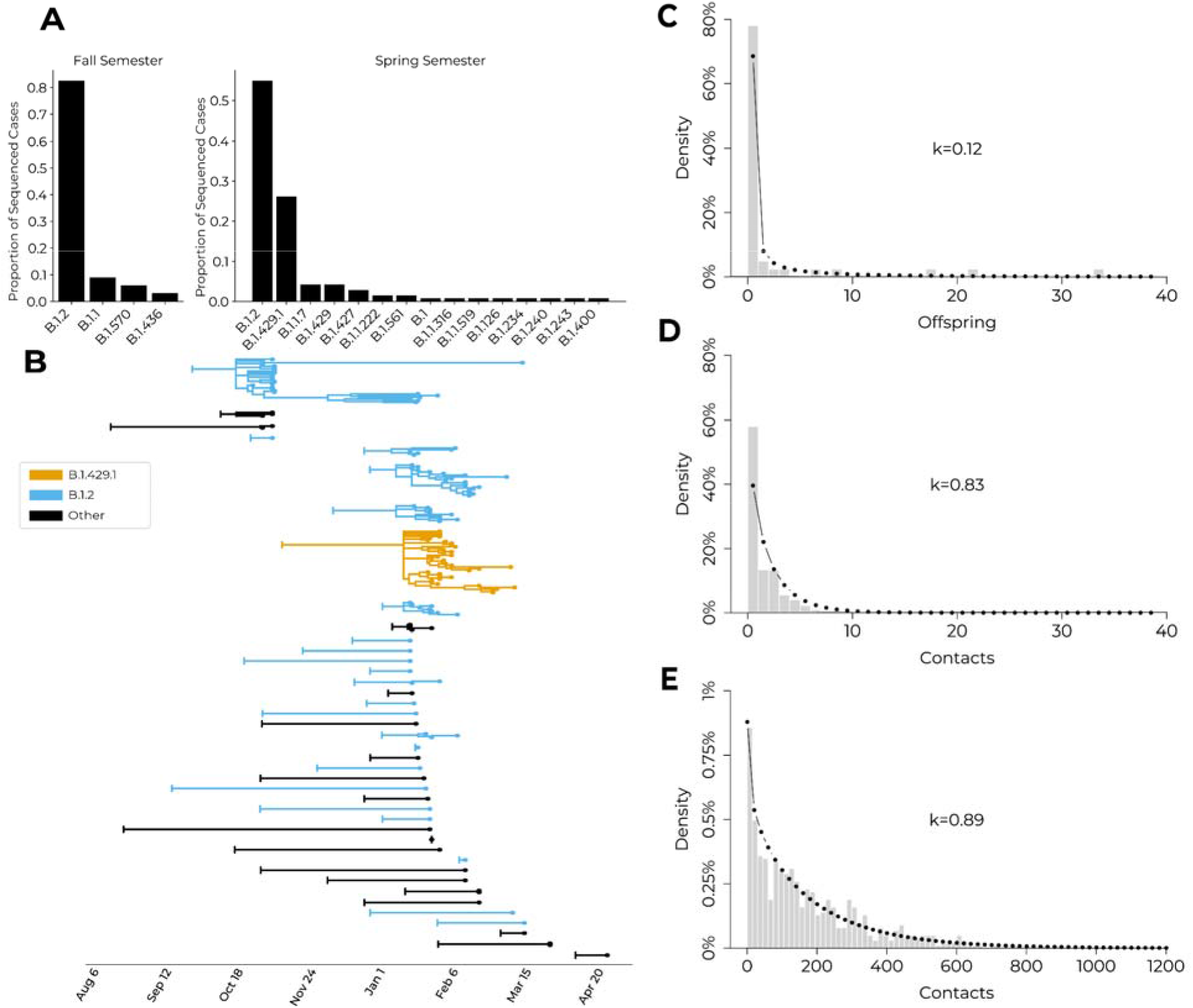
Case clusters and lineages identified by viral genomic sequencing, and comparison of genomic cases to reported social network degree distributions **A.** Pango lineage proportions for university cases during the Fall 2020 and Spring 2021 semesters. **B.** Phylogenetic trees for inferred introductions to the campus community with branch lengths scaled to time. B.1.2 clusters (blue), the B.1.429.1 cluster (orange), and all other lineages (black). Vertical bar on the left of each introduction indicates the inferred ancestral root date of each university case or case cluster; cases are tip dots at right of each tree. **C.** Distribution of offspring from phylogenetic clusters, with a negative binomial distribution fit and overlaid (dotted line) to quantify overdispersion. Unique introductions and resulting offspring were grouped into phylogenetic clusters; mean=2.56 offspring/introduction; k=0.13 (dispersion parameter). **D.** Distribution of the number of contacts from positive individuals identified during contact tracing, with a negative binomial distribution fit and overlaid (dotted line). Contacts were defined as any individuals with interaction dutations greater than 15 minutes in the 48-hour period prior to positive test or symptom onset; mean=1.71 contacts per positive individual; k=0.83. **E.** Distribution of the number of wifi contacts observed from positive individuals, with a negative binomial distribution fit and overlaid (dotted line). Contacts were defined as individuals with interaction durations greater than 15 minutes in the 48-hour period prior to testing positive or symptom onset; mean=177.94 contacts per positive individual; k=0.89.

For every increase of 10% in occupancy, our model predicted an increase of 0.015 in incidence, supporting institutional de-densification measures to limit the transmission risks associated with denser populations. Strikingly, halls with en-suite or private bathrooms were predicted to have an incidence 0.059 higher than those with hallway bathrooms (Appendix Figure 4B), consistent with reports that a majority of SARS-CoV-2 transmissions occur within households (here, within suites)^33^. Possible explanations include compensatory protective measures (*i.e.*, masking or social distancing) present in larger bathrooms and the increased hygiene of hallway bathrooms, which were cleaned by professional staff rather than residents. Importantly, our model does not account for possible social confounders such as clustering of certain groups (*e.g.*, athletes or freshmen) within specific residence halls.

### 3. Distinct interaction dynamics of positive individuals within wifi proximity dataset reveal potential for incorporation into disease surveillance

Here, we explore a dataset of anonymized daily logged connection locations (*i.e.*, access point and building) for students connected to campus wifi for at least 15 minutes, and describe how such data can be extended for real-time disease surveillance. Data were obtained from an existing program implemented in 2018 to assess facility use and student engagement, in order to aid the university in allocation of resources. All students were alerted via a campus-wide notice about the program, and could choose to opt out; an overwhelming majority (98%) participated.

Through an examination of campus-wide connectivity patterns, we identified associations between student activity and CMU’s COVID-19-related policies. We found elevated on-campus presence during weekdays (*vs.* weekends) and in residence halls (*vs.* other building types) in fall 2020 (Appendix Figures 5, 6A, 6C), reflecting university policies that discouraged on-campus gathering. When mitigation policies relaxed in Spring 2021, weekend presence increased relative to fall 2020 (Appendix Figure 6B and 6D). Additionally, after testing positive, individuals had 42% fewer contacts than during the preceding 10 days, indicating adherence to isolation policies (Appendix Figure 7A). This quantification of policy adherence suggests that wifi datasets can be used to assess policy implementation or to determine the effects of policy updates in real time.

**Figure 5.**
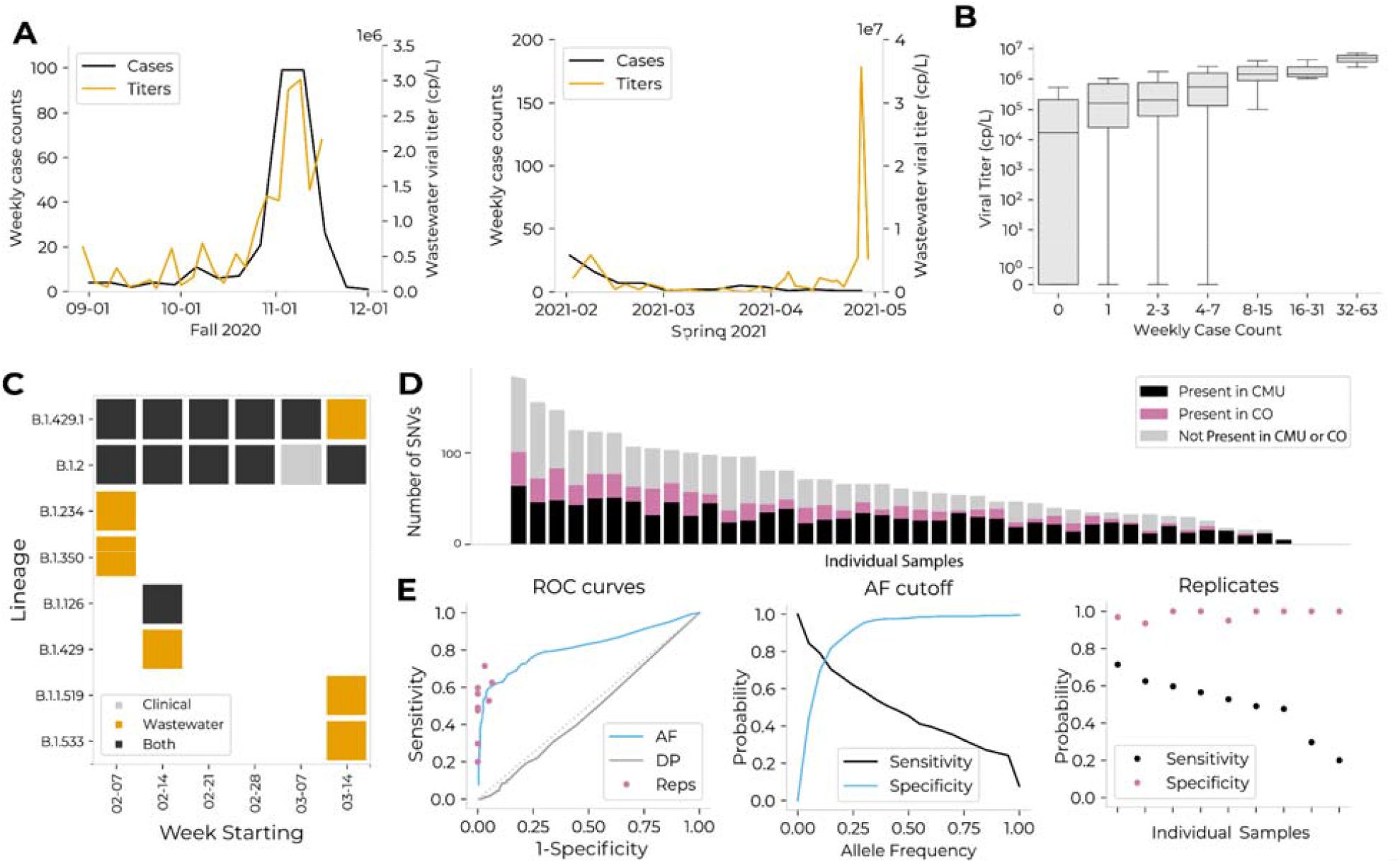
Wastewater surveillance and sequencing for measurement of aggregate viral load, identification of circulating lineages, and comparison with viral genomes from contemporaneous clinical cases **A.** Average wastewater viral titers (orange) *vs.* weekly residential case count (black). Residential case counts were calculated relative to the subsets of dorms monitored (75% of residential population in Fall 2020; 99% in Spring 2021). There was an anomalous peak in wastewater viral titer observed in April, which could be due to technical error, differential shedding patterns, or undiscovered positive individuals. **B.** Viral titer (*y* axis) *vs.* binned weekly case count (*x* axis; binned by powers of 2) for each wastewater sample. Viral titer and case count were significantly associated *via* Fisher’s exact test (p=0.04) and Spearman’s correlation=0.40 (p<0.001). **C.** Lineages detected on campus via wastewater or clinical sequencing. **D.** The number of single nucleotide variants (SNVs) detected in wastewater samples; each bar represents a single sample. Individual samples are organized on the *x* axis in order of total number of SNVs. For each sample, SNVs are categorized by whether they were present in clinical sequences from CMU (black), in clinical sequences from Colorado (pink), or in neither (gray). On average, 51% of SNVs in a single wastewater sample were not found in CMU clinical samples, and 36% were not found in Colorado clinical samples. **E.** Comparison of qualit control methods to remove SNVs not validated by presence in Colorado clinical sequences. The three methods compared are: 1) allele frequency (AF), or discarding SNVs present at an allele frequency below a given threshold; 2) read depth (DP), or discarding SNVs located at a site with a read depth below a given threshold; and 3) replicates (Reps), or discarding SNVs not present within both of two technical replicates of a given sample. These analyses are limited to the nine samples for which technical replicates exist. Left: ROC curves for each of the three filters. Middle: Sensitivity and specificity for allele frequency-based quality control method. Right: Sensitivity and specificity for replicate-based quality control method.

**Figure 6.**
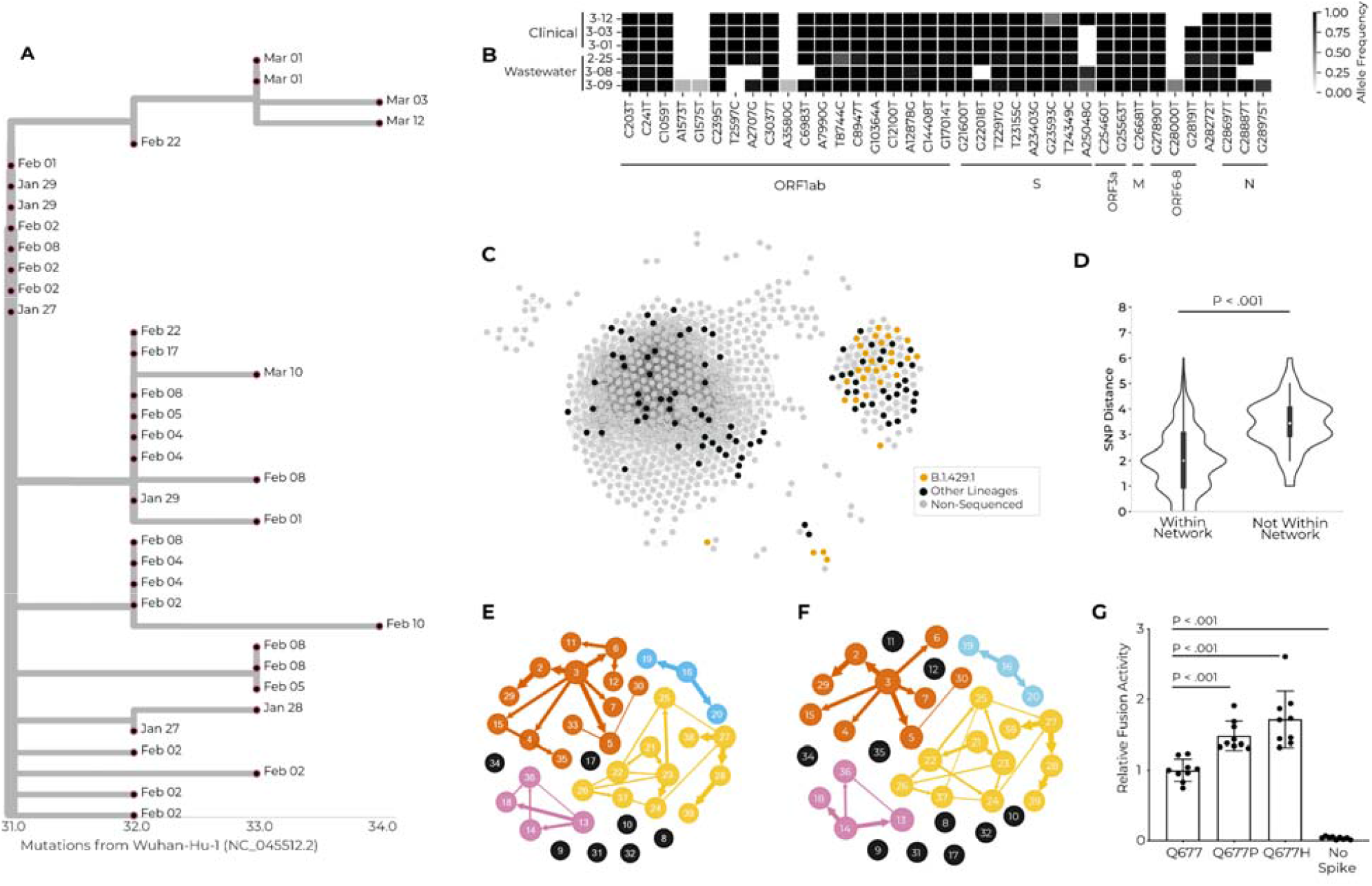
A multimodal exploration of the novel lineage B.1.429.1 *via* clinical and environmental genomic sequencing, wifi proximity analyses, transmission reconstruction networks, and experimental validation **A.** Phylogenetic tree showing the relationship between cases within the B.1.429.1 case cluster. Tree tips are anchored at their dates of sample collection, and branch lengths are scaled by maximum likelihood. One-to-many polytomies depict clonal amplification, with multiple cases with the same ancestral node rather than as a series of bifurcating branches. **B.** Three wastewater samples and three clinical samples (*y* axis), all of the B.1.429.1 lineage. The three wastewater samples had B.1.429.1 as the sole identified lineage, and were extracted from Site 3, for which residential halls B and M are the only upstream contributors. Shown in comparison are clinical viral genomes from three students believed to have contributed effluent to these wastewater samples, based on residential status and test date. The *x* axis represents all single-nucleotide variants present in at least one wastewater sample with 25% allele frequency or greater. SNVs are grouped by genomic position. **C.** Social proximity network for interactions occurring between two individuals who are both within a 10-day window prior to testing positive. Edges in this network represent one or more simultaneous wifi access point connections. Each node represents a positive individual. The node color represents their sequencing status (legend). **D.** Comparison of genetic distance for individuals of the B.1.429.1 lineage who are or are not connected *via* the social proximity network shown in (**C**). Effect size = 1 mutation. **E-F.** Transmission reconstruction network for B.1.429.1 cases created with genomic information as well as manual contact tracing data (**E**) or wifi-inferred 10-day contact data (**F**). **G.** Results of cell-cell fusion activity of viral pseudotypes with the ancestral allele, or with the S:Q677P or S:Q677H amino acid changes, relative to a luminescent control with no Spike protein expressed.

We found that positive individuals exhibited distinct patterns in their social networks and behaviors. Individuals who eventually tested positive exhibited larger social networks than those who remained negative: they spent more days on campus (Appendix Figure 7B), had more daily contacts (Figure 3A, left), and interacted for a longer duration with each contact (Figure 3A, right), creating more opportunities for potential viral transmission. Furthermore, in the 10 days preceding both of their positive tests, pairs of students identified *via* contact tracing had significantly longer daily interaction times than other pairs of positive students (*e.g.*, median of 104 *vs.* 45 minutes per interaction per day in the fall; Appendix Figure 7CD). Moreover, these pairs of positive students between whom COVID-19 transmission may have occurred interacted for significantly longer than for pairs in which transmission did not or could not have occurred (*i.e.*, pairs involving one or more students who never tested positive; Appendix Figure 7CD). These distinct qualities support supplementing, and if necessary substituting, manual contact tracing with a wifi-based system to automatically flag close contacts of positive individuals.

We further explored interactions between positive and negative individuals using the attribute assortativity metric (AA), which quantifies the extent to which individuals interact within-group *vs.* between-group compared to random assortment (Figure 3B). We found that both positive (*i.e.*, individuals positive at any point over the semester) and pre-positive individuals (*i.e.*, individuals within 10 days of a positive test) were more likely to associate with one another than with negative individuals (Figure 3C, Appendix Figure 8A–C). Importantly, this relationship remained significant, albeit with a lower effect size, when removing pre-positive individuals who identified one another as close contacts, suggesting that this finding is not limited to positive individuals identified via reflexive testing secondary to manual contact tracing (Appendix Figure 9). Interestingly, the AA for pre-positive individuals was a leading indicator of daily case counts, by 8 days (Fall) and 3 days (Spring; Figure 3DE, Appendix Figure 8D–F), suggesting that the degree of within-group interactions among putatively infectious individuals increases in the days leading to these individuals’ positive tests.

### 4. Phylogenetic analysis of clinical viral genomes identified cluster size overdispersion and cryptic transmissions, leading to concrete policy decisions

Viral genomic sequencing from residual biomaterial enables exploration of transmission dynamics and rapid detection and monitoring of SARS-CoV-2 variants. At CMU, sequencing facilitated detection of 18 distinct Pango lineages (Figure 4A; Appendix Figure 2BC)^34^. Of these, B.1.2 was the most abundant both at CMU and in Colorado, reflecting circulation between CMU and the surrounding community and highlighting the importance of CMU’s sponsored testing for Mesa County^35^. We identified continuous transmission of this lineage between semesters, with 7 spring cases descending from 17 fall cases as the likely result of 2–3 cryptic intermediate transmissions during winter recess^36^. This cluster was non-significantly enriched for off-campus students relative to the remaining sequenced cases (83% *vs.* 70% off-campus; p = 0.15); possible off-campus continuation of the transmission chain over the break suggests that institutional surveillance programs would be wise to maintain testing availability for nearby students during school breaks.

We detected overdispersion in both genomic and social clustering data, highlighting the importance of policies that minimize super-spreading events. Of 41 detected introductions to the university, onward transmission was only observed from 13, with 5 of the resulting clusters containing 80% of sequenced cases (k=0.12 in a negative binomial model, consistent with other studies^37, 38^; Figure 4BC). We also observed overdispersion in the number of contacts between individuals in both contact tracing and wifi proximity analyses, where 80% of reported contacts were made by 33% (k=0.83) and 43% (k=0.89) of positive individuals, respectively (Figure 4DE). The similar shape of these contact distributions suggests that both datasets capture the true underlying structure of social interactions. Notably, phylogenomic cluster size demonstrates greater overdispersion than the two contact count distributions, suggesting that overdispersion in SARS-CoV-2 transmission can be partitioned into both social and biological components. Given the high variance observed amongst all three distributions, we suggest that both biological and social factors influence large SARS-CoV-2 case clusters, and that their interplay warrants further investigation. Below, we investigate the impact of a variety of variables on a distinct spring cluster.

### 5. Contemporaneous wastewater viral sequencing supplements lineage detection and enables detection of emergent mutations

During 6 weeks from February to mid-March 2021, we obtained 42 wastewater samples from 10 sites for sequencing, 9 of which were sequenced in duplicate (Appendix Figure 3C). The concurrent collection of wastewater samples and clinical specimens, with high breadth of coverage among the residential population, allowed us to directly compare viral sequences from wastewater with those from contemporaneous individual cases. Through this, we validated the utility of wastewater viral sequencing as a component of a comprehensive pathogen surveillance program, as currently instantiated by *Lookout*.

As expected, wastewater viral titers were lower than titers of clinical specimens collected from upstream individuals (Appendix Figure 10A). Wastewater and clinical samples from CMU had similar sequence coverage, suggesting that there was no particular bias in viral RNA degradation in wastewater (Appendix Figure 10B–E). This demonstrates that these wastewater samples were comparable to clinical specimens in integrity and suitable for sequencing and genomic surveillance.

We used the pre-existing Freyja tool^39, 40^ to detect 8 lineages in wastewater, 3 of which were found in concurrent clinical cases (Figure 5C, Appendix Table 6). Another 3 were observed in clinical cases prior to wastewater collection, suggesting undetected circulation on campus, shedding from previously-infected individuals, or environmental persistence. The remaining 2, B.1.533 and B.1.350, were present in the US but not the campus or state^35^; each was detected at low abundance in single samples and likely originated from a small number of individuals. Wastewater sequencing thus identified lineages not concurrently detected via clinical sequencing, proving itself to be particularly relevant in instances of incomplete clinical genomic sampling.

In addition to detection of defined lineages, wastewater sequencing can also identify novel mutations; for this latter use case, we found that quality control mechanisms were essential to identify true variation. Of 1521 wastewater single nucleotide variants (SNVs), 85% and 68% were not found at consensus-level in CMU and Colorado clinical samples, respectively, and only 4% were derived from clinical minor alleles (Figure 5D). We thus hypothesized that many mutations arose from amplification errors (*e.g.*, formation of chimeras), a theory supported by the order-of-magnitude difference in the number of SNVs detected in wastewater *vs*. clinical samples as a function of sample count (Appendix Figure 11). We subsequently developed quality control methods to corroborate mutations *via* detection in state-wide clinical genomes.

We achieved high specificity for discarding SNVs not seen in Colorado when we required presence in both of two technical replicates (specificity = 98%) or an allele frequency exceeding 25% (specificity = 92%); both methods had low sensitivity (50% and 62%, respectively; Figure 5E), as each excluded SNVs corroborated by clinical viral genomes. This analysis provides real-word evidence of the importance of replicates for identifying true SNVs in wastewater samples, a finding previously shown for clinical minor allele validation^41^.

Of 68 replicate-corroborated SNVs found across 9 wastewater samples with 2 technical replicates, 11 (16%) were not seen in clinical CMU samples (Appendix Table 7). 6 of the 11 were present in Colorado and had allele frequencies >96% in single wastewater specimens, likely reflecting on-campus circulation of viral genotypes unsampled by clinical sequencing. Of the 5 remaining mutations, 2 were non-synonymous mutations in ORF1ab (I1970S, T3462I) and were novel compared with published global variation^35^, 2 were synonymous mutations, and 1 was a premature stop codon. The latter mutation, with an allele frequency of 4%, may be spurious; the other 4, with allele frequencies between 27% and 100%, could reflect either gut tropism or cryptic transmission. Though these mutations’ phenotypic effects remain unknown, their identification serves as a proof-of-concept and provides a framework for detection of novel mutations in wastewater.

### 6. Detection of novel lineage B.1.429.1 on campus leads to high-resolution characterization of social and biological factors implicated in its spread

In Spring 2021, we detected a cluster of cases on campus that was concerning due to its unprecedented size and genomic ancestry, and proceeded to characterize it analytically and experimentally to identify the social and biological factors that contributed to its rise. This B.1.429.1 cluster initiated as a single introduction to campus, which proliferated into several star-like descendant sub-clusters, consistent with clonal amplification (Figure 6A). In total, the outbreak lasted for 45 days; in its final 4 weeks, it represented 33% of sequenced clinical samples and was the most abundant lineage in 47% of wastewater samples (Figure 6AB). B.1.429.1 descended from B.1.429 – then deemed a Variant of Concern due to reduced antibody neutralization and increased viral shedding, infectivity, and transmissibility^42^ – but also included the recurrent S:Q677H substitution, posited to further increase transmissibility^43^ (Appendix Table 8).

We did not detect differences in interaction patterns among individuals with B.1.429.1 *vs.* those with other viral lineages (Appendix Figure 12AB), suggesting that its transmissibility was due to inherent qualities of the lineage rather than the specific social dynamics of the individuals within the cluster. We did find that these individuals clustered together in social networks (Figure 6C); *i.e.*, they were on average one social connection closer to one another than to other positive individuals (p<0.001). Wifi-connected B.1.429.1 pairs also had significantly lower genetic distances between their viral genomes than non-connected B.1.429.1 pairs (Figure 6D), demonstrating that connections observed within the wifi network are enriched for genuine transmission events.

We inferred direct transmission links among B.1.429.1 cases and found that wifi connectivity data inferred transmission networks that paralleled those constructed following traditional contact tracing. Alone, manual contact tracing or genomic sequencing resolved transmission links for 61% and 68% of individuals, respectively (Appendix Figure 12DE). We thus combined genomic data with traditional contact (2 days prior to tests) or wifi-derived contract tracing (for both 2 and 10 days prior to tests), producing transmission models connecting 82%, 87%, and 74% of sequenced cases, respectively (Figure 6EF, Appendix Figure 12F). We compared the cluster topology for these networks and found that the wifi 10-day data best approximated the traditional contact tracing network (*via* Jaccard distance; Appendix Table 10). Due to the paucity of distinguishing mutations present between individual consensus mutations^44^, we used intrahost viral variation to supplement our transmission linkages. We identified a clear transmission chain where a single mutation present at low frequency in one specimen (#26 in Figure 6EF) reached fixation in two specimens (#27 & 28 in Figure 6EF) collected one week later, consistent with bottlenecked transmission. These three individuals clustered together in all reconstruction networks, but without the directionality of transmission inferred from minor alleles or phylogenetic descent, implying that transmission network reconstruction tools would benefit from further refinement.

We next analyzed inherent phenotypic factors that could explain the increased transmission of B.1.429.1 on campus. We assessed the impact of the S:Q677H mutation found in B.1.429.1 on single-cycle infectivity and on cell-to-cell fusogenicity in lentiviral pseudotypes (Appendix Figure 12GH). While the mutation did not alter virion spike protein levels or cell-free virion infectivity (Appendix Figure 12C), it significantly increased fusion efficiency relative to the ancestral B.1.429 spike protein (Figure 6G), likely due to proximity to the spike protein polybasic cleavage site. We found similar results for the S:Q677P mutation, which was detected in other contemporaneous CMU lineages. This finding is consistent with greater within-host dissemination of SARS-CoV-2 haplotypes bearing S:Q677H or S:Q677P. Fortunately, B.1.429.1 was minimally detected outside the campus, pointing to the success of CMU’s containment policies. This vignette highlights the power of systematic, multimodal surveillance programs to not only identify and mitigate transmission events, but also to contribute to novel biological characterization of viral lineages.

## Discussion

In this paper, we analyzed clinical diagnostic data, case attributes, wifi co-location logs, wastewater samples, and viral genomic sequences to quantify the success of CMU’s pandemic response program and to assess the relevance of each data type for infectious disease surveillance. Our analyses showed that through contact tracing, wastewater surveillance, and increased focus on high-risk groups, CMU effectively identified positive cases. Via novel analyses of wifi connectivity, we confirmed adherence to school policies and evaluated metrics that could allow wifi data to replace or supplement traditional contact tracing.

In addition, we leveraged phylogenetic and epidemiological analyses to propose future policies to limit disease spread (*e.g.*, continued testing during school breaks, community testing, risk prediction for testing prioritization) and to identify and mitigate specific causal factors shown to increase risk (*e.g.*, requiring high-quality masking or increased testing to participate in high-contact sports). Our sequencing of extant wastewater samples not only identified lineages independently of clinical sequencing, but also led us to apply and verify methodological procedures necessary for the detection of novel mutations from wastewater. Finally, through analysis of case cluster overdispersion and the novel lineage B.1.429.1, we highlighted the relevance of investigating both virological and sociobehavioral factors that can influence transmission.

Our results lead us to formulate a framework combining the analyzed tools within an integrated disease surveillance system (described in Figure 7). First, we emphasize beginning with mitigation policies such as symptom reporting, contact tracing, and quarantine and continuing with efficient testing strategies such as wastewater surveillance. While contact tracing is essential, it is also time-intensive and expensive to maintain; with further work to address technological and privacy concerns, wifi proximity and geolocation data could supplement and perhaps ultimately replace these efforts. Gathering epidemiological metadata, symptom attestations, and diagnostic test results digitally and with programmatic synthesis is also a high priority because it can facilitate real-time analyses and subsequent policy adjustments; the *Lookout* system serves as a useful template (Figure 1)^30^. If finances allow, we would add genomic surveillance to aid identification of transmission patterns and concerning lineages or mutations. For communities with wastewater surveillance, sequencing these samples provides a cheaper alternative to clinical sequencing of all upstream individuals and still allows for identification of lineages or mutations of interest. This tool cannot wholly replace clinical sequencing due to its inability to discern transmission trends. It is important to emphasize that disease surveillance is not a one-size-fits-all endeavor; in fact, we found parallel results across data types. We suggest that automated integration of a subset of data types will more powerfully combat infectious disease outbreaks than a siloed implementation of all data types.

**Figure 7:**
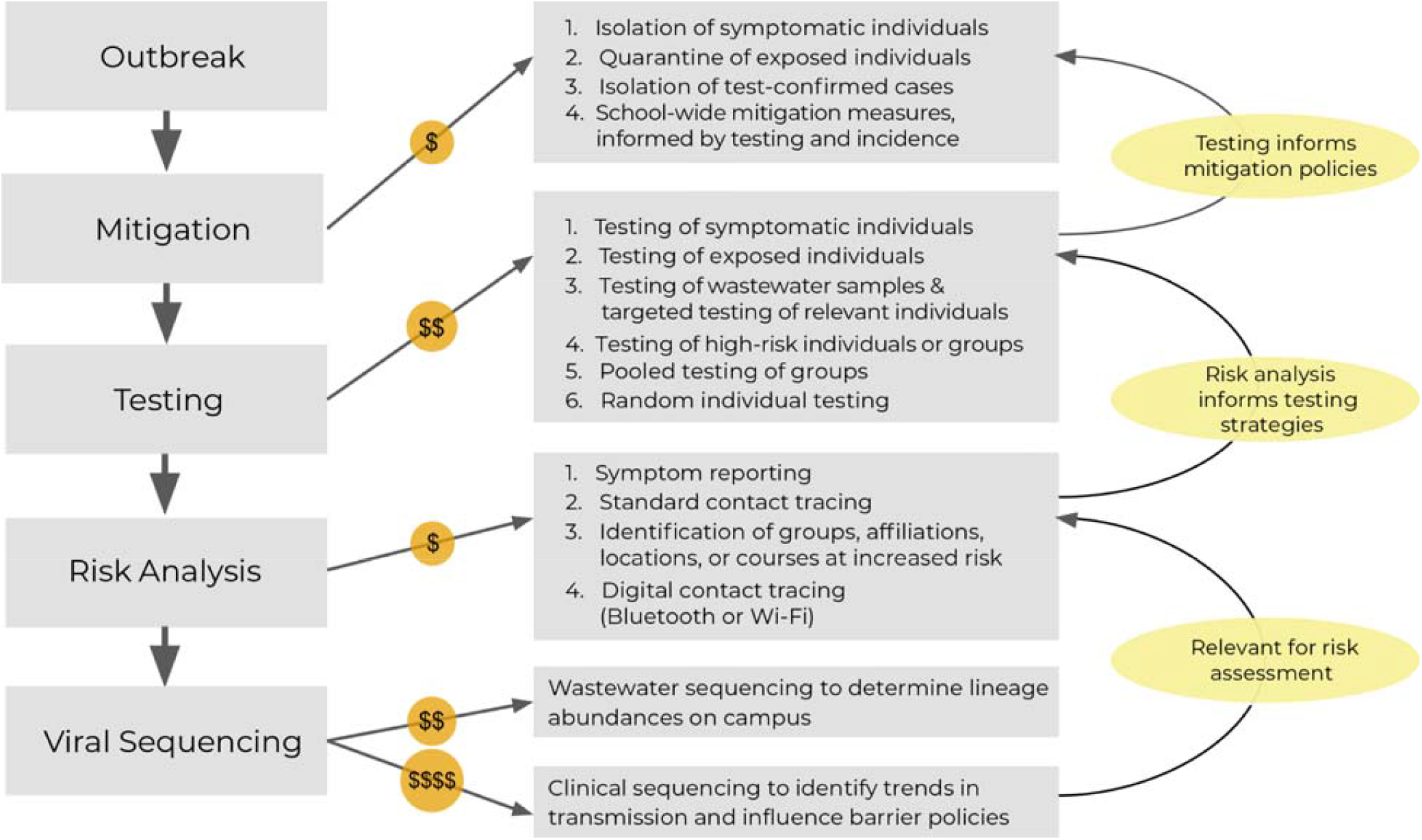
A step-wise approach to outbreak surveillance with consideration for resource limitations Illustration of the flow of actions to employ during an institutional outbreak, with delineation of relative cost and information feedback cycles. During an outbreak, initial mitigation measures can be deployed prior to and independent of a surveillance program. A basic surveillance program will first incorporate testing, the results of which will inform additional mitigation policies. Next, analyses of case attributes can be used to assess the risk of infection for specific sub-populations; these analyses will allow for development of specialized, directed testing strategies Finally, while more expensive, viral genomic sequencing of clinical or environmental samples can be used to identify transmission trends and to detect emergent viral genomic variation with potential public health or clinical relevance. This in turn can be used to inform institutional policy and the intensity of mitigation efforts. Actions involving solely personnel time are the least expensive to implement (*i.e.*, mitigation, risk analyses), while actions requiring both personnel and laboratory consumables are more expensive (*i.e.*, testing), and actions requiring laboratory consumables, highly trained personnel, and prolonged instrument time are most expensive to implement (*i.e.*, viral sequencing).

Our findings are subject to statistical, methodological, and policy-based limitations. As with all studies of infectious disease surveillance, transmission events and clustering can violate statistical assumptions of independence among individuals. Additionally, while CMU had access to attribute data for individuals who never tested positive in the *Lookout* system, we were unable to access it for research under our existing regulatory approval; this impeded our ability to separate the impact that individual attributes (*e.g*., a particular sports team or residence hall) had on risk of infection. Moreover, incomplete sampling of residual diagnostic biospecimen and wastewater samples limited us to a partial snapshot of SARS-CoV-2 genetic diversity at CMU (Figure 2A; Appendix Figure 3C). Further, wifi co-location logs remain an underexplored data source that does not capture off-campus interactions and likely records superfluous or false interactions. As our study largely took place prior to the widespread availability of SARS-CoV-2 vaccines^45^ and of rapid at-home antigen tests^46^, we cannot assess their impact on infectious disease transmission or policy. Finally, CMU’s surveillance paradigm prioritized community safety over individual privacy; thus, some of our findings may not be generalizable to institutions with different prioritizations.

Accounting for resource constraints, we built upon CMU’s community-driven mindset to develop an efficient surveillance program and lay the groundwork for future advances. While a number of analyses here were conducted only retrospectively, updates to surveillance software like *Lookout*^30^ could enable timely identification of risk factors for infection and spread, proximity and location patterns, and lineages or mutations that are rising in frequency or that have been categorized as Variants of Concern. They could further refine the accuracy of outbreak reconstruction by incorporating genomic data including major and minor alleles, and contact tracing obtained from manual efforts or wifi analyses, with reported contacts weighted by the length or nature of the interaction. In summary, we propose the seamless, automated integration of multiple data types as the most powerful way to combat infectious disease outbreaks as they unfold.

## Methods

Code associated with the analyses of this study is available at: https://github.com/broadinstitute/sc2-cmu-study. Detailed methods are included in an appendix.

We identified factors associated with testing positive for SARS-CoV-2 *via* calculation of 1) relative risk for athletic participation, campus residency, and sex, and 2) Pearson’s chi-square test for distinct sports teams, residence halls, and class years. We also fit a linear model to residence hall incidence rates as a function of various binary and continuous attributes of the buildings (Appendix Figure 4A).

We examined differences in daily wifi connectivity patterns between subgroups defined by infection status or viral lineage and quantified differences in relevant metrics (Appendix Table 11) across subgroups via the Mann-Whitney U test. We calculated attribute assortativity (AA) to assess the tendency of individuals to associate with in-group *vs.* across groups, and determined the lag time that maximizes the correlation between AA and case count.

We constructed a network of known positive individuals, and connected pairs of individuals with interactions within 10 days of both positive tests. We compared wifi network path distances (i.e cumulative number of edges on the shortest path connecting two individuals in the network) between individuals in each pair where both were infected with B.1.429.1 lineage virus against pairs where only one individual had this viral lineage. We next compared viral genome distances (i.e., number of SNVs) within pairs of individuals infected with B.1.429.1 lineage virus that were connected by a path in the network *vs.* not connected.

Rolling 24-hour wastewater aliquots were collected from sewer sites to (1) monitor SARS-CoV-2 concentration via RT-qPCR and (2) provide source material for viral genomic sequencing. Across all collection sites, we compared weekly case counts from source buildings against daily and weekly wastewater viral titers (via Spearman correlation and Fisher’s exact test, respectively).

Excess material from clinical diagnostic tests and wastewater aliquots was sequenced following enrichment via PCR with ARTICv3-tiled amplicons.

LoFreq was used to call major and minor alleles within wastewater samples. Several quality control metrics were considered to discard spurious variation: minimum allele frequency, local sequencing depth, and concordance in variation across replicates^47^. Freyja was used to identify and estimate the abundance of constituent SARS-CoV-2 lineages^39^. We determined the average number of unique SNVs that a single additional sequenced sample contributes to a set of a given number of samples, via bootstrapping of distinct subsets of clinical or wastewater samples.

Clinical genomes were assembled using the Broad Institute viral-ngs pipelines and Nextstrain pipelines and assigned Pango lineages^34, 48^. Genomes were placed in the phylogenetic context of genomes from cases outside the university community, weighted toward genomes from Colorado and surrounding states. Case clusters were identified based on a state change from not university-associated to university-associated, following ancestral state reconstruction of internal tree nodes.

Negative binomial distributions were fit to distributions of cluster size, the number of contacts in the wifi data set, and the number of contacts in the standard contact tracing data set. Contacts were defined as any interaction >15 minutes in the 48-hour period prior to testing or symptom onset.

Transmission networks were inferred using sequenced viral genomes and contact data using *outbreaker2*^49^.

Lentiviral pseudotype assays were performed to assess the functional implications of the S:Q677H substitution on infectivity and fusogenicity.

## Data Availability

All non-identifiable data produced in the present work are contained in the manuscript. Genomic sequences were deposited in NCBI GenBank, GISAID, and the Sequence Read Archive, as applicable.

https://github.com/broadinstitute/sc2-cmu-study

## Funding Acknowledgments

This work was supported by the National Science Foundation (NSF) Graduate Research Fellowship Program to J.S.P. (grant number 1745303), and C.H.T-T. (grant number 1745303); the Hertz Foundation to J.S.P.; the National Institute of General Medical Sciences to B.A.P. (grant number T32GM007753); the National Science Foundation (NSF) to B.C.L. (grant number 2046440); the Moore Foundation to B.C.L., the National Institute of Allergy and Infectious Diseases to B.C.L. (grant number 2U19AI110818) and to P.C.S. (grant number U19AI110818 and U01AI151812); the National Institutes of Health to J.L. (grant numbers R37AI147868 and R01AI148784); the Massachusetts Consortium on Pathogen Readiness to J.L.; the Centers for Disease Control and Prevention (CDC) COVID-19 baseline genomic surveillance contract to the Clinical Research Sequencing Platform (grant number 75D30121C10501) and a CDC Broad Agency Announcement to B.L.M. (grant number 75D30120C09605); and the Howard Hughes Medical Institute to P.C.S. This work is also made possible by support from Flu Lab and a cohort of donors through the Audacious Project, a collaborative funding initiative housed at TED, including The ELMA Foundation, MacKenzie Scott, the Skoll Foundation, and Open Philanthropy.

Any opinion, findings, and conclusions or recommendations expressed in this material are those of the author(s) and do not necessarily reflect the views of the National Science Foundation. This content is solely the responsibility of the authors and does not necessarily represent the official views of the National Institute of General Medical Sciences or the National Institutes of Health.

## Acknowledgements

The authors wish to thank Jacob E. Lemieux for helpful discussions and feedback. The authors also wish to thank Laurent Hébert-Dufresne for providing insights and feedback on the wifi co-location analyses.

## Ethics Statement

The study was conducted at the Broad Institute with approval from the MIT Institutional Review Board under Protocol #1612793224 and from WCG IRB Protocol #20210166 under Protocol: Investigating Viral Emergence and Spread in Community Settings.

---

**Appendix Figure 1.**
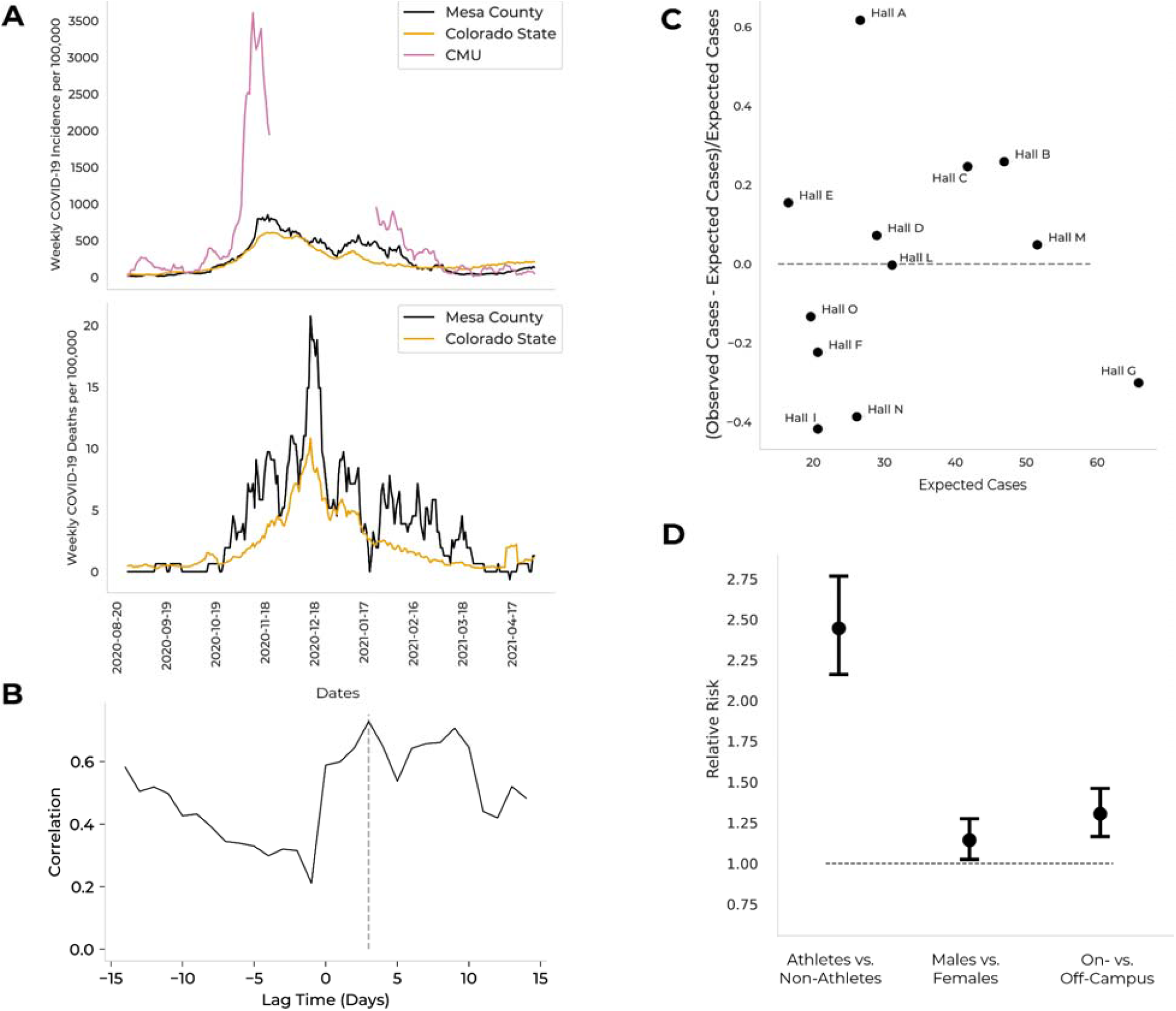
Incidence rates at Colorado Mesa University (CMU), with surrounding areas for context, and epidemiological risk factors for SARS-CoV-2 positivity at CMU **A.** Weekly COVID-19 incidence at Colorado Mesa University (CMU; pink), in Mesa County (black), and in Colorado (orange) over the 2020–2021 academic year (upper). Weekly COVID-19 death rate in Mesa County (black) and in Colorado (orange) over the 2020–2021 academic year (lower). Data from CMU were not analyzed from November 21–January 18 due to winter recess. The correlations between CMU incidence rates and Mesa County incidence rates (correlation = 0.73, lag = 3 days) and Mesa County death rates (correlation = 0.48, lag = 8 days) were tested with lag times between −14 days (0 days for deaths) and 14 days, and the maximum correlations and corresponding lag times are reported. **B.** The correlation between CMU incidence rates and Mesa County incidence rates *vs.* the lag time. The dashed line at *y* = 3 indicates the lag time that maximizes the cross-correlation. **C.** The difference between the number of cases observed and the number of cases expected (given residence hall population size and scaled by number of cases expected) *vs.* the number of cases expected, per residence hall. The dashed line at *y* = 0 separates halls with more (above) or fewer (below) cases observed than expected. **D.** Relative risk (RR) of testing positive for COVID-19 given sports team membership (athletes *vs.* non-athletes), sex (males *vs.* females), or residential status (on-*vs.* off-campus). Circles represent the relative risk, with whiskers extending to the upper and lower bounds of the 95% confidence interval. The dashed line at RR = 1 represents the null hypothesis (*i.e.*, that there is no association between the risk factor and test positivity). An RR greater than 1 implies that the first group listed (*i.e.*, athletes, males, or on-campus students) had a greater risk of testing positive than the second group listed (*i.e.*, non-athletes, females, or off-campus students).

**Appendix Figure 2.**
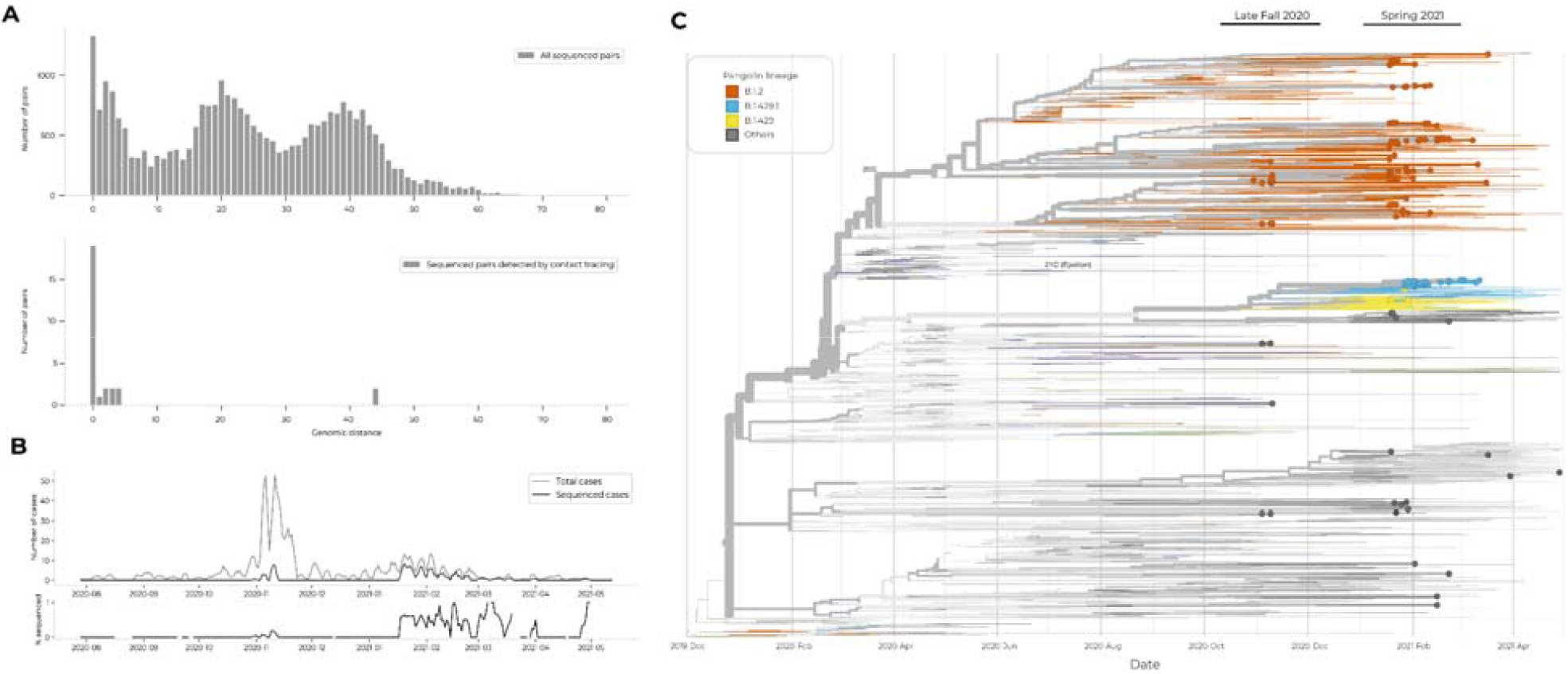
Detection of outbreak groups using viral genomic sequencing of cases **A.** Distributions of genetic distance for all pairs of viral genomes (upper) and for pairs of viral genomes from positive individuals sharing interactions reported during contact tracing (lower). We sequenced viral samples from 18 pairs of positive individuals where one of the individuals identified the other as a close contact, and 11 pairs where both individuals identified one another as a close contact. Two outlier pairs had a genetic distance of 44 SNVs. In each of these pairs, an individual testing positive at the end of January 2021 had virus of the B.1.2 lineage and indicated close contact with an individual who tested positive in early February (5-6 days later) with virus of the B.1.429.1 lineage. **B.** Sequencing of positive cases via genomic surveillance across both semesters. Sequencing began October 2020, capturing cases associated with a Halloween-related outbreak. Shown are the total number of cases (top, thin line), total number of sequenced cases (top, bolded line), and percent of positive cases that were sequenced (bottom). **C.** Phylogenetic placement and temporal sampling of viral genomes sequenced from CMU clinical diagnostic tests. CMU nodes are depicted as dots among a global tree of contextual sequences, weighted toward sequences collected from Colorado and surrounding states, but including uniformly-sampled genomes from all dates and geographic regions, as well as genomes close in genetic distance to CMU sequences. Interactive version available at https://auspice.broadinstitute.org.

**Appendix Figure 3.**
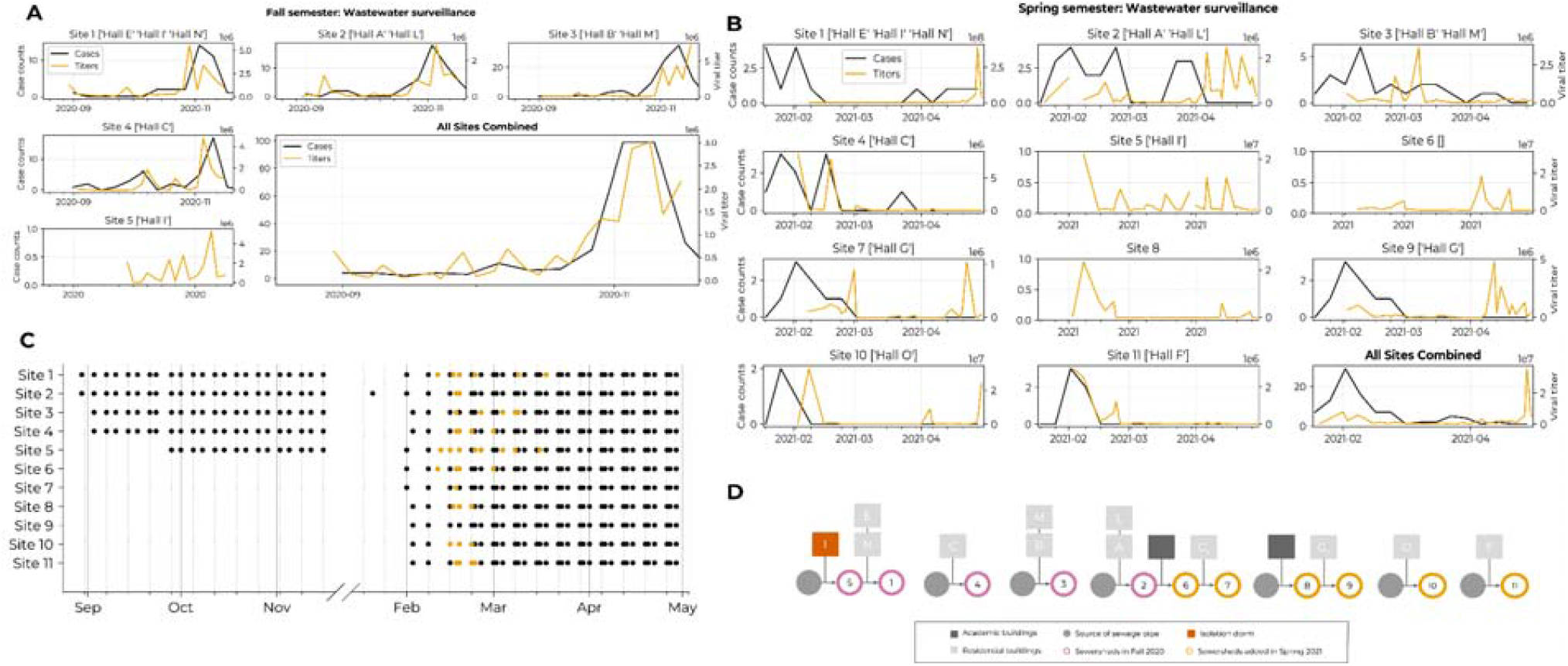
Summary of wastewater surveillance implemented at Colorado Mesa University across Fall 2020 and Spring 2021 **A-B.** Comparison of wastewater viral titer (orange) and weekly case count (black) for each wastewater site in the Fall semester (**A**) and Spring semester (**B**). **C.** Sample collection frequency for wastewater surveillance in the 2020–2021 academic year. Wastewater samples were collected twice weekly in the Fall across 5 sites, and three times weekly in the Spring across 11 sites. Each dot represents an independent collection event, with dots shown in orange representing sequenced wastewater samples. **D.** Schematic showing which residential or academic buildings (squares) contributed effluent to each monitored wastewater site (circles). Residence hall I (red square) housed a small number of students who had tested positive and were in isolation. Wastewater sites are represented by circles, with filled-in gray circles representing sites utilized to collect baseline measurements where there were no upstream contributors to that particular sewage system. Sites shown in pink were added in Fall 2020 and remained in use into Spring 2021, while sites shown in orange were added in Spring 2021.

**Appendix Figure 4.**
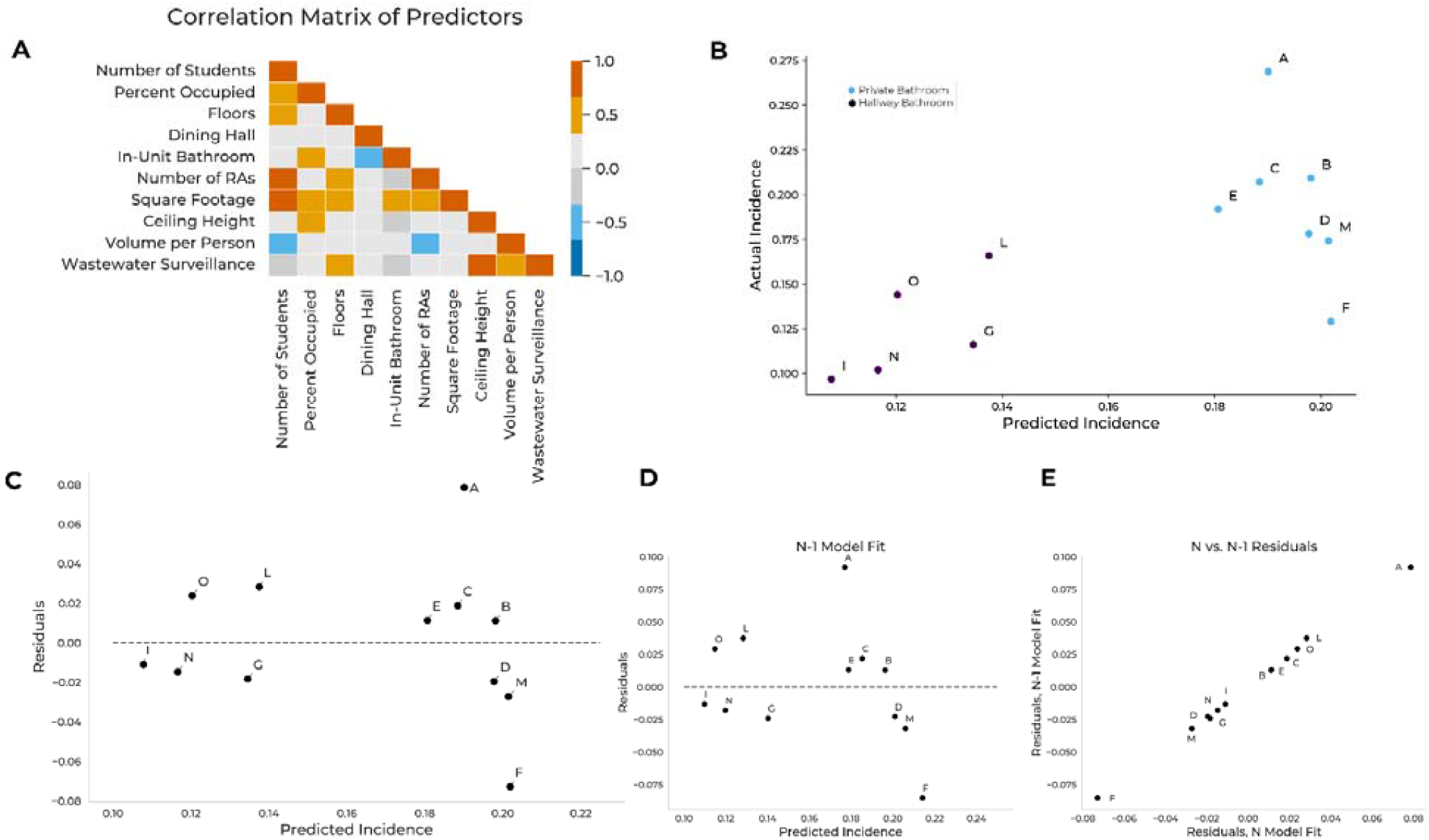
Predictions of COVID-19 incidence rates in Colorado Mesa University residence halls *via* a linear regression model **A.** Descriptive attributes of residence halls, including number of students, percent occupied (number of available beds / number of students), number of floors, the presence of a meal plan requirement (‘dining hall’), the presence of an in-unit bathroom (‘private bath’), the number of resident advisors (RAs), square footage, ceiling height, volume per person, and the presence of wastewater surveillance, were used as predictors in the regression model. Non-zero correlation coefficients between pairs of predictive variables indicate multicollinearity. **B.** Observed incidence rates *vs.* predicted incidence rates, by residence hall. Predictions come from the model that minimized the Akaike and Bayesian Information Criteria. Colors indicate the difference in predicted incidence associated with a private *vs.* a hallway bathroom. **C.** Residuals *vs.* predicted incidence rates, by residence hall. Root-mean-square-error = 3.6%. **D.** Residuals *vs.* predicted incidence rates, from leave-one-out cross-validation, by residence hall. Leave-one-out cross-validation (*i.e.*, *N*-1 model fit, where *N* = number of residence halls) was conducted such that each hall’s incidence rate was predicted *via* a linear model whose coefficients were determined with training data from all other halls. Root-mean-square-error = 4.2%. **E.** Residuals from the leave-one-out cross-validation (*N*-1 model fit) *vs.* residuals from the full model (*N* model fit).

**Appendix Figure 5.**
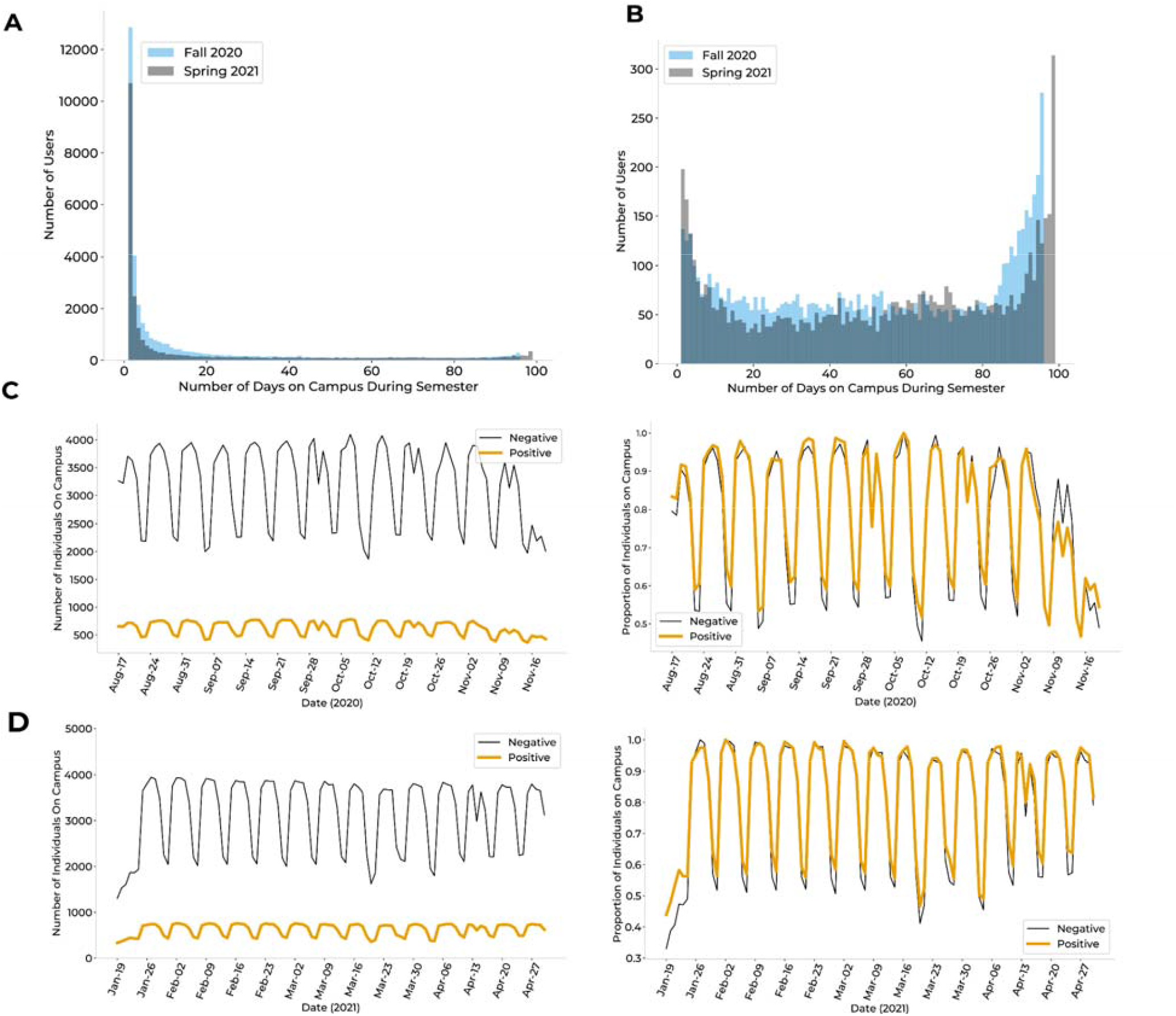
On-campus presence, as inferred by wifi proximity data, by semester and by user category **A.** Distribution of the number of days present on campus for all users found in the wifi network, colored by semester. **B.** Distribution of the number of days present on campus for all users found in the wifi network after removal of unauthenticated users, non-students, and infrequent users, colored by semester. This cleaned wifi proximity network was used for all future analyses. **C.** Number (left) and proportion (right) of positive students (orange) and negative students (black) on campus by day for Fall 2020 (Appendix Table 11). There was no significant difference in the proportion distributions (p = 0.282). We found a significant correlation in the trends of the proportion distributions (Pearson Correlation: .976, p < .0001). **D.** Number (left) and proportion (right) of positive students (orange) and negative students (black) on campus by day for Spring 2021 (Appendix Table 11). There was no significant difference in the proportion distributions (p = 0.248). We found a significant correlation in the trends of the proportion distributions (Pearson Correlation: .99, p < .0001).

**Appendix Figure 6.**
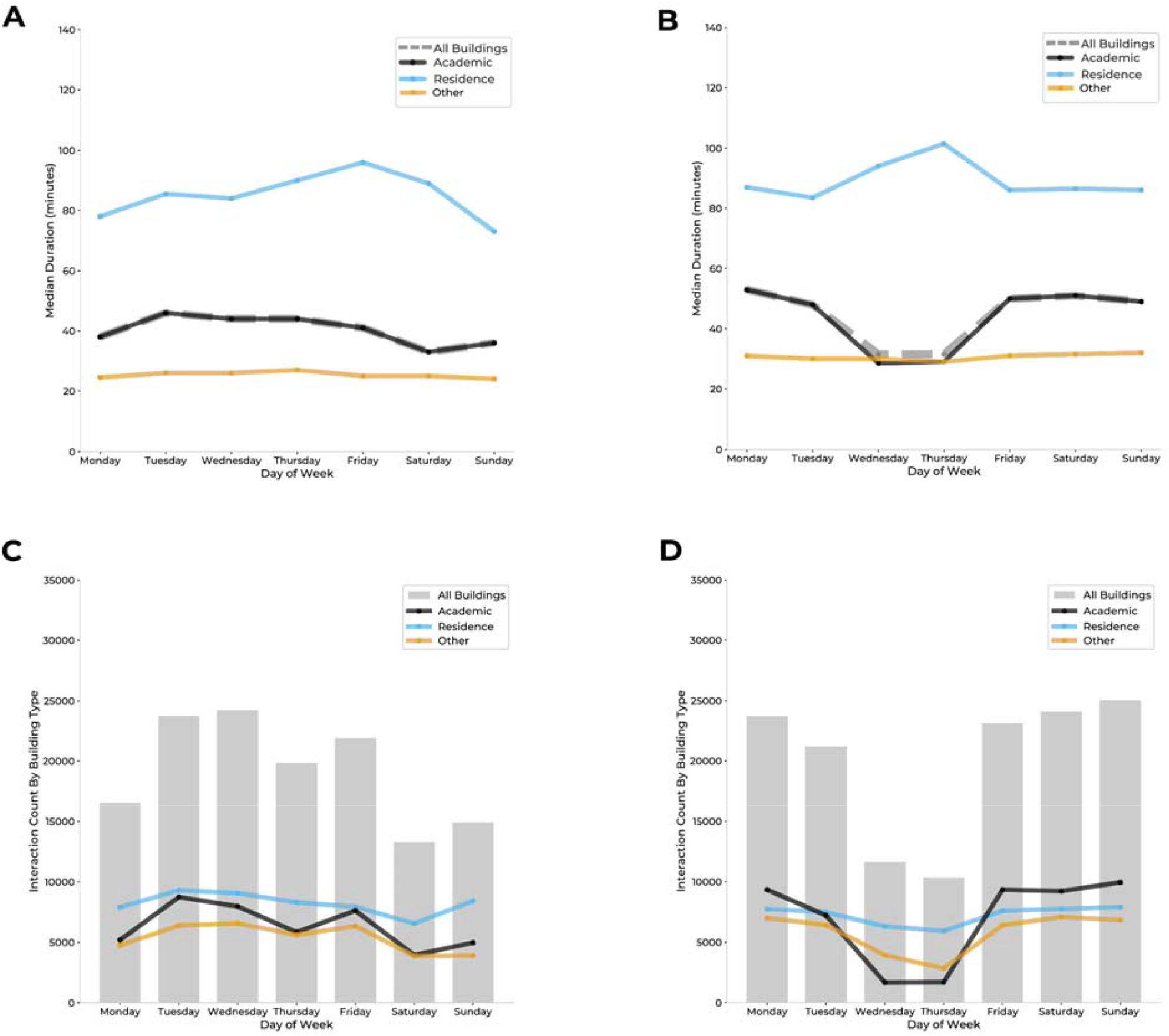
Wifi network access patterns by building and by semester. **A-B.** Median duration of access point connections in minutes, per day of the week and by building type (academic, residential, other, or all buildings) for Fall 2020 (**A**) and Spring 2021 (**B**; Appendix Table 11). **C-D.** Daily number of AP connections, per day of the week and by building type (academic, residential, other, or all buildings) for Fall 2020 (**C**) and Spring 2021 (**D**; Appendix Table 11).

**Appendix Figure 7.**
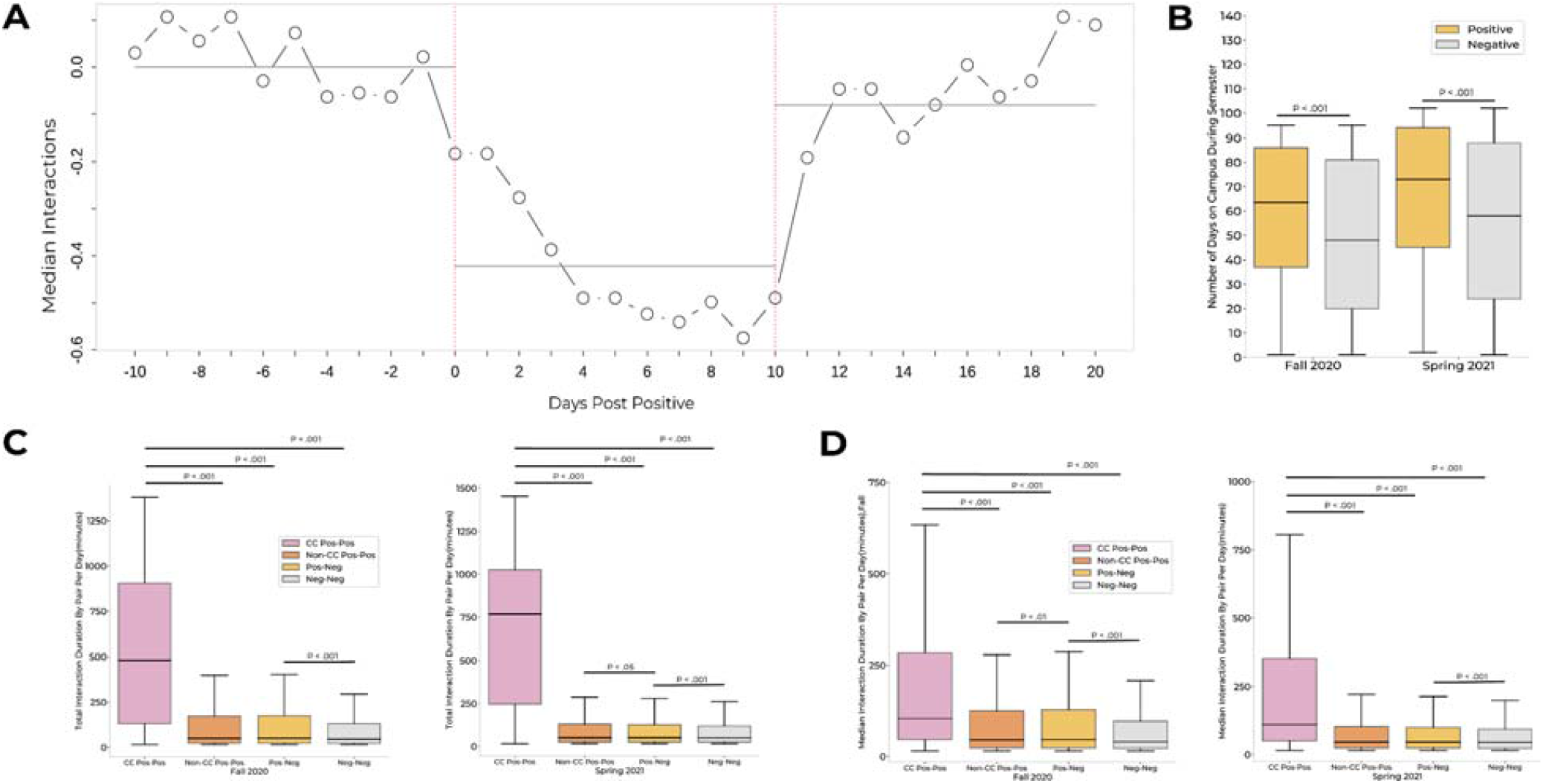
Interaction patterns of individuals and of pairs of individuals by testing status **A.** Median number of wifi contacts per day for the 30-day period surrounding the isolation period of positive cases. Day 0 was calculated as the earliest of symptom onset date and test date. Black lines indicate the median number of contacts (y-axis) per individual for every day across the period (x-axis). **B.** The number of days on campus for positive (orange) *vs.* negative (gray) individuals. Effect sizes, 15 days (median positive: 66; median negative: 51; Fall 2020) and 16 days (median positive: 76; median negative: 60; Spring 2021). Gray bars indicate averages for three 10-day periods: pre-isolation, isolation, and post-isolation. **C.** Distributions of total pairwise daily interaction duration, for pairings categorized by testing status: CC Pos-Pos, pairs of pre-positive individuals with an association reported in manual contact tracing; Non-CC Pos-Pos, pairs of pre-positive individuals without an association reported in manual contact tracing; Pos-Neg, pairs of one pre-positive and one negative individual; Neg-Neg, pairs of two negative individuals. Pre-positive individuals were defined as students within the 10-day window prior to testing positive. **D.** Distributions of median interaction duration by pair per day, for pairings categorized by test status (see above description). Positive individuals were defined as students within the 10-day window prior to testing positive. See Appendix Table 11 for equations used in panels **A**, **C**, and **D**.

**Appendix Figure 8.**
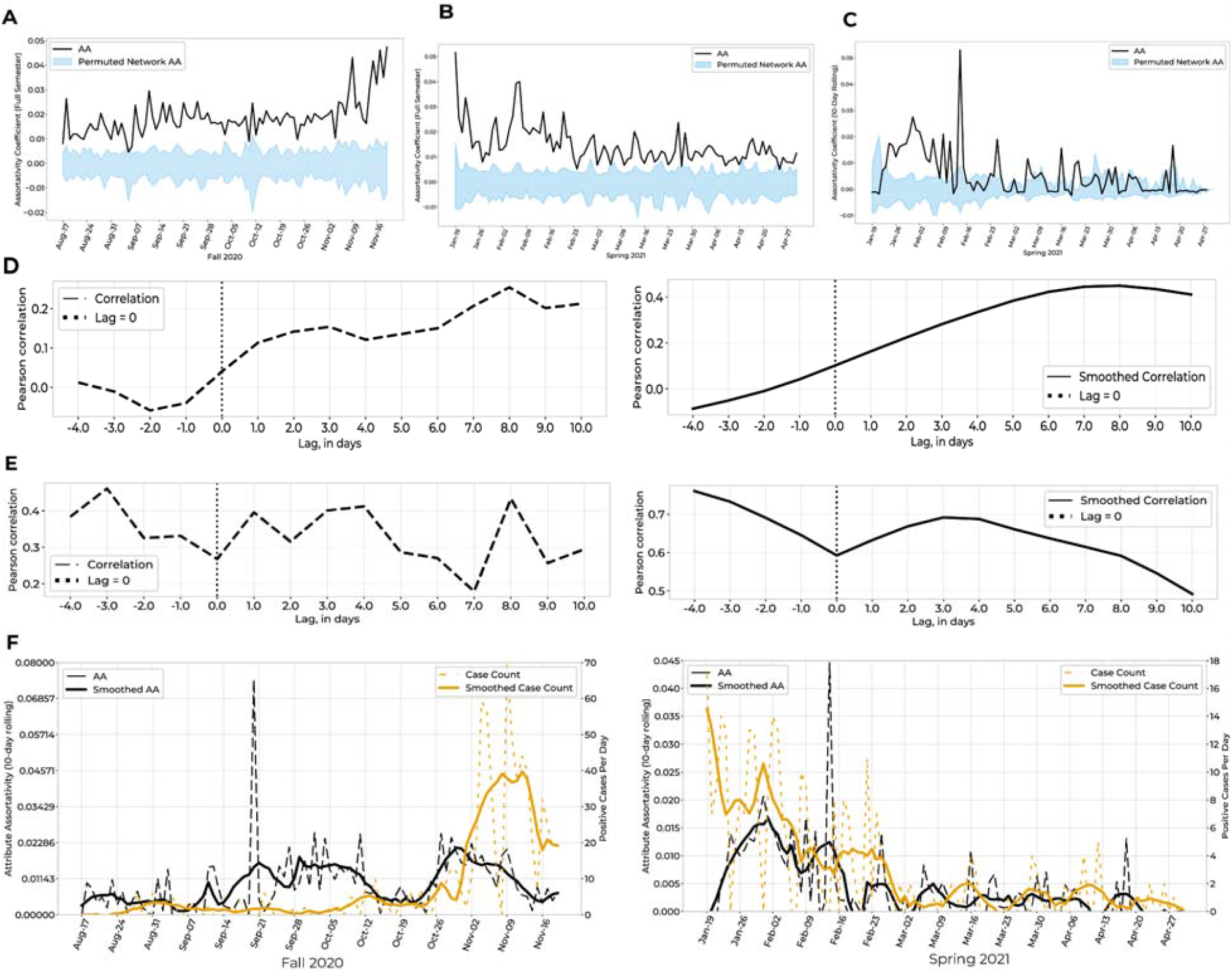
Comparison of randomness vs true assortativity within the wifi proximity network in conjunction with semester daily case counts **A-B.** Comparison of attribute assortativity (AA) for positives *vs*. negatives, for Fall 2020 (**A**) and Spring 2021 (**B**). 95% Confidence Intervals (CI; blue) were calculated by permuting positive and negative labels for individuals within the proximity network each day. The AA of the proximity network (black) was above the upper bound of the CI for 98.9% (94/95 days) in the Fall 2020 semester and 98%(100/102 days) in the Spring 2021 semester, indicating significance with p < 0.025. **C.** We calculated the same metric with altered groupings: pre-positives (all individuals within a 10-day window prior to their positive test) *vs.* negatives (individuals not within a 10-day window prior to a positive test, regardless of overall semester testing status). The AA of the proximity network (black) was above the upper bound of the CI for 37.2% (38/102 days) in the Spring 2021 semester, indicating significance with p < 0.025. **D.** Correlations between daily AA (pre-positives *vs*. negatives) and daily case count with varying lag times, where both AA and case count are non-smoothed (top) or smoothed using Savitzky Golay filter (bottom), for Fall 2020. The highest correlation occurs with a lag time such that AA preceded the case count by 8 days (Pearson correlation = 0.253 for non-smoothed, 0.449 for smoothed). **E.** Cross-correlations between daily AA (pre-positives *vs*. negatives) and daily case count with varying lag times, where both AA and case count are non-smoothed (top) or smoothed using Savitzk Golay filter (bottom), for Spring 2021. The highest correlation occurs with lag time such that AA preceded the case count by 4 days (Pearson correlation=0.411 for non-smoothed; 3 days for smoothed, with correlation = 0.691). **F.** Unsmoothed case counts and unsmoothed AA *vs.* smoothed case counts and smoothed AA for Fall 2020 (left) and Spring 2021 (right).

**Appendix Figure 9.**
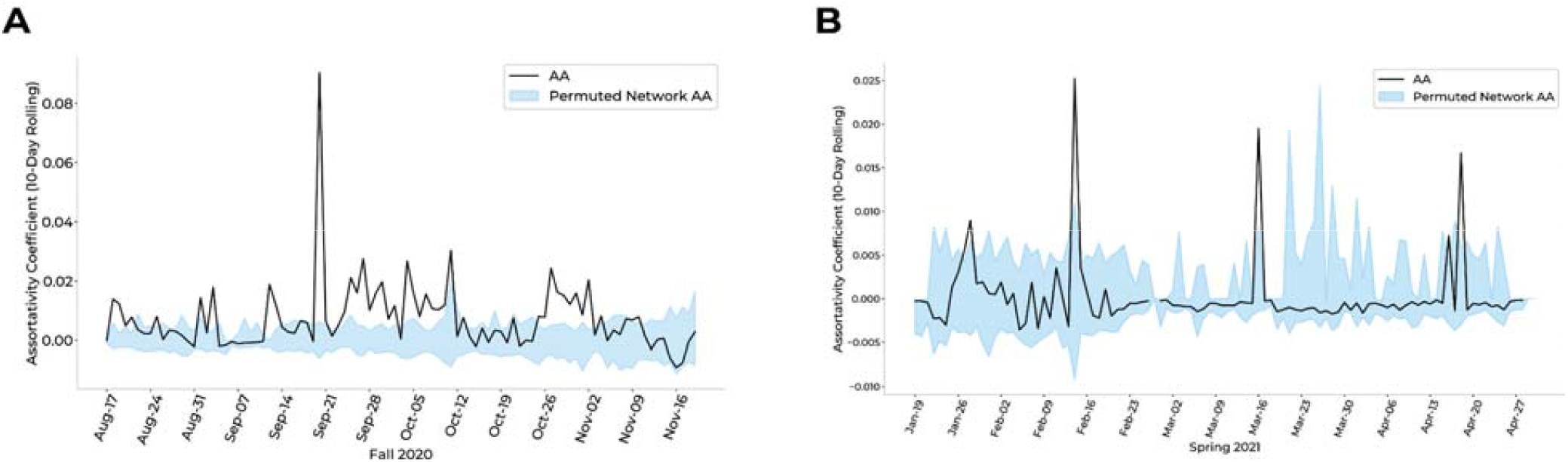
Comparison of assortativity within the wifi proximity network when defining the “pre-positive” attribute as positive individuals without pairwise associations within the contact tracing data. **A-B.** Comparison of attribute assortativity (AA) for pre-positives *vs*. negatives, for Fall 2020 (**A**) and Spring 2021 (**B**). 95% Confidence Intervals (CI; blue) were calculated by permuting pre-positive and negative labels for individuals within the proximity network each day. We calculated the same metric with pre-positives defined as individuals within a 10-day window prior to their positive test without pairwise associations with other pre-positive individuals in the contact tracing data and negatives defined as individuals not within a 10-day window prior to a positive test, regardless of overall semester testing status. The AA of the proximity network (black) was above the upper bound of the CI for 45.2% (43/95 days) of the Fall 2020 semester (**A**) and 8.8% (9/102 days) of the Spring 2021 semester (**B**), indicating significance with p < 0.025.

**Appendix Figure 10.**
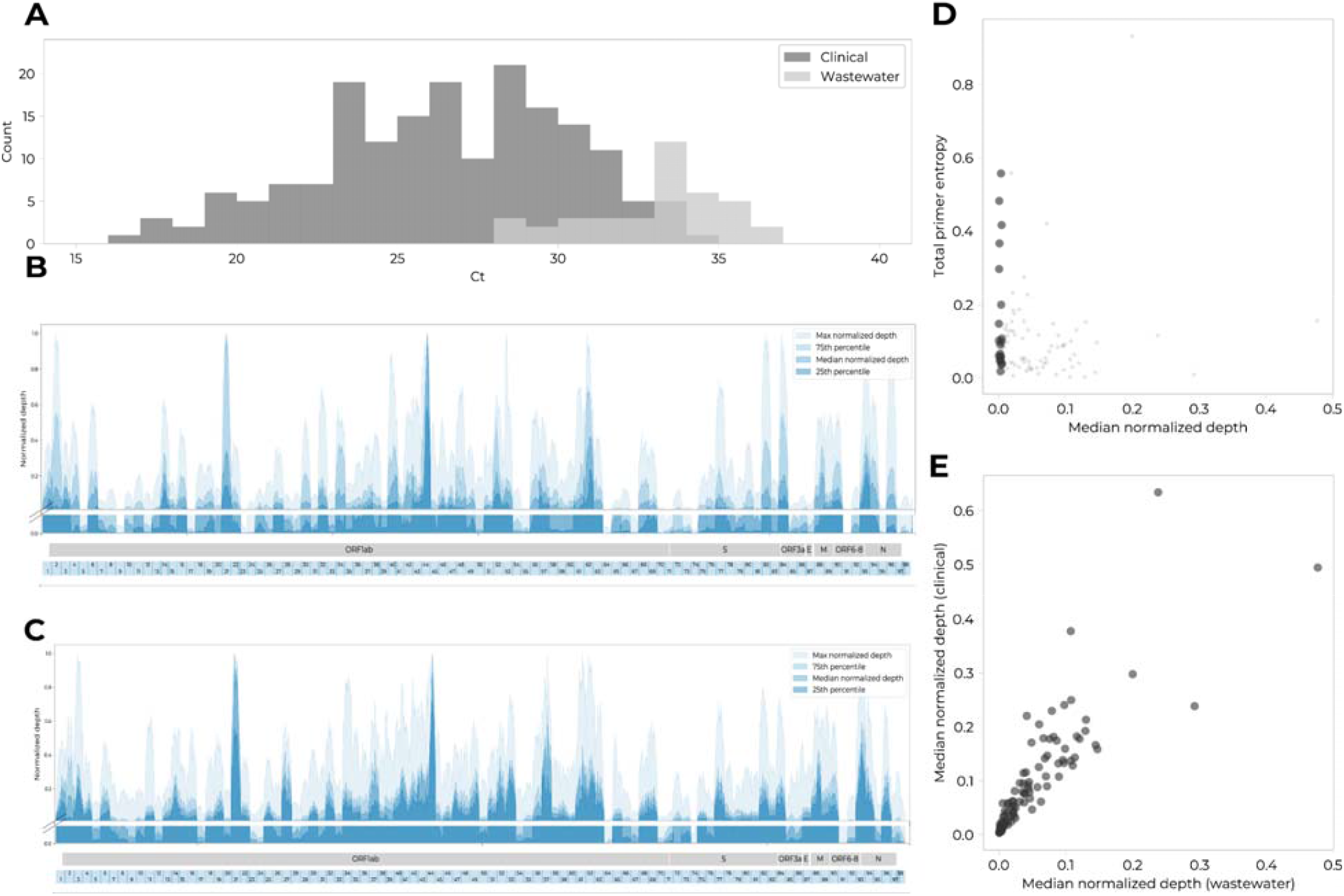
Sequenced wastewater samples, though subject to increased degradation, are of similar genome coverage as sequenced clinical samples **A.** Cycle thresholds (Ct) for sequenced wastewater and clinical samples from Colorado Mesa University. **B-C.** Median normalized read depth per base, across all wastewater (**B**) and clinical (**C**) samples. The coloring indicates quartiles across all wastewater or clinical samples. Regions of lower depth in wastewater sequencing are of similarly low depth in clinical sequencing. **D.** Comparison of average amplicon read depth (*x* axis) and average amplicon primer entropy (*y* axis) for wastewater samples. We found no correlation between amplicon read depth and primer entropy (Pearson correlation coefficient of r = .02, p = 0.81). The 20 amplicons with lowest median normalized depth are shown as larger circles. **E.** Comparison of median normalized depth per amplicon between wastewater (*x* axis) and clinical (*y* axis) samples. There is a linear correlation between corresponding amplicons (Pearson correlation coefficient of r = 0.86; p<0.001).

**Appendix Figure 11.**
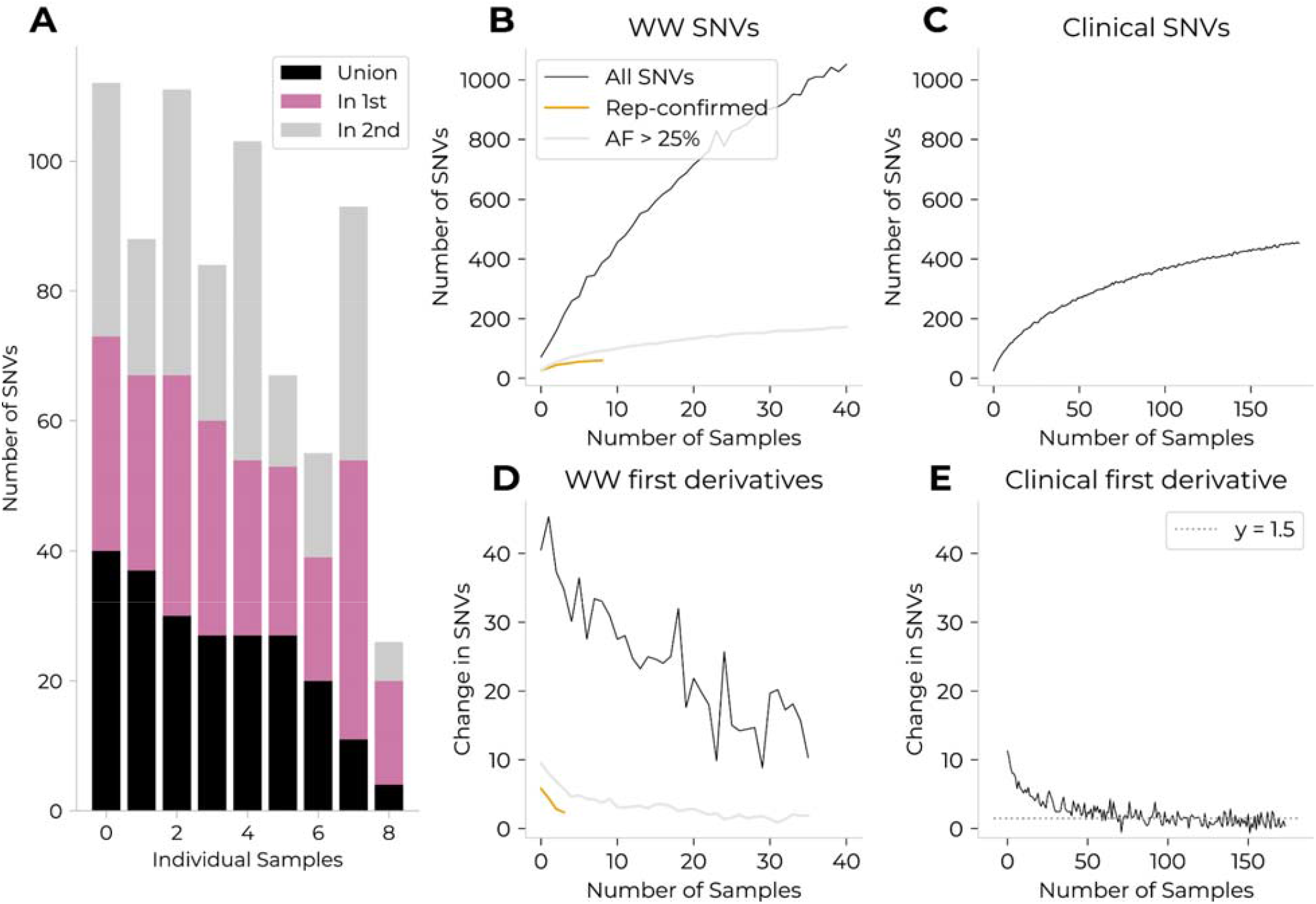
Presence of previously undetected mutations in sequenced wastewater samples suggests sequencing error **A.** Distribution of single-nucleotide variants (SNVs) that were present in only one technical duplicate (pink and gray) or in both duplicates (black). The samples are ordered relative to the number of replicate-confirmed SNVs. **B.** Predictions for the number of unique SNVs that can be expected for a given number of wastewater samples, calculated per the methodology described in the Supplemental Methods. We additionally calculated the number of unique SNVs that would be expected to pass an allele frequency quality control threshold of 25% (gray) or to be seen in at least both technical replicates of any given sample (orange). **C**. Predictions for the number of unique consensus-level SNVs that can be expected for a given number of clinical samples, calculated per the methodology described in the Supplemental Methods. **D.** Smoothed (window=5) first derivative of unique wastewater SNVs per sample. Legend is the same as in panel (**B**). This highlights that while the first derivatives for both replicate-confirmed SNVs (orange) and SNVs with an allele frequency greater than 25% (gray) approaches a rate of 1-3 new SNVs per sample, the first derivative for all SNVs of any allele frequency (black) approaches a rate of >10 new SNVs per sample. **E.** Smoothed (window=5) first derivative of unique clinical consensus-level SNVs per sample. This highlights that the first derivative for all consensus-level SNVs seen in clinical samples approaches a rate of 1.5 new SNVs per sample. The comparison between (**D**) and (**E**) lends support to the hypothesis that most SNVs seen in wastewater samples are spurious, and that either confirming SNVs via technical replicates or a high allele frequency threshold is sufficient to remove most spurious SNVs.

**Appendix Figure 12.**
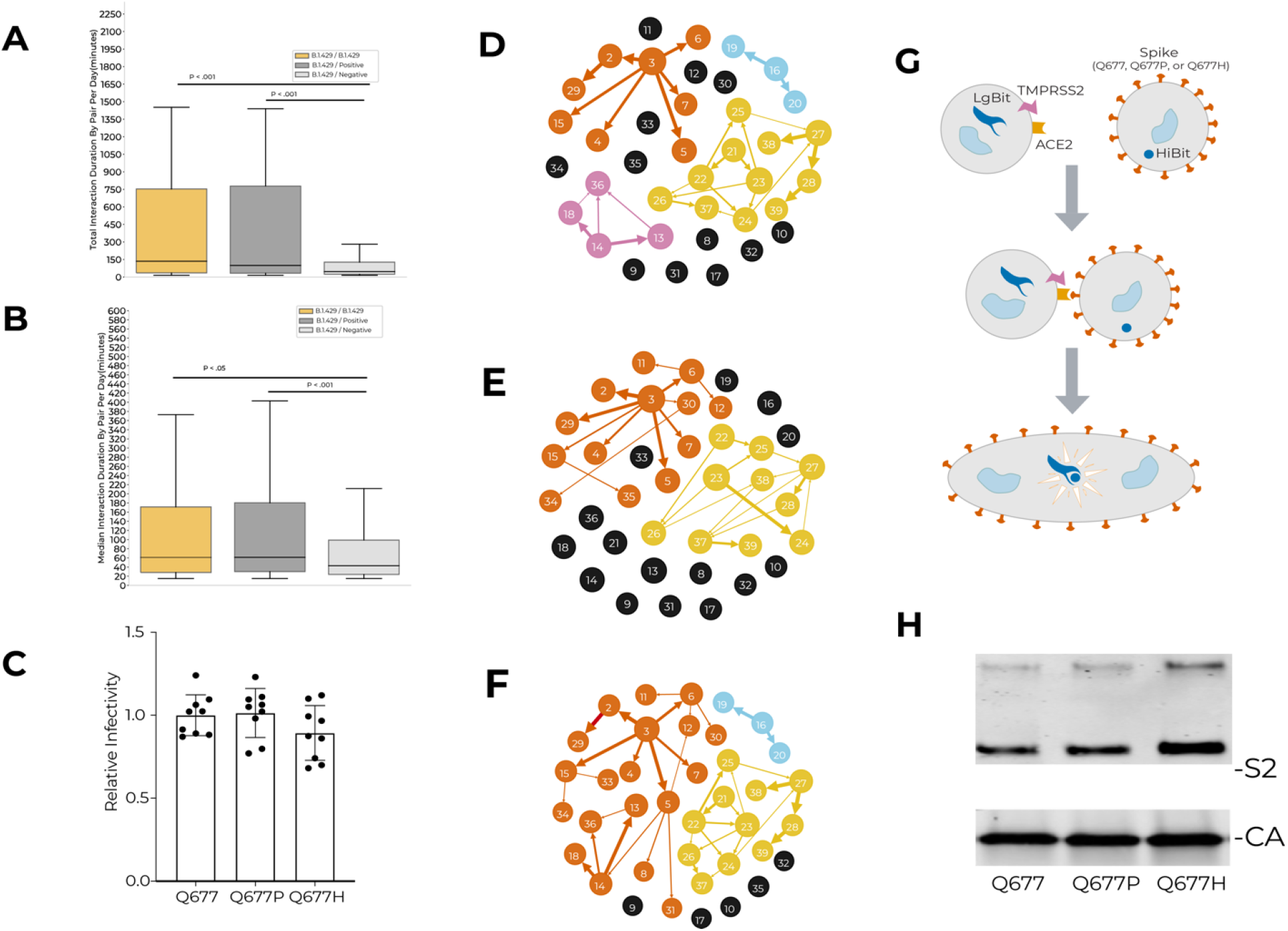
Social proximity patterns, transmission reconstruction networks, and experimental viral phenotypes associated with B.1.429.1 **A.** Distributions of total daily interaction duration (Appendix Table 11), per pair and per day, for pairs of B.1.429.1-positive (within 10 days of positive tests), non-B.1.429.1-positive (within 10 days of positive tests), and negative individuals. Non-B.1.429.1-positive individuals are defined as positive individuals whose viral genome was not sequenced or was sequenced and determined not to be B.1.429.1. **B.** Distributions of median daily interaction duration (Appendix Table 11), per pair per day, for pairs of B.1.429.1-positive (within 10 days of positive tests), non-B.1.429.1-positive (within 10 days of positive tests), and negative individuals. **C.** Results of infectivity of viral pseudotypes with the ancestral allele, or with the S:Q677P or S:Q677H amino acid changes, relative to a luminescent control with no Spike protein expressed. **D-F.** Transmission reconstruction network for B.429.1 cases created with solely genomic information (**D**), solely manual contact tracing data (**E**), and both genomic information and wifi-inferred 2-day contact data (**F**). **G.** Conceptual diagram showing the experimental strategy for assessing the fusogenicity of the S:Q677H or S:Q677P amino acid changes relative to the ancestral residue, using two populations of cells: one expressing the modified Spike protein on the cell surface, and a second expressing the ACE2/TMPRSS2 receptors. The two populations were combined and fusion signal was assessed via HiBit-LgBit luminescent reporter. **H.** Western blot illustrating successful creation of viral pseudotype particles bearing Spike protein with S:677Q, S:Q677H, and S:Q677P.

**Appendix Table 1.**
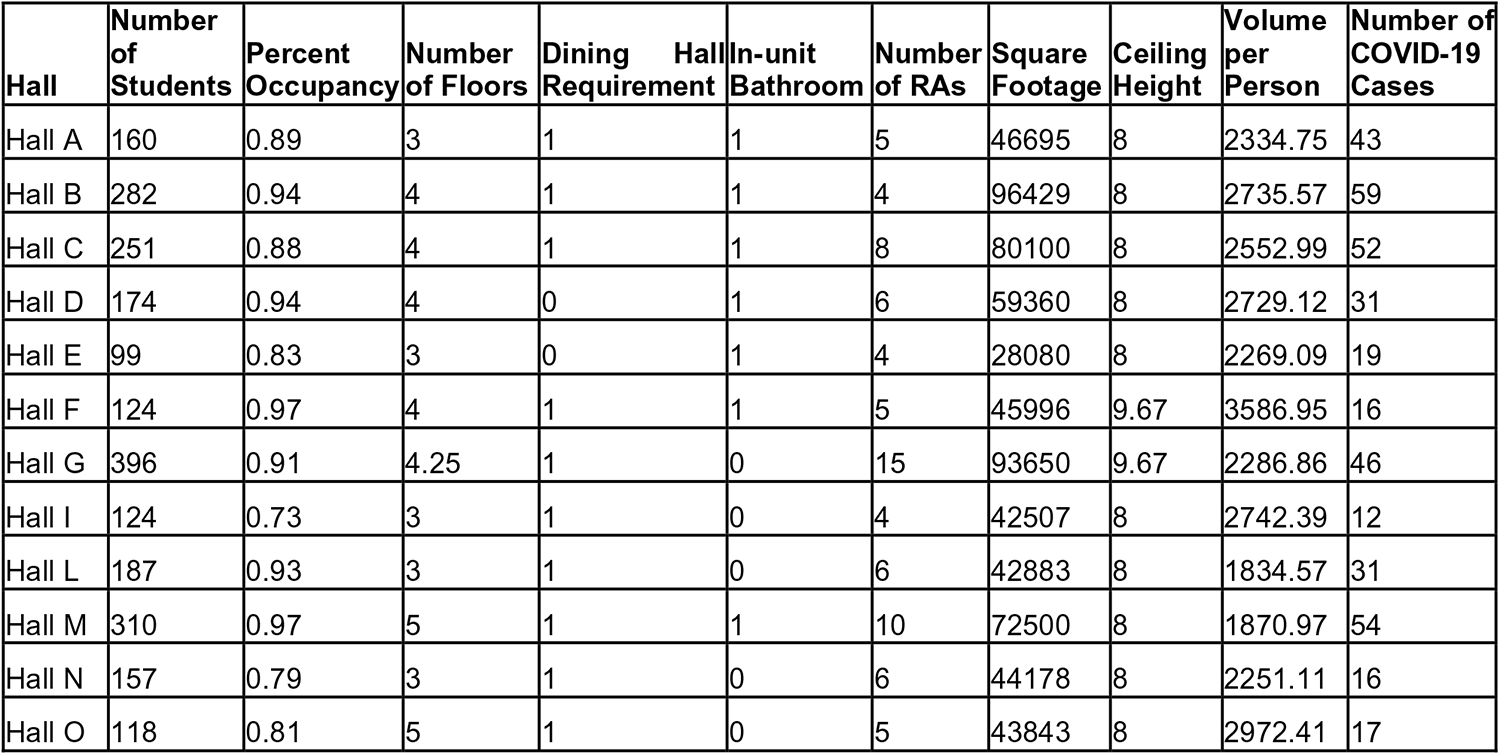
Properties of residence halls at Colorado Mesa University Structural and communal properties of residence halls. Volume per person was calculated as square footage * ceiling height / number of students per hall and serves as a proxy for air volume per person. Hall G has 4 floors and a partial basement, covering approximately 25% of the square footage of the other floors.

**Appendix Table 2.** Properties of athletic teams at Colorado Mesa University Number of students, contact level (as determined via standardized definitions in **Appendix Table 3**), season, location, and number of COVID-19 cases per sports team. Teams with fewer than 5 cases were labeled as “< 5” to minimize the risk of de-anonymity. Teams are sorted by number of COVID-19 cases, then by contact level.

| Team | Number of Students | Contact Level | Season | Location | Number of COVID-19 Cases |
| --- | --- | --- | --- | --- | --- |
| Women's Golf | 11 | Low | both | outdoor | <5 |
| Women's Club Volleyball | 11 | Low | both | indoor | <5 |
| Men's Golf | 12 | Low | both | outdoor | <5 |
| Women's Tennis | 12 | Low | both | outdoor | <5 |
| Women's Cross Country | 14 | Low | fall | outdoor | <5 |
| Men's Triathlon | 18 | Low | both | outdoor | <5 |
| Cycling | 65 | Low | both | outdoor | <5 |
| Women's Swimming | 33 | Low | both | indoor | 6 |
| Men's Tennis | 10 | Low | both | outdoor | 7 |
| Women's Triathlon | 14 | Low | both | outdoor | 7 |
| Cheerleading | 30 | Moderate | both | indoor | 7 |
| Men's Soccer | 30 | High | fall | outdoor | 8 |
| Sand Volleyball | 19 | Moderate | spring | outdoor | 8 |
| Women's Basketball | 15 | High | both | indoor | 9 |
| Women's Wrestling | 22 | High | both | indoor | 9 |
| Indoor Volleyball | 20 | Moderate | fall | indoor | 9 |
| Men's Track | 55 | Moderate | both | both | 10 |
| Men's Basketball | 19 | High | both | indoor | 11 |
| Men's Swimming | 38 | Low | both | indoor | 11 |
| Women's Lacrosse | 30 | Moderate | spring | outdoor | 12 |
| Women's Soccer | 35 | High | fall | outdoor | 13 |
| Women's Track | 55 | Moderate | both | both | 13 |
| Baseball | 48 | Moderate | spring | outdoor | 14 |
| Men's Lacrosse | 43 | Moderate | spring | outdoor | 17 |
| Men's Wrestling | 40 | High | both | indoor | 23 |
| Football | 95 | High | fall | outdoor | 48 |

**Appendix Table 3.**
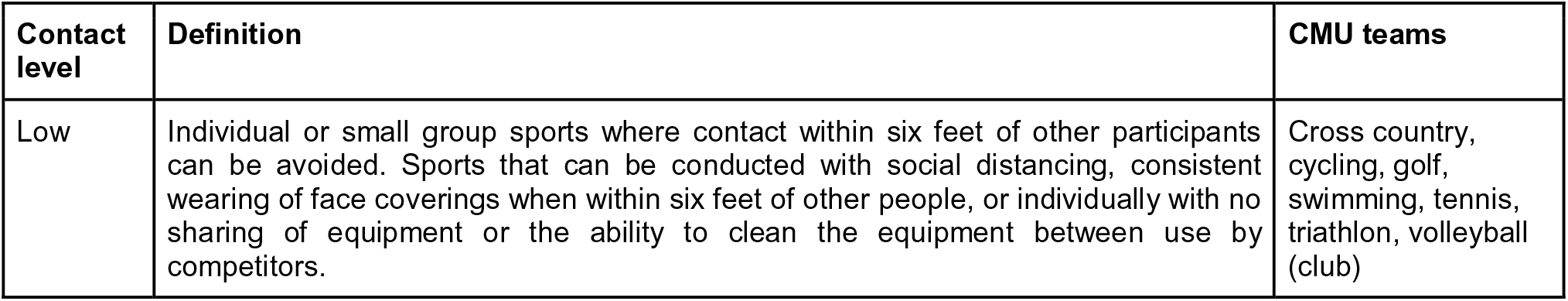
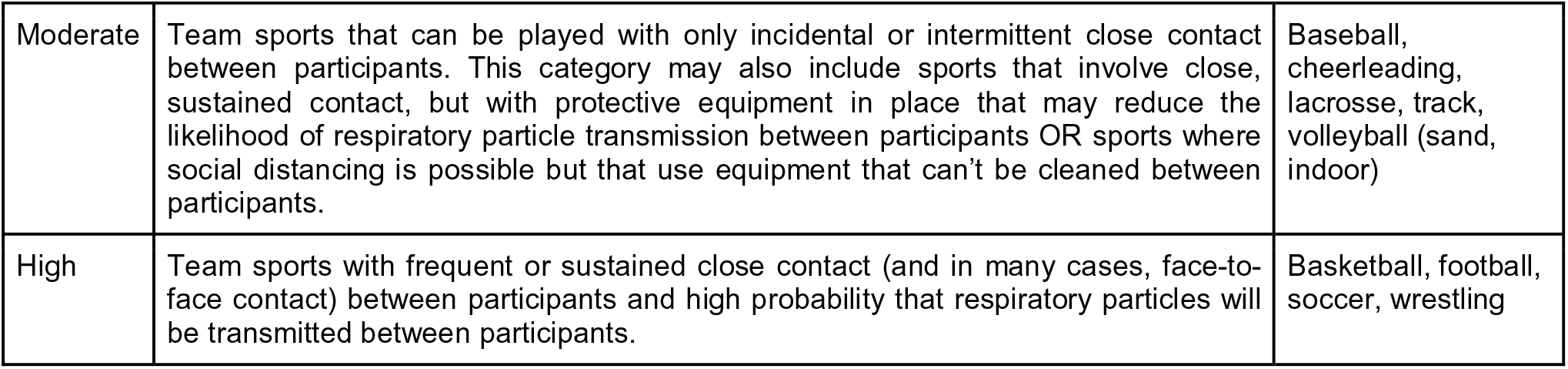
Sports contact level definitions. Definitions of contact levels for sports teams. Sports teams were assigned a contact level of low, medium, or high based on variables that affect transmission risk, including physical proximity and the use of face coverings. Contact-level categories were created by synthesizing risk profiles defined from the California Department of Health’s “Outdoor and Indoor Youth and Recreational Adult Sports” communication in April 2021 and the Colorado High School Activities Association 2020-2021 sports risk profiles.

**Appendix Table 4.** Chi-square analyses. Results of Pearson’s chi-squared test for categorical variables assessed for COVID-19 disease risk. Test statistics, degrees of freedom, and uncorrected p-values are reported.

| Variable | Chi-Square Statistic (df) | P-value (Uncorrected) |
| --- | --- | --- |
| Class years (Fall 2020) | 95.67 (5) | $4.31 \times 10^{-19}$ |
| Class years (Spring 2021) | 296.95 (5) | $4.54 \times 10^{-62}$ |
| Sports contact level | 36.59 (2) | $1.13 \times 10^{-8}$ |
| Sports location (indoor vs. outdoor) | 1.60 (1) | 0.21 |
| Sports season | 10.22 (2) | 0.006 |
| Sports teams | 75.08 (25) | $6.60 \times 10^{-7}$ |
| Residence halls | 31.30 (11) | $9.87 \times 10^{-4}$ |

**Appendix Table 5.**
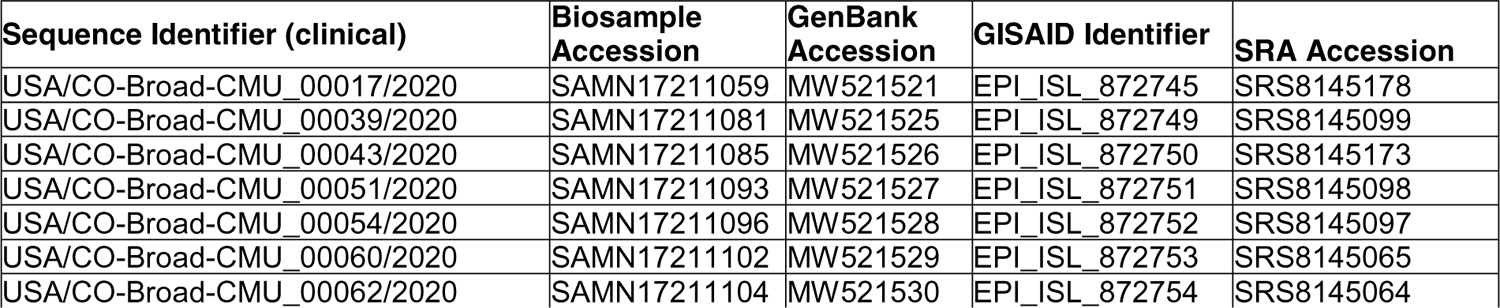
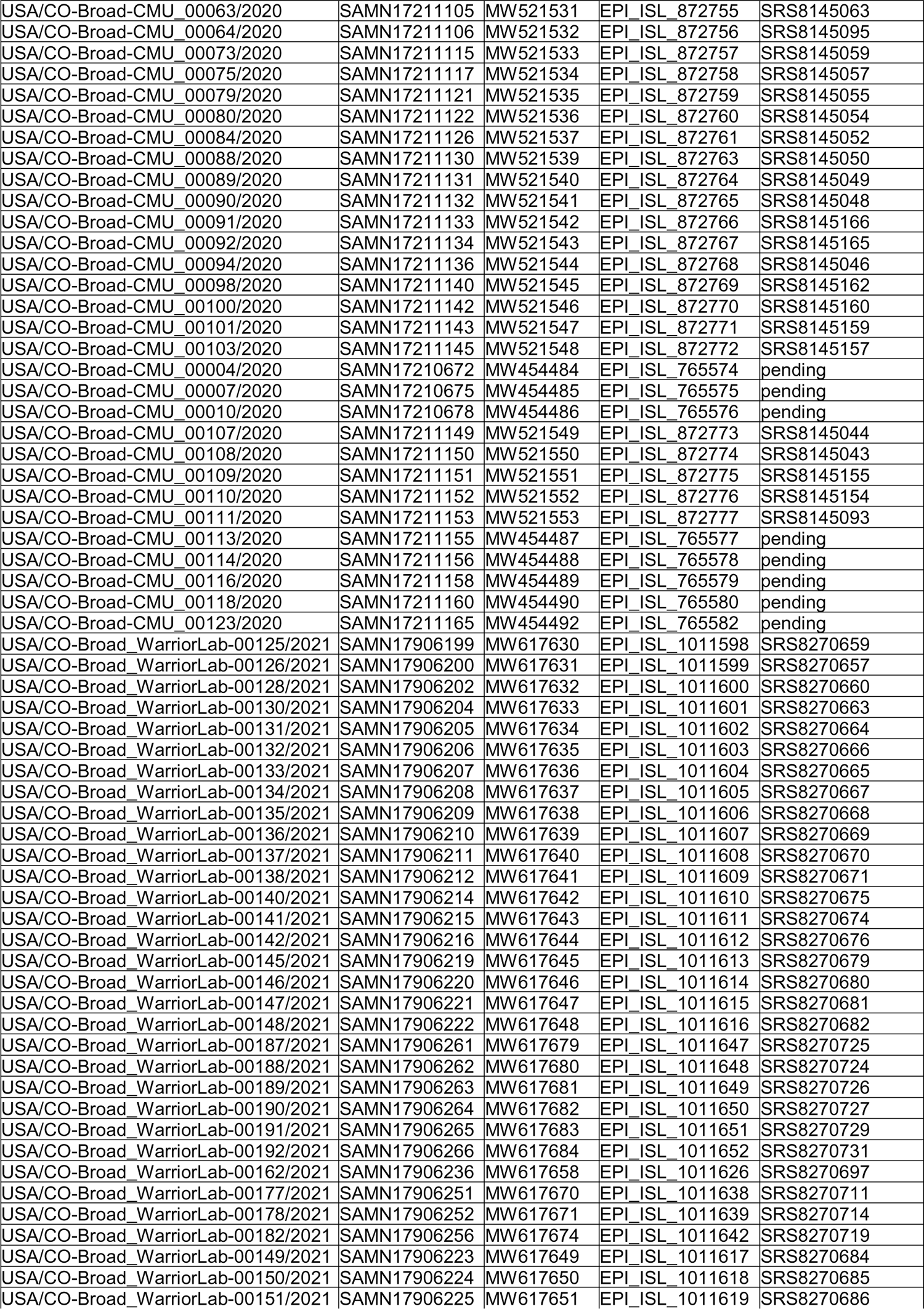
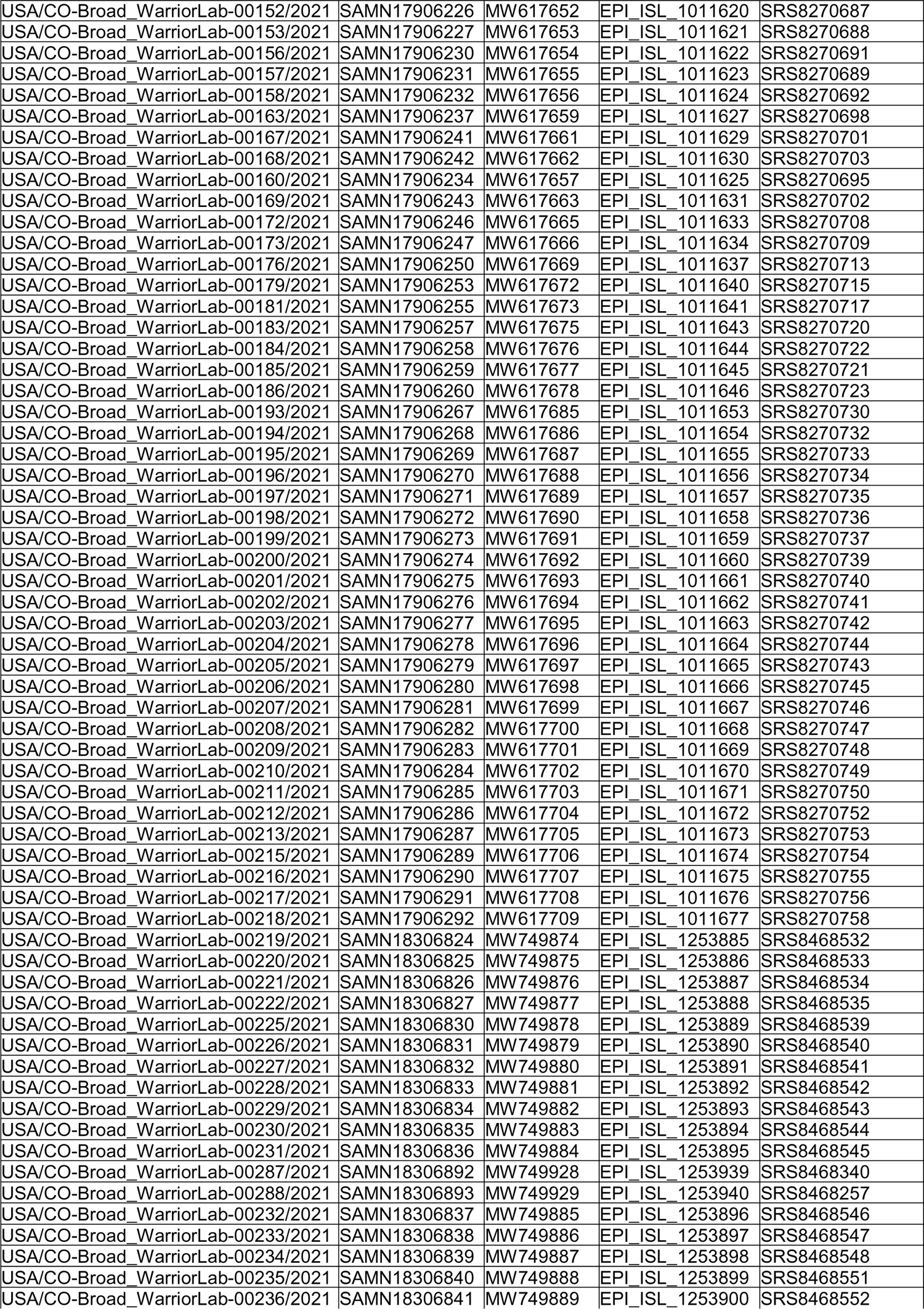
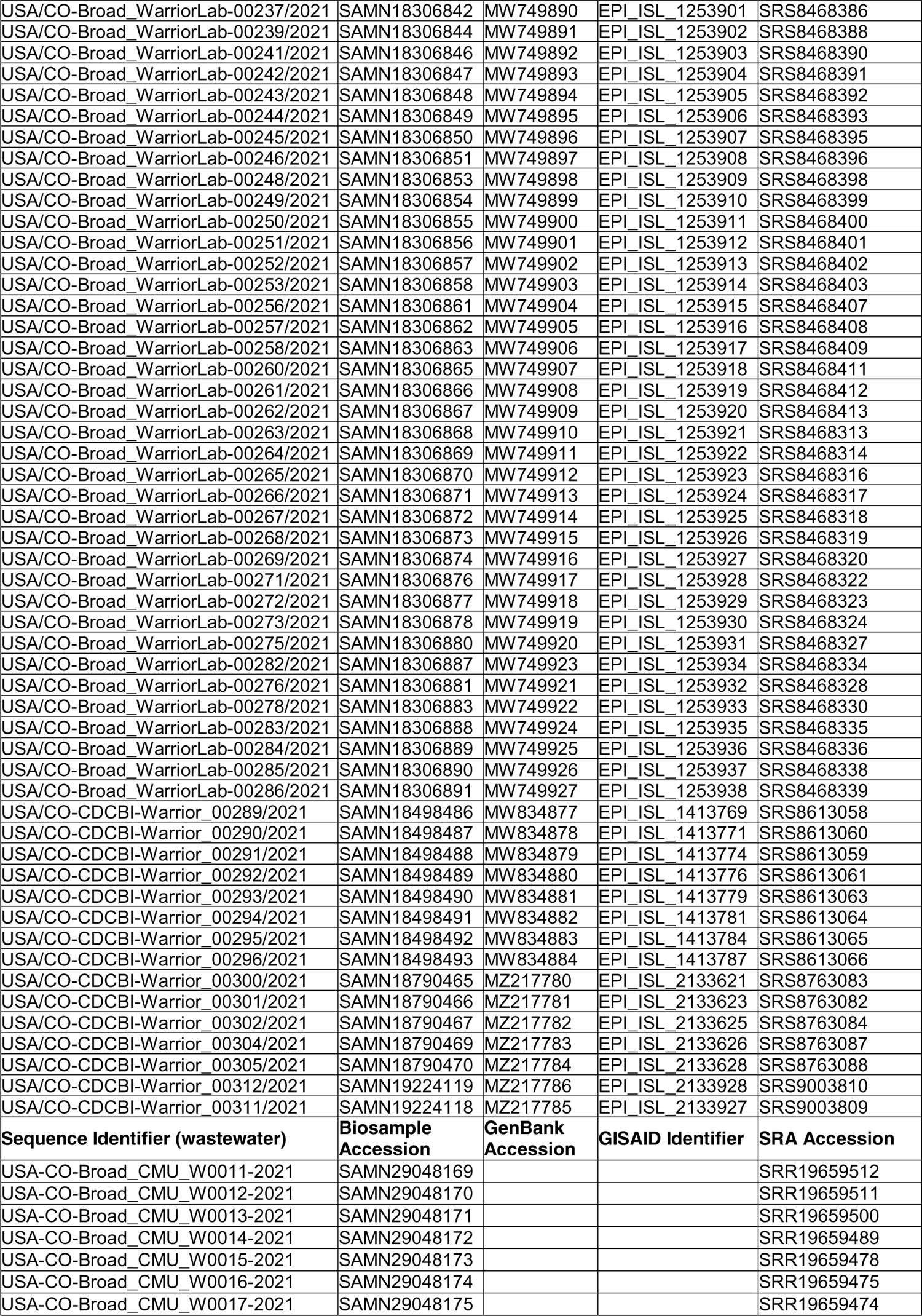
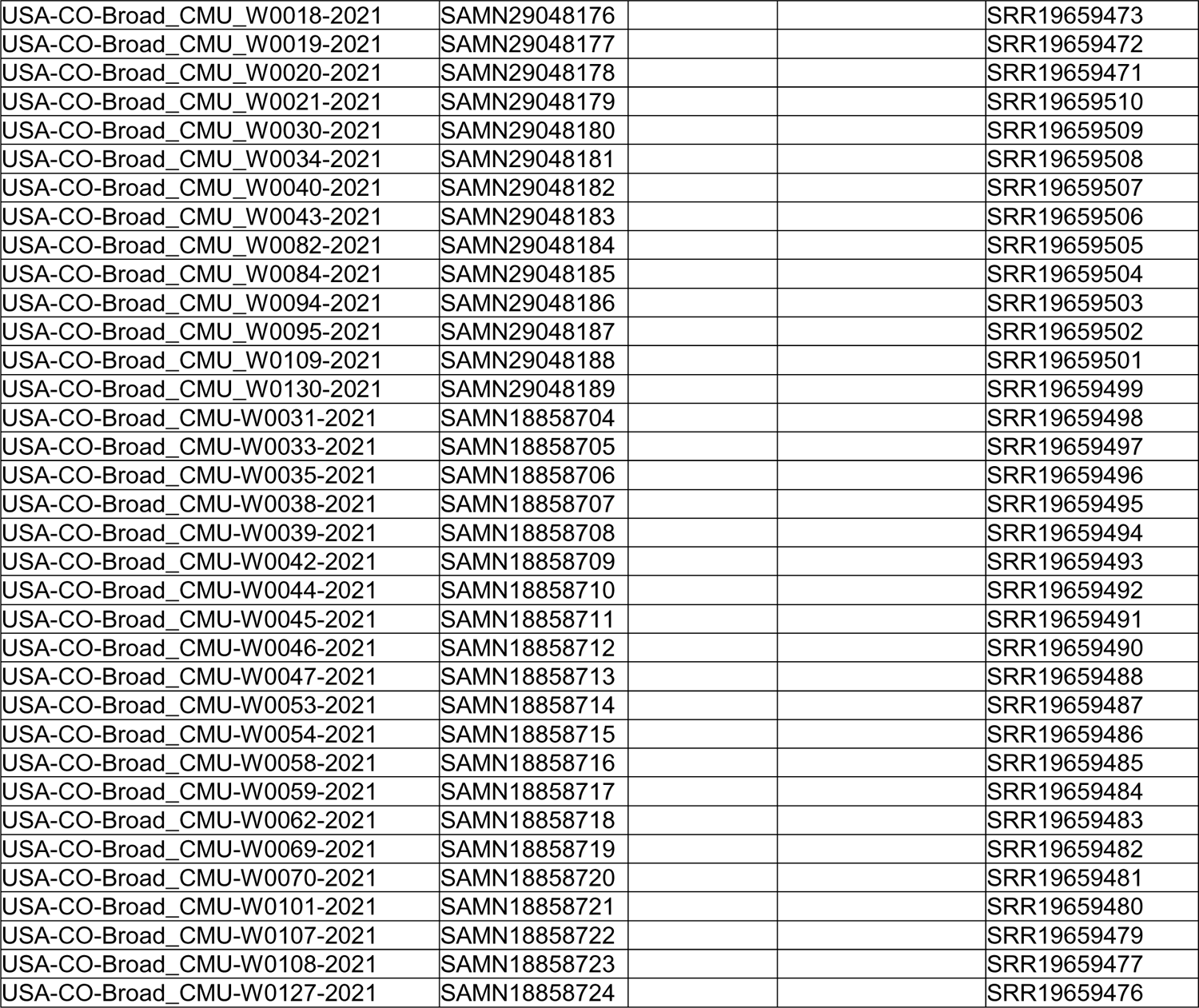
Published viral genomic data for clinical and environmental specimens Published sequences used in this study, listed with NCBI BioSample accessions, NCBI GenBank sequence accessions, GISAID identifiers, and/or SRA accessions.

| Sequence Identifier (clinical) | Biosample Accession | GenBank Accession | GISAID Identifier | SRA Accession |
| --- | --- | --- | --- | --- |
| USA/CO-Broad-CMU_00017/2020 | SAMN17211059 | MW521521 | EPI_ISL_872745 | SRS8145178 |
| USA/CO-Broad-CMU_00039/2020 | SAMN17211081 | MW521525 | EPI_ISL_872749 | SRS8145099 |
| USA/CO-Broad-CMU_00043/2020 | SAMN17211085 | MW521526 | EPI_ISL_872750 | SRS8145173 |
| USA/CO-Broad-CMU_00051/2020 | SAMN17211093 | MW521527 | EPI_ISL_872751 | SRS8145098 |
| USA/CO-Broad-CMU_00054/2020 | SAMN17211096 | MW521528 | EPI_ISL_872752 | SRS8145097 |
| USA/CO-Broad-CMU_00060/2020 | SAMN17211102 | MW521529 | EPI_ISL_872753 | SRS8145065 |
| USA/CO-Broad-CMU_00062/2020 | SAMN17211104 | MW521530 | EPI_ISL_872754 | SRS8145064 |
| USA/CO-Broad-CMU_00063/2020 | SAMN17211105 | MW521531 | EPI_ISL_872755 | SRS8145063 |
| USA/CO-Broad-CMU_00064/2020 | SAMN17211106 | MW521532 | EPI_ISL_872756 | SRS8145095 |
| USA/CO-Broad-CMU_00073/2020 | SAMN17211115 | MW521533 | EPI_ISL_872757 | SRS8145059 |
| USA/CO-Broad-CMU_00075/2020 | SAMN17211117 | MW521534 | EPI_ISL_872758 | SRS8145057 |
| USA/CO-Broad-CMU_00079/2020 | SAMN17211121 | MW521535 | EPI_ISL_872759 | SRS8145055 |
| USA/CO-Broad-CMU_00080/2020 | SAMN17211122 | MW521536 | EPI_ISL_872760 | SRS8145054 |
| USA/CO-Broad-CMU_00084/2020 | SAMN17211126 | MW521537 | EPI_ISL_872761 | SRS8145052 |
| USA/CO-Broad-CMU_00088/2020 | SAMN17211130 | MW521539 | EPI_ISL_872763 | SRS8145050 |
| USA/CO-Broad-CMU_00089/2020 | SAMN17211131 | MW521540 | EPI_ISL_872764 | SRS8145049 |
| USA/CO-Broad-CMU_00090/2020 | SAMN17211132 | MW521541 | EPI_ISL_872765 | SRS8145048 |
| USA/CO-Broad-CMU_00091/2020 | SAMN17211133 | MW521542 | EPI_ISL_872766 | SRS8145166 |
| USA/CO-Broad-CMU_00092/2020 | SAMN17211134 | MW521543 | EPI_ISL_872767 | SRS8145165 |
| USA/CO-Broad-CMU_00094/2020 | SAMN17211136 | MW521544 | EPI_ISL_872768 | SRS8145046 |
| USA/CO-Broad-CMU_00098/2020 | SAMN17211140 | MW521545 | EPI_ISL_872769 | SRS8145162 |
| USA/CO-Broad-CMU_00100/2020 | SAMN17211142 | MW521546 | EPI_ISL_872770 | SRS8145160 |
| USA/CO-Broad-CMU_00101/2020 | SAMN17211143 | MW521547 | EPI_ISL_872771 | SRS8145159 |
| USA/CO-Broad-CMU_00103/2020 | SAMN17211145 | MW521548 | EPI_ISL_872772 | SRS8145157 |
| USA/CO-Broad-CMU_00004/2020 | SAMN17210672 | MW454484 | EPI_ISL_765574 | pending |
| USA/CO-Broad-CMU_00007/2020 | SAMN17210675 | MW454485 | EPI_ISL_765575 | pending |
| USA/CO-Broad-CMU_00010/2020 | SAMN17210678 | MW454486 | EPI_ISL_765576 | pending |
| USA/CO-Broad-CMU_00107/2020 | SAMN17211149 | MW521549 | EPI_ISL_872773 | SRS8145044 |
| USA/CO-Broad-CMU_00108/2020 | SAMN17211150 | MW521550 | EPI_ISL_872774 | SRS8145043 |
| USA/CO-Broad-CMU_00109/2020 | SAMN17211151 | MW521551 | EPI_ISL_872775 | SRS8145155 |
| USA/CO-Broad-CMU_00110/2020 | SAMN17211152 | MW521552 | EPI_ISL_872776 | SRS8145154 |
| USA/CO-Broad-CMU_00111/2020 | SAMN17211153 | MW521553 | EPI_ISL_872777 | SRS8145093 |
| USA/CO-Broad-CMU_00113/2020 | SAMN17211155 | MW454487 | EPI_ISL_765577 | pending |
| USA/CO-Broad-CMU_00114/2020 | SAMN17211156 | MW454488 | EPI_ISL_765578 | pending |
| USA/CO-Broad-CMU_00116/2020 | SAMN17211158 | MW454489 | EPI_ISL_765579 | pending |
| USA/CO-Broad-CMU_00118/2020 | SAMN17211160 | MW454490 | EPI_ISL_765580 | pending |
| USA/CO-Broad-CMU_00123/2020 | SAMN17211165 | MW454492 | EPI_ISL_765582 | pending |
| USA/CO-Broad_WarriorLab-00125/2021 | SAMN17906199 | MW617630 | EPI_ISL_1011598 | SRS8270659 |
| USA/CO-Broad_WarriorLab-00126/2021 | SAMN17906200 | MW617631 | EPI_ISL_1011599 | SRS8270657 |
| USA/CO-Broad_WarriorLab-00128/2021 | SAMN17906202 | MW617632 | EPI_ISL_1011600 | SRS8270660 |
| USA/CO-Broad_WarriorLab-00130/2021 | SAMN17906204 | MW617633 | EPI_ISL_1011601 | SRS8270663 |
| USA/CO-Broad_WarriorLab-00131/2021 | SAMN17906205 | MW617634 | EPI_ISL_1011602 | SRS8270664 |
| USA/CO-Broad_WarriorLab-00132/2021 | SAMN17906206 | MW617635 | EPI_ISL_1011603 | SRS8270666 |
| USA/CO-Broad_WarriorLab-00133/2021 | SAMN17906207 | MW617636 | EPI_ISL_1011604 | SRS8270665 |
| USA/CO-Broad_WarriorLab-00134/2021 | SAMN17906208 | MW617637 | EPI_ISL_1011605 | SRS8270667 |
| USA/CO-Broad_WarriorLab-00135/2021 | SAMN17906209 | MW617638 | EPI_ISL_1011606 | SRS8270668 |
| USA/CO-Broad_WarriorLab-00136/2021 | SAMN17906210 | MW617639 | EPI_ISL_1011607 | SRS8270669 |
| USA/CO-Broad_WarriorLab-00137/2021 | SAMN17906211 | MW617640 | EPI_ISL_1011608 | SRS8270670 |
| USA/CO-Broad_WarriorLab-00138/2021 | SAMN17906212 | MW617641 | EPI_ISL_1011609 | SRS8270671 |
| USA/CO-Broad_WarriorLab-00140/2021 | SAMN17906214 | MW617642 | EPI_ISL_1011610 | SRS8270675 |
| USA/CO-Broad_WarriorLab-00141/2021 | SAMN17906215 | MW617643 | EPI_ISL_1011611 | SRS8270674 |
| USA/CO-Broad_WarriorLab-00142/2021 | SAMN17906216 | MW617644 | EPI_ISL_1011612 | SRS8270676 |
| USA/CO-Broad_WarriorLab-00145/2021 | SAMN17906219 | MW617645 | EPI_ISL_1011613 | SRS8270679 |
| USA/CO-Broad_WarriorLab-00146/2021 | SAMN17906220 | MW617646 | EPI_ISL_1011614 | SRS8270680 |
| USA/CO-Broad_WarriorLab-00147/2021 | SAMN17906221 | MW617647 | EPI_ISL_1011615 | SRS8270681 |
| USA/CO-Broad_WarriorLab-00148/2021 | SAMN17906222 | MW617648 | EPI_ISL_1011616 | SRS8270682 |
| USA/CO-Broad_WarriorLab-00187/2021 | SAMN17906261 | MW617679 | EPI_ISL_1011647 | SRS8270725 |
| USA/CO-Broad_WarriorLab-00188/2021 | SAMN17906262 | MW617680 | EPI_ISL_1011648 | SRS8270724 |
| USA/CO-Broad_WarriorLab-00189/2021 | SAMN17906263 | MW617681 | EPI_ISL_1011649 | SRS8270726 |
| USA/CO-Broad_WarriorLab-00190/2021 | SAMN17906264 | MW617682 | EPI_ISL_1011650 | SRS8270727 |
| USA/CO-Broad_WarriorLab-00191/2021 | SAMN17906265 | MW617683 | EPI_ISL_1011651 | SRS8270729 |
| USA/CO-Broad_WarriorLab-00192/2021 | SAMN17906266 | MW617684 | EPI_ISL_1011652 | SRS8270731 |
| USA/CO-Broad_WarriorLab-00162/2021 | SAMN17906236 | MW617658 | EPI_ISL_1011626 | SRS8270697 |
| USA/CO-Broad_WarriorLab-00177/2021 | SAMN17906251 | MW617670 | EPI_ISL_1011638 | SRS8270711 |
| USA/CO-Broad_WarriorLab-00178/2021 | SAMN17906252 | MW617671 | EPI_ISL_1011639 | SRS8270714 |
| USA/CO-Broad_WarriorLab-00182/2021 | SAMN17906256 | MW617674 | EPI_ISL_1011642 | SRS8270719 |
| USA/CO-Broad_WarriorLab-00149/2021 | SAMN17906223 | MW617649 | EPI_ISL_1011617 | SRS8270684 |
| USA/CO-Broad_WarriorLab-00150/2021 | SAMN17906224 | MW617650 | EPI_ISL_1011618 | SRS8270685 |
| USA/CO-Broad_WarriorLab-00151/2021 | SAMN17906225 | MW617651 | EPI_ISL_1011619 | SRS8270686 |
| USA/CO-Broad_WarriorLab-00152/2021 | SAMN17906226 | MW617652 | EPI_ISL_1011620 | SRS8270687 |
| USA/CO-Broad_WarriorLab-00153/2021 | SAMN17906227 | MW617653 | EPI_ISL_1011621 | SRS8270688 |
| USA/CO-Broad_WarriorLab-00156/2021 | SAMN17906230 | MW617654 | EPI_ISL_1011622 | SRS8270691 |
| USA/CO-Broad_WarriorLab-00157/2021 | SAMN17906231 | MW617655 | EPI_ISL_1011623 | SRS8270689 |
| USA/CO-Broad_WarriorLab-00158/2021 | SAMN17906232 | MW617656 | EPI_ISL_1011624 | SRS8270692 |
| USA/CO-Broad_WarriorLab-00163/2021 | SAMN17906237 | MW617659 | EPI_ISL_1011627 | SRS8270698 |
| USA/CO-Broad_WarriorLab-00167/2021 | SAMN17906241 | MW617661 | EPI_ISL_1011629 | SRS8270701 |
| USA/CO-Broad_WarriorLab-00168/2021 | SAMN17906242 | MW617662 | EPI_ISL_1011630 | SRS8270703 |
| USA/CO-Broad_WarriorLab-00160/2021 | SAMN17906234 | MW617657 | EPI_ISL_1011625 | SRS8270695 |
| USA/CO-Broad_WarriorLab-00169/2021 | SAMN17906243 | MW617663 | EPI_ISL_1011631 | SRS8270702 |
| USA/CO-Broad_WarriorLab-00172/2021 | SAMN17906246 | MW617665 | EPI_ISL_1011633 | SRS8270708 |
| USA/CO-Broad_WarriorLab-00173/2021 | SAMN17906247 | MW617666 | EPI_ISL_1011634 | SRS8270709 |
| USA/CO-Broad_WarriorLab-00176/2021 | SAMN17906250 | MW617669 | EPI_ISL_1011637 | SRS8270713 |
| USA/CO-Broad_WarriorLab-00179/2021 | SAMN17906253 | MW617672 | EPI_ISL_1011640 | SRS8270715 |
| USA/CO-Broad_WarriorLab-00181/2021 | SAMN17906255 | MW617673 | EPI_ISL_1011641 | SRS8270717 |
| USA/CO-Broad_WarriorLab-00183/2021 | SAMN17906257 | MW617675 | EPI_ISL_1011643 | SRS8270720 |
| USA/CO-Broad_WarriorLab-00184/2021 | SAMN17906258 | MW617676 | EPI_ISL_1011644 | SRS8270722 |
| USA/CO-Broad_WarriorLab-00185/2021 | SAMN17906259 | MW617677 | EPI_ISL_1011645 | SRS8270721 |
| USA/CO-Broad_WarriorLab-00186/2021 | SAMN17906260 | MW617678 | EPI_ISL_1011646 | SRS8270723 |
| USA/CO-Broad_WarriorLab-00193/2021 | SAMN17906267 | MW617685 | EPI_ISL_1011653 | SRS8270730 |
| USA/CO-Broad_WarriorLab-00194/2021 | SAMN17906268 | MW617686 | EPI_ISL_1011654 | SRS8270732 |
| USA/CO-Broad_WarriorLab-00195/2021 | SAMN17906269 | MW617687 | EPI_ISL_1011655 | SRS8270733 |
| USA/CO-Broad_WarriorLab-00196/2021 | SAMN17906270 | MW617688 | EPI_ISL_1011656 | SRS8270734 |
| USA/CO-Broad_WarriorLab-00197/2021 | SAMN17906271 | MW617689 | EPI_ISL_1011657 | SRS8270735 |
| USA/CO-Broad_WarriorLab-00198/2021 | SAMN17906272 | MW617690 | EPI_ISL_1011658 | SRS8270736 |
| USA/CO-Broad_WarriorLab-00199/2021 | SAMN17906273 | MW617691 | EPI_ISL_1011659 | SRS8270737 |
| USA/CO-Broad_WarriorLab-00200/2021 | SAMN17906274 | MW617692 | EPI_ISL_1011660 | SRS8270739 |
| USA/CO-Broad_WarriorLab-00201/2021 | SAMN17906275 | MW617693 | EPI_ISL_1011661 | SRS8270740 |
| USA/CO-Broad_WarriorLab-00202/2021 | SAMN17906276 | MW617694 | EPI_ISL_1011662 | SRS8270741 |
| USA/CO-Broad_WarriorLab-00203/2021 | SAMN17906277 | MW617695 | EPI_ISL_1011663 | SRS8270742 |
| USA/CO-Broad_WarriorLab-00204/2021 | SAMN17906278 | MW617696 | EPI_ISL_1011664 | SRS8270744 |
| USA/CO-Broad_WarriorLab-00205/2021 | SAMN17906279 | MW617697 | EPI_ISL_1011665 | SRS8270743 |
| USA/CO-Broad_WarriorLab-00206/2021 | SAMN17906280 | MW617698 | EPI_ISL_1011666 | SRS8270745 |
| USA/CO-Broad_WarriorLab-00207/2021 | SAMN17906281 | MW617699 | EPI_ISL_1011667 | SRS8270746 |
| USA/CO-Broad_WarriorLab-00208/2021 | SAMN17906282 | MW617700 | EPI_ISL_1011668 | SRS8270747 |
| USA/CO-Broad_WarriorLab-00209/2021 | SAMN17906283 | MW617701 | EPI_ISL_1011669 | SRS8270748 |
| USA/CO-Broad_WarriorLab-00210/2021 | SAMN17906284 | MW617702 | EPI_ISL_1011670 | SRS8270749 |
| USA/CO-Broad_WarriorLab-00211/2021 | SAMN17906285 | MW617703 | EPI_ISL_1011671 | SRS8270750 |
| USA/CO-Broad_WarriorLab-00212/2021 | SAMN17906286 | MW617704 | EPI_ISL_1011672 | SRS8270752 |
| USA/CO-Broad_WarriorLab-00213/2021 | SAMN17906287 | MW617705 | EPI_ISL_1011673 | SRS8270753 |
| USA/CO-Broad_WarriorLab-00215/2021 | SAMN17906289 | MW617706 | EPI_ISL_1011674 | SRS8270754 |
| USA/CO-Broad_WarriorLab-00216/2021 | SAMN17906290 | MW617707 | EPI_ISL_1011675 | SRS8270755 |
| USA/CO-Broad_WarriorLab-00217/2021 | SAMN17906291 | MW617708 | EPI_ISL_1011676 | SRS8270756 |
| USA/CO-Broad_WarriorLab-00218/2021 | SAMN17906292 | MW617709 | EPI_ISL_1011677 | SRS8270758 |
| USA/CO-Broad_WarriorLab-00219/2021 | SAMN18306824 | MW749874 | EPI_ISL_1253885 | SRS8468532 |
| USA/CO-Broad_WarriorLab-00220/2021 | SAMN18306825 | MW749875 | EPI_ISL_1253886 | SRS8468533 |
| USA/CO-Broad_WarriorLab-00221/2021 | SAMN18306826 | MW749876 | EPI_ISL_1253887 | SRS8468534 |
| USA/CO-Broad_WarriorLab-00222/2021 | SAMN18306827 | MW749877 | EPI_ISL_1253888 | SRS8468535 |
| USA/CO-Broad_WarriorLab-00225/2021 | SAMN18306830 | MW749878 | EPI_ISL_1253889 | SRS8468539 |
| USA/CO-Broad_WarriorLab-00226/2021 | SAMN18306831 | MW749879 | EPI_ISL_1253890 | SRS8468540 |
| USA/CO-Broad_WarriorLab-00227/2021 | SAMN18306832 | MW749880 | EPI_ISL_1253891 | SRS8468541 |
| USA/CO-Broad_WarriorLab-00228/2021 | SAMN18306833 | MW749881 | EPI_ISL_1253892 | SRS8468542 |
| USA/CO-Broad_WarriorLab-00229/2021 | SAMN18306834 | MW749882 | EPI_ISL_1253893 | SRS8468543 |
| USA/CO-Broad_WarriorLab-00230/2021 | SAMN18306835 | MW749883 | EPI_ISL_1253894 | SRS8468544 |
| USA/CO-Broad_WarriorLab-00231/2021 | SAMN18306836 | MW749884 | EPI_ISL_1253895 | SRS8468545 |
| USA/CO-Broad_WarriorLab-00287/2021 | SAMN18306892 | MW749928 | EPI_ISL_1253939 | SRS8468340 |
| USA/CO-Broad_WarriorLab-00288/2021 | SAMN18306893 | MW749929 | EPI_ISL_1253940 | SRS8468257 |
| USA/CO-Broad_WarriorLab-00232/2021 | SAMN18306837 | MW749885 | EPI_ISL_1253896 | SRS8468546 |
| USA/CO-Broad_WarriorLab-00233/2021 | SAMN18306838 | MW749886 | EPI_ISL_1253897 | SRS8468547 |
| USA/CO-Broad_WarriorLab-00234/2021 | SAMN18306839 | MW749887 | EPI_ISL_1253898 | SRS8468548 |
| USA/CO-Broad_WarriorLab-00235/2021 | SAMN18306840 | MW749888 | EPI_ISL_1253899 | SRS8468551 |
| USA/CO-Broad_WarriorLab-00236/2021 | SAMN18306841 | MW749889 | EPI_ISL_1253900 | SRS8468552 |
| USA/CO-Broad_WarriorLab-00237/2021 | SAMN18306842 | MW749890 | EPI_ISL_1253901 | SRS8468386 |
| USA/CO-Broad_WarriorLab-00239/2021 | SAMN18306844 | MW749891 | EPI_ISL_1253902 | SRS8468388 |
| USA/CO-Broad_WarriorLab-00241/2021 | SAMN18306846 | MW749892 | EPI_ISL_1253903 | SRS8468390 |
| USA/CO-Broad_WarriorLab-00242/2021 | SAMN18306847 | MW749893 | EPI_ISL_1253904 | SRS8468391 |
| USA/CO-Broad_WarriorLab-00243/2021 | SAMN18306848 | MW749894 | EPI_ISL_1253905 | SRS8468392 |
| USA/CO-Broad_WarriorLab-00244/2021 | SAMN18306849 | MW749895 | EPI_ISL_1253906 | SRS8468393 |
| USA/CO-Broad_WarriorLab-00245/2021 | SAMN18306850 | MW749896 | EPI_ISL_1253907 | SRS8468395 |
| USA/CO-Broad_WarriorLab-00246/2021 | SAMN18306851 | MW749897 | EPI_ISL_1253908 | SRS8468396 |
| USA/CO-Broad_WarriorLab-00248/2021 | SAMN18306853 | MW749898 | EPI_ISL_1253909 | SRS8468398 |
| USA/CO-Broad_WarriorLab-00249/2021 | SAMN18306854 | MW749899 | EPI_ISL_1253910 | SRS8468399 |
| USA/CO-Broad_WarriorLab-00250/2021 | SAMN18306855 | MW749900 | EPI_ISL_1253911 | SRS8468400 |
| USA/CO-Broad_WarriorLab-00251/2021 | SAMN18306856 | MW749901 | EPI_ISL_1253912 | SRS8468401 |
| USA/CO-Broad_WarriorLab-00252/2021 | SAMN18306857 | MW749902 | EPI_ISL_1253913 | SRS8468402 |
| USA/CO-Broad_WarriorLab-00253/2021 | SAMN18306858 | MW749903 | EPI_ISL_1253914 | SRS8468403 |
| USA/CO-Broad_WarriorLab-00256/2021 | SAMN18306861 | MW749904 | EPI_ISL_1253915 | SRS8468407 |
| USA/CO-Broad_WarriorLab-00257/2021 | SAMN18306862 | MW749905 | EPI_ISL_1253916 | SRS8468408 |
| USA/CO-Broad_WarriorLab-00258/2021 | SAMN18306863 | MW749906 | EPI_ISL_1253917 | SRS8468409 |
| USA/CO-Broad_WarriorLab-00260/2021 | SAMN18306865 | MW749907 | EPI_ISL_1253918 | SRS8468411 |
| USA/CO-Broad_WarriorLab-00261/2021 | SAMN18306866 | MW749908 | EPI_ISL_1253919 | SRS8468412 |
| USA/CO-Broad_WarriorLab-00262/2021 | SAMN18306867 | MW749909 | EPI_ISL_1253920 | SRS8468413 |
| USA/CO-Broad_WarriorLab-00263/2021 | SAMN18306868 | MW749910 | EPI_ISL_1253921 | SRS8468313 |
| USA/CO-Broad_WarriorLab-00264/2021 | SAMN18306869 | MW749911 | EPI_ISL_1253922 | SRS8468314 |
| USA/CO-Broad_WarriorLab-00265/2021 | SAMN18306870 | MW749912 | EPI_ISL_1253923 | SRS8468316 |
| USA/CO-Broad_WarriorLab-00266/2021 | SAMN18306871 | MW749913 | EPI_ISL_1253924 | SRS8468317 |
| USA/CO-Broad_WarriorLab-00267/2021 | SAMN18306872 | MW749914 | EPI_ISL_1253925 | SRS8468318 |
| USA/CO-Broad_WarriorLab-00268/2021 | SAMN18306873 | MW749915 | EPI_ISL_1253926 | SRS8468319 |
| USA/CO-Broad_WarriorLab-00269/2021 | SAMN18306874 | MW749916 | EPI_ISL_1253927 | SRS8468320 |
| USA/CO-Broad_WarriorLab-00271/2021 | SAMN18306876 | MW749917 | EPI_ISL_1253928 | SRS8468322 |
| USA/CO-Broad_WarriorLab-00272/2021 | SAMN18306877 | MW749918 | EPI_ISL_1253929 | SRS8468323 |
| USA/CO-Broad_WarriorLab-00273/2021 | SAMN18306878 | MW749919 | EPI_ISL_1253930 | SRS8468324 |
| USA/CO-Broad_WarriorLab-00275/2021 | SAMN18306880 | MW749920 | EPI_ISL_1253931 | SRS8468327 |
| USA/CO-Broad_WarriorLab-00282/2021 | SAMN18306887 | MW749923 | EPI_ISL_1253934 | SRS8468334 |
| USA/CO-Broad_WarriorLab-00276/2021 | SAMN18306881 | MW749921 | EPI_ISL_1253932 | SRS8468328 |
| USA/CO-Broad_WarriorLab-00278/2021 | SAMN18306883 | MW749922 | EPI_ISL_1253933 | SRS8468330 |
| USA/CO-Broad_WarriorLab-00283/2021 | SAMN18306888 | MW749924 | EPI_ISL_1253935 | SRS8468335 |
| USA/CO-Broad_WarriorLab-00284/2021 | SAMN18306889 | MW749925 | EPI_ISL_1253936 | SRS8468336 |
| USA/CO-Broad_WarriorLab-00285/2021 | SAMN18306890 | MW749926 | EPI_ISL_1253937 | SRS8468338 |
| USA/CO-Broad_WarriorLab-00286/2021 | SAMN18306891 | MW749927 | EPI_ISL_1253938 | SRS8468339 |
| USA/CO-CDCBI-Warrior_00289/2021 | SAMN18498486 | MW834877 | EPI_ISL_1413769 | SRS8613058 |
| USA/CO-CDCBI-Warrior_00290/2021 | SAMN18498487 | MW834878 | EPI_ISL_1413771 | SRS8613060 |
| USA/CO-CDCBI-Warrior_00291/2021 | SAMN18498488 | MW834879 | EPI_ISL_1413774 | SRS8613059 |
| USA/CO-CDCBI-Warrior_00292/2021 | SAMN18498489 | MW834880 | EPI_ISL_1413776 | SRS8613061 |
| USA/CO-CDCBI-Warrior_00293/2021 | SAMN18498490 | MW834881 | EPI_ISL_1413779 | SRS8613063 |
| USA/CO-CDCBI-Warrior_00294/2021 | SAMN18498491 | MW834882 | EPI_ISL_1413781 | SRS8613064 |
| USA/CO-CDCBI-Warrior_00295/2021 | SAMN18498492 | MW834883 | EPI_ISL_1413784 | SRS8613065 |
| USA/CO-CDCBI-Warrior_00296/2021 | SAMN18498493 | MW834884 | EPI_ISL_1413787 | SRS8613066 |
| USA/CO-CDCBI-Warrior_00300/2021 | SAMN18790465 | MZ217780 | EPI_ISL_2133621 | SRS8763083 |
| USA/CO-CDCBI-Warrior_00301/2021 | SAMN18790466 | MZ217781 | EPI_ISL_2133623 | SRS8763082 |
| USA/CO-CDCBI-Warrior_00302/2021 | SAMN18790467 | MZ217782 | EPI_ISL_2133625 | SRS8763084 |
| USA/CO-CDCBI-Warrior_00304/2021 | SAMN18790469 | MZ217783 | EPI_ISL_2133626 | SRS8763087 |
| USA/CO-CDCBI-Warrior_00305/2021 | SAMN18790470 | MZ217784 | EPI_ISL_2133628 | SRS8763088 |
| USA/CO-CDCBI-Warrior_00312/2021 | SAMN19224119 | MZ217786 | EPI_ISL_2133928 | SRS9003810 |
| USA/CO-CDCBI-Warrior_00311/2021 | SAMN19224118 | MZ217785 | EPI_ISL_2133927 | SRS9003809 |
| **Sequence Identifier (wastewater)** | **Biosample Accession** | **GenBank Accession** | **GISAID Identifier** | **SRA Accession** |
| USA-CO-Broad_CMU_W0011-2021 | SAMN29048169 |  |  | SRR19659512 |
| USA-CO-Broad_CMU_W0012-2021 | SAMN29048170 |  |  | SRR19659511 |
| USA-CO-Broad_CMU_W0013-2021 | SAMN29048171 |  |  | SRR19659500 |
| USA-CO-Broad_CMU_W0014-2021 | SAMN29048172 |  |  | SRR19659489 |
| USA-CO-Broad_CMU_W0015-2021 | SAMN29048173 |  |  | SRR19659478 |
| USA-CO-Broad_CMU_W0016-2021 | SAMN29048174 |  |  | SRR19659475 |
| USA-CO-Broad_CMU_W0017-2021 | SAMN29048175 |  |  | SRR19659474 |

|  |  |  |  |
| --- | --- | --- | --- |
| USA-CO-Broad_CMU_W0018-2021 | SAMN29048176 |  | SRR19659473 |
| USA-CO-Broad_CMU_W0019-2021 | SAMN29048177 |  | SRR19659472 |
| USA-CO-Broad_CMU_W0020-2021 | SAMN29048178 |  | SRR19659471 |
| USA-CO-Broad_CMU_W0021-2021 | SAMN29048179 |  | SRR19659510 |
| USA-CO-Broad_CMU_W0030-2021 | SAMN29048180 |  | SRR19659509 |
| USA-CO-Broad_CMU_W0034-2021 | SAMN29048181 |  | SRR19659508 |
| USA-CO-Broad_CMU_W0040-2021 | SAMN29048182 |  | SRR19659507 |
| USA-CO-Broad_CMU_W0043-2021 | SAMN29048183 |  | SRR19659506 |
| USA-CO-Broad_CMU_W0082-2021 | SAMN29048184 |  | SRR19659505 |
| USA-CO-Broad_CMU_W0084-2021 | SAMN29048185 |  | SRR19659504 |
| USA-CO-Broad_CMU_W0094-2021 | SAMN29048186 |  | SRR19659503 |
| USA-CO-Broad_CMU_W0095-2021 | SAMN29048187 |  | SRR19659502 |
| USA-CO-Broad_CMU_W0109-2021 | SAMN29048188 |  | SRR19659501 |
| USA-CO-Broad_CMU_W0130-2021 | SAMN29048189 |  | SRR19659499 |
| USA-CO-Broad_CMU-W0031-2021 | SAMN18858704 |  | SRR19659498 |
| USA-CO-Broad_CMU-W0033-2021 | SAMN18858705 |  | SRR19659497 |
| USA-CO-Broad_CMU-W0035-2021 | SAMN18858706 |  | SRR19659496 |
| USA-CO-Broad_CMU-W0038-2021 | SAMN18858707 |  | SRR19659495 |
| USA-CO-Broad_CMU-W0039-2021 | SAMN18858708 |  | SRR19659494 |
| USA-CO-Broad_CMU-W0042-2021 | SAMN18858709 |  | SRR19659493 |
| USA-CO-Broad_CMU-W0044-2021 | SAMN18858710 |  | SRR19659492 |
| USA-CO-Broad_CMU-W0045-2021 | SAMN18858711 |  | SRR19659491 |
| USA-CO-Broad_CMU-W0046-2021 | SAMN18858712 |  | SRR19659490 |
| USA-CO-Broad_CMU-W0047-2021 | SAMN18858713 |  | SRR19659488 |
| USA-CO-Broad_CMU-W0053-2021 | SAMN18858714 |  | SRR19659487 |
| USA-CO-Broad_CMU-W0054-2021 | SAMN18858715 |  | SRR19659486 |
| USA-CO-Broad_CMU-W0058-2021 | SAMN18858716 |  | SRR19659485 |
| USA-CO-Broad_CMU-W0059-2021 | SAMN18858717 |  | SRR19659484 |
| USA-CO-Broad_CMU-W0062-2021 | SAMN18858718 |  | SRR19659483 |
| USA-CO-Broad_CMU-W0069-2021 | SAMN18858719 |  | SRR19659482 |
| USA-CO-Broad_CMU-W0070-2021 | SAMN18858720 |  | SRR19659481 |
| USA-CO-Broad_CMU-W0101-2021 | SAMN18858721 |  | SRR19659480 |
| USA-CO-Broad_CMU-W0107-2021 | SAMN18858722 |  | SRR19659479 |
| USA-CO-Broad_CMU-W0108-2021 | SAMN18858723 |  | SRR19659477 |
| USA-CO-Broad_CMU-W0127-2021 | SAMN18858724 |  | SRR19659476 |

**Appendix Table 6.** Lineages present in sequenced wastewater samples Wastewater-detected lineages and their relative abundances. For each wastewater sample, collection date and site of collection are also listed. Lineages and abundances were determined via application of the Freyja program, as described in detail in the Supplementary Methods.

| Sample ID | Date | Site | B.1.1.519 | B.1.126 | B.1.2 | B.1.234 | B.1.350 | B.1.429 | B.1.429.1 | B.1.533 |
| --- | --- | --- | --- | --- | --- | --- | --- | --- | --- | --- |
| CO_CMU_W0011 | 2-12 | Site 5 |  |  | 0.09 |  |  |  | 0.84 |  |
| CO_CMU_W0012 | 2-12 | Site 5 |  |  | 0.11 |  |  |  | 0.84 |  |
| CO_CMU_W0013 | 2-12 | Site 5 |  |  | 0.12 |  |  |  | 0.82 |  |
| CO_CMU_W0014 | 2-16 | Site 1 |  | 1.00 |  |  |  |  |  |  |
| CO_CMU_W0015 | 2-16 | Site 8 |  |  | 0.12 |  |  | 0.11 | 0.77 |  |
| CO_CMU_W0016 | 2-17 | Site 3 |  |  |  |  |  |  | 1.00 |  |
| CO_CMU_W0017 | 2-17 | Site 6 |  |  | 0.91 |  |  |  |  |  |
| CO_CMU_W0018 | 2-17 | Site 7 |  |  | 1.00 |  |  |  |  |  |
| CO_CMU_W0019 | 2-17 | Site 11 |  |  |  |  |  |  |  |  |
| CO_CMU_W0020 | 2-17 | Site 4 |  |  |  |  |  |  | 1.00 |  |
| CO_CMU_W0021 | 2-17 | Site 2 |  |  | 0.93 |  |  |  |  |  |
| CO_CMU_W0030 | 2-11 | Site 6 |  |  | 0.13 | 0.31 |  |  | 0.54 |  |
| CO_CMU_W0034 | 2-15 | Site 5 |  |  | 0.28 |  |  |  | 0.47 |  |
| CO_CMU_W0040 | 2-18 | Site 11 |  |  | 0.79 |  |  |  |  |  |
| CO_CMU_W0043 | 2-18 | Site 8 |  |  |  |  |  | 0.16 | 0.81 |  |
| CO_CMU_W0082 | 3-01 | Site 6 |  |  |  |  |  |  | 0.93 |  |
| CO_CMU_W0084 | 3-01 | Site 4 |  |  | 0.99 |  |  |  |  |  |
| CO_CMU_W0094 | 3-04 | Site 5 |  |  | 0.58 |  |  |  | 0.34 |  |
| CO_CMU_W0095 | 3-04 | Site 3 |  |  | 0.49 |  |  |  | 0.26 |  |
| CO_CMU_W0109 | 3-09 | Site 1 |  |  |  |  |  |  | 1.00 |  |
| CO_CMU_W0130 | 3-18 | Site 1 |  |  |  |  |  |  | 0.31 |  |
| CO_CMU_W0031 | 2-11 | Site 1 |  |  |  |  | 0.03 |  | 0.96 |  |
| CO_CMU_W0033 | 2-15 | Site 10 |  |  |  |  |  |  | 0.99 |  |
| CO_CMU_W0035 | 2-18 | Site 5 |  |  | 0.99 |  |  |  |  |  |
| CO_CMU_W0038 | 2-18 | Site 4 |  |  | 0.99 |  |  |  |  |  |
| CO_CMU_W0039 | 2-18 | Site 1 |  |  | 0.93 |  |  |  | 0.05 |  |
| CO_CMU_W0042 | 2-18 | Site 10 |  |  |  |  |  |  | 0.99 |  |
| CO_CMU_W0046 | 2-22 | Site 4 |  |  | 0.99 |  |  |  |  |  |
| CO_CMU_W0058 | 2-23 | Site 11 |  |  | 0.98 |  |  |  |  |  |
| CO_CMU_W0059 | 2-23 | Site 5 |  |  |  |  |  |  | 0.85 |  |
| CO_CMU_W0062 | 2-23 | Site 1 |  |  |  |  |  |  | 1.00 |  |
| CO_CMU_W0069 | 2-25 | Site 5 |  |  | 0.99 |  |  |  |  |  |
| CO_CMU_W0070 | 2-25 | Site 3 |  |  |  |  |  |  | 0.96 |  |
| CO_CMU_W0101 | 3-08 | Site 3 |  |  |  |  |  |  | 0.99 |  |
| CO_CMU_W0107 | 3-08 | Site 5 |  |  |  |  |  |  | 0.94 |  |
| CO_CMU_W0108 | 3-09 | Site 3 |  |  |  |  |  |  | 0.99 |  |
| CO_CMU_W0127 | 3-16 | Site 5 | 0.12 |  | 0.08 |  |  |  | 0.72 | 0.07 |

**Appendix Table 7.** Replicate-confirmed single nucleotide variant mutations identified in sequenced wastewater samples but not present in sequenced clinical samples Replicate-confirmed mutations identified in wastewater samples from CMU, but not in clinical samples. SNV nucleotide changes and positions are listed in bp, relative to the reference ancestral genome, NC_045512.2 (1st column) along with corresponding amino acid changes (4th and 5th columns), the class of change (3rd column), the proportion of samples bearing the change (6th column), and the highest allele frequency of that change amongst those samples (final column). Some of these mutations were present in Colorado clinical viral genomes (2nd column), or representative of specific Pango lineages (penultimate column).

| SNV | In CO | Change | Gene | Amino acid | Presence in proportion of 42 wastewater samples | Maximum allele frequency |
| --- | --- | --- | --- | --- | --- | --- |
| T5260A | False | Synonymous | ORF1ab | T1665T | 10% | 29% |
| T6174G | False | Missense | ORF1ab | I1970S | 17% | 32% |
| T9843A | False | Stop | ORF1ab | L3193* | 14% | 4% |
| C10650T | False | Missense | ORF1ab | T3462I | 31% | 100% |
| T27444G | False | Synonymous | ORF7a | L17L | 7% | 27% |
| C835T | True | Synonymous | ORF1ab | F190F | 38% | 100% |
| C6501T | True | Missense | ORF1ab | P2079L | 38% | 99% |
| G14126A | True | Missense | ORF1ab | S4621N | 5% | 100% |
| C21952T | True | Synonymous | S | V130V | 38% | 100% |
| T26099C | True | Missense | ORF3a | I236T | 33% | 100% |
| C28344T | True | Missense | N | T24I | 38% | 96% |

**Appendix Table 8.** Nucleotide and amino acid substitutions present in the B.1.429.1 lineage Characteristic amino acid mutations for B.1.429.1 and parent lineage B.1.429. Mutations are categorized by location / gene. Mutations with an accompanying asterisk were not seen in all B.1.429.1 sequenced genomes; however, this may have been due to gaps in sequencing coverage.

| Region | Amino Acid Changes Characteristic of B.1.429 | Additional Amino Acid Changes Characteristic of B.1.429.1 |
| --- | --- | --- |
| ORF1a | T265I, I4205V | F2827L, V3367I |
| ORF1b | P314L, D1183Y | P314L, D1183Y |
| S | S13I, W152C*, L452R*, D614G* | Q677H |
| ORF3a | Q57H | A23V |
| N | T205I* | P142S, M234I* |
| ORF8 |  | V100L |

**Appendix Table 9.** Summary of data sources used at Colorado Mesa University for their infectious disease surveillance program and additional recommendations for future institutional surveillance programs. Comparison of the utility and costs of each data source utilized in this study. We recommend incorporation of these tools in a specific manner given resource availability (Figure 7). VoC = Variant of Concern; VOI = Variant of Interest.

|  | **Epidemiological Analyses** | **Clinical Viral Genomic Sequencing** | **Wastewater Surveillance & Sequencing** | **Wifi Proximity Analyses** |
| --- | --- | --- | --- | --- |
| **General Utility** | Identify epidemiological risk factors | Parametrize cluster distribution and identify lineages and mutations | Establish quality control standards | Discriminate interaction patterns by test positivity |
| **Current Utility to Institution** | Specify athletic, residential, and class-year risks | Identify VoC/VoI & transmission clusters | Designate testing resources based on surveillance; sequencing to detect mutations not seen clinically | _____ |
| **Potential for Future Utility** | Automatically integrate metadata, wastewater surveillance, sequencing, and wifi-based contact tracing into comprehensive surveillance system made directly accessible for the community or public health officials |  |  |  |
| **Cost of Acquisition** | *Medium:*<br>Personnel (cataloging information) | *Medium:*<br>Transportation of excess clinical samples | *Medium–High:*<br>Establishment of collection devices | *Low:*<br>Established infrastructure |
| **Cost of Analysis** | *Low:* Personnel (analysis) | *High:*<br>Sequencing reagents and computing resources | *Low:* PCR reagents | *Low:*<br>Computing resources |
|  |  |  | *Medium:*<br>Sequencing costs reduced relative to clinical due to fewer samples |  |
| **Difficulty of Analysis** | *Low:*<br>Established statistical methods | *Low:*<br>Established tools for phylogenetic trees | *Low:*<br>Established tools for PCR analysis | *High:*<br>Few established tools |
|  |  |  | *Medium:*<br>Few established tools/methods<br>for mixed sequence analysis |  |

**Appendix Table 10.** Jaccard similarity between clusters derived from genomic reconstruction supplemented by different contact tracing data sets We computed the Jaccard similarity for distinct transmission reconstruction networks for individuals with the B.1.429.1 lineage. We supplemented the genomic reconstruction in the trials using three different definitions of close contacts: (1) manual contact tracing data, (2) 10-day wifi-derived contacts, and (3) 2-day wifi-derived contacts.

| Comparison Groups | Cluster Color | Jaccard Similarity |
| --- | --- | --- |
| CT vs. 2-Day Wifi | Orange | .421 |
|  | Light Blue | 1.0 |
|  | Pink | 0.0 |
|  | Yellow | 1.0 |
| CT vs. 10-Day Wifi | Orange | .889 |
|  | Light Blue | 1.0 |
|  | Pink | 1.0 |
|  | Yellow | 1.0 |
| 2-Day Wifi vs. 10-Day Wifi | Orange | .474 |
|  | Light Blue | 1.0 |
|  | Pink | 0 |
|  | Yellow | 1.0 |

**Appendix Table 11.**
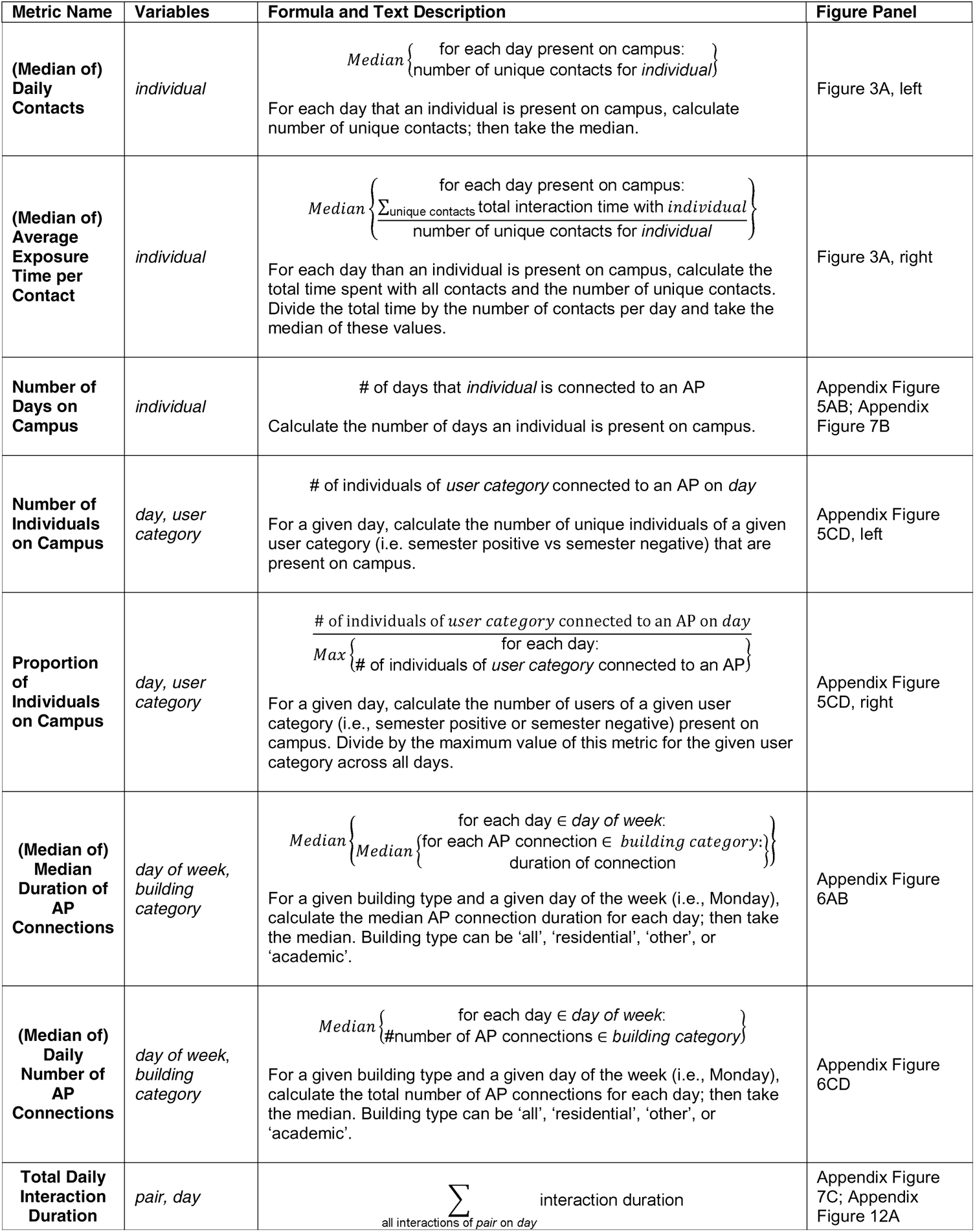
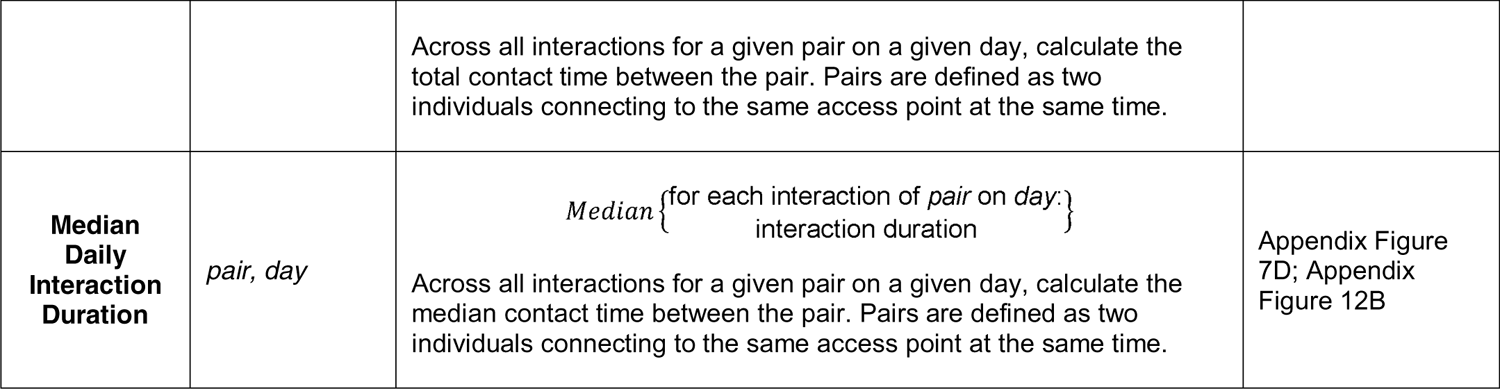
Summary of metrics used for the wifi proximity network analyses The metric names, associated variables, formulas, text descriptions, and the associated figure panel are listed for all metrics used in wifi proximity network analyses.

