## Supplemental Methods for "Multimodal surveillance of SARS-CoV-2 at a university enables development of a robust outbreak response framework"

#### **Epidemiological modeling**

##### 1. Relative risk analysis

For each risk factor of interest (Appendix Figure 1C), we calculated the relative risk and its confidence interval. The relative risk is defined as:

$\frac{\left( \frac{cases with risk factor}{students with risk factor} \right)}{(\frac{cases without risk factor}{students without risk factor})}$.

The natural logarithm of relative risk is approximately normally distributed with squared standard error defined as:

$\frac{\frac{students with risk factor-cases with risk factor}{cases with risk factor}}{students with risk factor}$+ $\frac{\frac{students without risk factor-cases without risk factor}{cases without risk factor}}{students without risk factor}$,

enabling calculation of 95% confidence intervals^1^.

Each risk factor was studied individually; we could not assess the relationship between factors (*e.g.*, whether the increased risk for males is explained by the increased risk for athletes) as we do not have information on the number of individuals at the intersection of risk factors.

##### 2. Chi-squared analysis

For categorical variables with multiple levels (*i.e.*, sports teams, sports contact levels, sports locations, residence halls, and class years), we assessed whether SARS-CoV-2 cases were distributed uniformly across levels. Specifically, we conducted chi-square goodness-of-fit tests where the expected number of cases in level *i* was:

$$expected_{i}=sum_{j}\left( cases in category j \right)* \frac{students in category i}{sum_{j}(students in category j)}$$

P-values were calculated using the chi-square distribution, with degrees of freedom equal to one less than the number of levels (Appendix Table 4).

##### 3. Regression model

We constructed a linear model of COVID-19 incidence rates (*i.e.*, case counts / number of residents) in a residence hall as a function of: number of students, percent occupied (number of available beds / number of students), number of floors, the presence of a meal plan requirement (‘dining hall’), the presence of an in-unit bathroom (‘private bath’), the number of resident advisors (RAs), square footage, ceiling height, volume per person (*i.e.*, a proxy for air volume: floor area * ceiling height / number of residents), and the presence of Fall wastewater surveillance (*n.b.*: wastewater surveillance covered all residence halls in the Spring; using Fall surveillance as an indicator variable allowed us to assess for any relationship between reflexive testing due to a positive wastewater titer and incidence rates). We assessed for multicollinearity among our predictive variables and identified many correlations (Appendix Figure 4A). Because of these relationships, our coefficients and their confidence intervals may not be robust; however, the model’s predictive power and R^2^ remain unaffected.

We evaluated all 1023 possible models, using all possible combinations of predictors (2^10^ - 1 combinations). For each model, we calculated the AIC and the BIC and selected the model with the lowest AIC (-41.95) and BIC (-40.98). The model, with an adjusted R^2^ of 0.95 (Appendix Figure 4B) is as follows:

$$incidence_{i}=\beta_{1}*percent\_occupancy_{i}+\beta_{2}*privat{e\_bathroom}_{i}$$

$\beta_{1}$ = 0.0015, $\beta_{2}$ = 0.0587

We evaluated the model *via* examination of the residual plot for heteroscedasticity (Appendix Figure 4C) and *via* a leave-one-out cross-validation analysis, *i.e.*, we fit the model using (*N*-1) data points and calculated the residual for the remaining data point to determine the mean squared error (Appendix Figure 4DE).

##### 4. Time series data

We used 2019 US Census Bureau data to determine the population sizes of Mesa County and Colorado. Using the United States county-level COVID-19 data, we determined a per-day incidence of COVID-19 cases and deaths for Mesa County and for all of Colorado. We also determined a per-day incidence rate for COVID-19 cases at CMU. We plotted rolling sums of 7 daily COVID-19 incidence rates to determine weekly incidence rates (Figure 2C; Appendix Figure 1A). We calculated Pearson correlation coefficients between CMU’s and Mesa County’s weekly incidence rates using the numpy.corrcoef function.

We determined test positivity rates per semester by dividing the total number of cases by the total number of tests.

##### 5. Contact tracing analyses

A positive individual’s close contacts were defined as individuals who were within 6 feet of each other for 15 minutes or more, in the 48-hour time period prior to symptom onset or test date, regardless of whether masks were worn. Each individual who tested positive was asked to report their close contacts. This information was collated into the following format: for each positive individual, we received the number of contacts they reported, as well as a barcode ID to identify any reported contacts who tested positive at any point throughout the academic year. From this dataset, we calculated the number of individuals who tested positive within 1-7 days of being reported as a close contact of a positive individual. An upper limit of 7 days was chosen to account for potential latencies in test result reporting, notification of contacts, and subsequent testing.

For all reported pairs of two positive individuals where one was identified as a close contact of the other and where both had sequenced viral samples, we calculated genomic distance (i.e. the number of single-nucleotide mutations differing between their consensus-level viral genomes, Appendix Figure 2A).

#### Wifi analysis

##### 1. **Data acquisition and cleaning**

Our partners at Degree Analytics, a behavioral analytics company, collected wifi connectivity data for Colorado Mesa University from August 2020 through May 2021. Degree Analytics collected the date, starting time, and duration of a specific device’s connectivity to wifi access points (APs) that were distributed across all university-affiliated spaces, including academic buildings, residence halls, dining locations, administrative buildings, and athletic spaces. Degree Analytics then ran device-specific data through a proprietary algorithm to produce pairwise interactions, defined as when two individuals simultaneously access the same (one or more) APs for at least 15 minutes, and unpaired connections, where one individual connects to (one or more) APs for at least 15 minutes with no simultaneous connections. The proprietary algorithm accounts for some level of uncertainty; for example, if a device were to disconnect (*e.g.*, because its user went to the bathroom or because it lost power) and re-connect to the same AP a minute later, the device would be considered present at that AP before, during, and after the brief lapse.

Importantly, we cannot confirm if two individuals were within a reasonable range of each other for COVID-19 transmission; most AP-based interactions imply that the devices are within 10 meters of one another (but in the extreme cases, as much as 30 meters apart), which can span a wall or a floor. However, we anticipate that interactions at a distance that could lead to COVID-19 transmission are present in our pairwise data. Additionally, social networks, regardless of transmission mitigation strategies (*e.g.*, mask-wearing), can be inferred from these data.

The data was provided in the format of a device identifier (subsequently cross-referenced with metadata, including COVID-19 testing results of the user), the date, the AP (or set of APs), the building location (*e.g.*, “Hall B” or “academic building”), the length of time the user connected to the AP, and the (possibly empty) list of other devices that were simultaneously connected to the same AP, each with the duration of overlap.

To analyze pairwise interactions, the data were de-duplicated and cleaned, as instructed by Degree Analytics. We limited users within the dataset to only wifi-authenticated students, thus removing guests, faculty/staff, and stagnant devices on the network. Next, we removed student identifiers that were only ever present 3 or fewer times on campus over the entire year, as we assessed that these individuals were remote students who infrequently commuted to and participated in the on-campus CMU community or testing program. This cleaning removed 45,354 identifiers from the original 53,100 identifiers, producing a finalized dataset of 7,746 students.

To assess for differences in presence across buildings and semesters, we quantified the daily number of AP connections and the median duration of AP connections per building and per day of week, for each semester (Appendix Table 11; Appendix Figure 6). Due to the differences in connectivity patterns across semesters, we conducted all analyses on a per-semester and per-day basis.

##### 2. Interaction metric comparisons

We examined the daily interaction patterns for users (*i.e.*, nodes in our network), dividing them into students who tested positive at some point over the semester (“positives”) and students who did not test positive over the entire semester (“negatives”). Within our network, edges represent pairwise interactions. We quantified an individual student’s daily interactions via: (1) number of unique contacts, (2) average exposure time, and (3) number of days on campus (Appendix Table 11; Figure 3A, Appendix Figures 5AB and 7B). We assessed for significant differences between positive and negative individuals for both fall and spring semesters.

We assessed for differences in on-campus presence between positive and negative users. We calculated the proportion of users (positive or negative) on campus each day, calculated the Pearson correlation between the positive and negative proportions, and assessed for differences in these proportions via the Mann Whitney U test (Appendix Figure 5CD).

To investigate wifi-derived contacts during the isolation period, we calculated the median number of unique contacts across all positive individuals for each day in the 10 days prior to *vs.* after an individual’s positive test (Appendix Figure 7A). The average of the daily medians for each 10 day period was then used to calculate the percent change between the two periods.

Next, we redefined positive users as those within the 10-day window before a positive test (alternately referred to as “positives” or as “pre-positives”), and negative users as those who were not currently within a 10-day window prior to testing positive (regardless of testing status before or after the 10-day window). We examined pairwise interaction patterns for pairs of users with: (1) two positive users who were reported as contact tracing pairs (*i.e.*, CC positive pairs), (2) two positive users who were not listed as pairs in contact tracing (*i.e.*, non-CC positive pairs) (3) a positive and a negative user (mixed pairs), and (4) two negative users (negative pairs). We quantified daily pairwise interactions via the median and the total daily interaction duration and assessed for differences for both the Fall and Spring semesters (Appendix Table 11; Appendix Figures 7CD).

For all comparisons, we used the Mann Whitney U test to produce uncorrected p-values.

##### 3. Attribute assortativity

The attribute assortativity (AA) coefficient is a metric that quantifies the tendency for users to interact within *vs.* across particular cohorts ^2^. To compute this metric, we compared interactions either between positive and negative individuals, or pre-positive and negative individuals (as defined in the previous section). The AA coefficient is bounded between -1 and 1, where -1 represents a network where individuals only interact across-group, 0 represents a perfectly-mixed network, and 1 represents a network where individuals only interact within-group (Figure 3B).

We calculated the AA coefficient for sub-groups of individuals per day (using the NetworkX Python package) ^3^, and generated 95% confidence intervals (CI) for AA by randomly permuting attribute labels (40 times, with the lowest and second highest AA defining the bounds of the 95% CI) across individuals within each day’s network (Figure 3C, Appendix Figure 8A-C). We expect CIs to overlap with the per-day AAs for approximately 95% of the days if positive individuals were equally likely to interact with other positive individuals as they were with negative individuals. We ran this procedure three times for each semester (six times in total):

1. Defining positives as individuals who test positive for COVID-19 at some point during the semester (Appendix Figure 8AB)
2. Defining positives as all individuals who are within 10 days of testing positive (*i.e.*, “pre-positives”; Figure 3C, Appendix Figure 8C)
3. Defining pre-positives as individuals who are within 10 days of testing positive and are not listed in contact tracing data as having a pairwise association with another positive individual (*i.e.*, “non-CC pre-positives”; Figure 9AB)

For follow-up analysis for AA trends, we used the definition of pre-positives defined above as (2).

We assessed the relationship between pre-positive *v.* negative attribute assortativity and case counts, with the hypothesis that social network structure may be predictive of future case counts. We plotted both the raw data and the smoothed data (via the Savitzky-Golay filter; window length = 17, polynomial order = 4) ^4^. We determined the lag time, in days, that produces the maximal Pearson correlation between per-day AA and daily case counts, for both the raw and smoothed data and for both the fall and spring semesters (Figure 3DE, Appendix Figure 8D-F).

##### 4. Wifi analyses specific to the B.1.429.1 lineage

We compared the total exposure time and the median interaction duration (Appendix Table 11; Appendix Figure 12AB) for pairs of B.1.429.1-positive cases, pairs of non-B.1.429.1-positive cases, and pairs of negative individuals.

We constructed a subgraph of the network where nodes represent individuals in the pre-positive 10-day window, and edges connect two pre-positive individuals with documented proximity. We defined two individuals as within the same social network if they could be connected by a path, and defined two individuals as not within the same social network if they were disjoint and no path could connect them.

We examined whether the distribution of the viral genome SNV distances for B.1.429.1 pairs within the same social network differed from the distribution of SNV distances for B.1.429.1 pairs in different social networks (Figure 6D). We used the Mann Whitney U test.

To test the hypothesis that B.1.429.1 individuals clustered together in the network, we quantified the shortest path^5^ between individuals in pairs of two conditions, where: 1) both individuals had B.1.429.1-lineage virus, or 2) one individual had B.1.429.1 and one had a non-B.1.429.1 viral genome. We used the Mann Whitney U test.

#### Wastewater surveillance and sequencing

##### 1. **Installation and operation of wastewater samplers**

Five on-campus sewage sites were monitored in Fall 2020, with six additional sites added in Spring 2021. The effluent collected at all sites originated from only on-campus sources. Of the five original sites, three were downstream of specific non-isolation dormitories, a fourth contained the wastewater from a dorm housing COVID-positive individuals in isolation, and a fifth was located at the confluence of two dormitories and the wastestream that began at the isolation dorm. The six sites added in Spring included four dormitory sampling locations (two of which were downstream of academic buildings), and two sites near academic buildings but upstream of residential buildings (Appendix Figure 3D).

Automatic wastewater samplers were custom built based off of Reeves et al.^6^ Each sample was a composite from a 24-hour time period. Samples were collected twice weekly in the fall, and three times weekly in the spring across all available sites (Appendix Figure 3C). Automatic samplers pumped water continuously at a rate of approximately 4 gallons/24 hours or 10.5 mL/min. Samplers consisted of stainless-steel strainers deployed into the sanitary sewer. Silicone flexible tubing connected the strainer to a five-gallon high-density polyethylene (HDPE) jerrycan; water was displaced via a peristaltic pump run by a portable battery. After the 24-hour sample collection period, the jerrycans were gently mixed and three 40 mL samples were collected for processing at each site. Samples were collected with sterile serological pipette tips in an autopipetter and transferred to sterile 45 mL conical tubes. Samples were stored on ice or placed in a 4C refrigerator overnight for a maximum of 18 hours prior to processing.

After each sampling event, the strainer and silicone tubing were cleaned by pumping a 10% bleach solution through the system. These components were dried between each sampling event by storing them in the permanent wooden sampling boxes anchored above each open manhole. These boxes were locked to restrict access. Each jerrycan was first sanitized with 10% bleach thrice, and following bleach treatment was subsequently cleaned thrice with dish soap and water and allowed to dry. Upon deployment, wastewater was pumped through the system and sent back into the sewer prior to sample collection to rinse the strainer and tubing.

##### 2. Quantification of viral concentration

In the Fall semester, all samples were sent to GT Molecular for viral titer quantification. At the beginning of the Spring semester, samples were processed both at GT Molecular and on campus, with CMU validating its data against results received from GT Molecular. From Feb. 15, 2021 onwards, samples for all sites were processed on campus. During Spring 2021, technical duplicates were processed for three or four sites (of the eleven total) during each sampling event, to serve as an internal validation of viral concentration.

CMU followed a standard procedure to calculate viral titer^7^. Each sample volume was adjusted the next day to 40 mL and spiked with 13.6 μL Bovilis Coronavirus Calf Vaccine (BCoV) (Merck Animal Health Cat. No.16445), reconstituted in 2 mL 0.01% Tween20 in 1x PBS. BCoV was added to determine viral recovery yield of the concentration step during subsequent RT-qPCR. The samples were inverted three times to mix. 400 μL of 5% Tween 20 was added to each tube and samples were inverted three times to mix. The samples were centrifuged at 7000 x g at 4 °C for 10 minutes. The supernatant was carefully transferred to a fresh 50 mL conical tube without disturbing the pellet. The supernatant was concentrated with the InnovaPrep concentrating pipette. Elution was done in 0.075% Tween 20/25 mM Tris. The concentrated samples were stored on ice until all samples were processed.

Virus RNA was extracted with the QIAamp Viral RNA Mini Kit (Qiagen) with minor changes to the manufacturer’s protocol. The tubes were incubated for 15 minutes at room temperature upon pipetting 140 μL of the concentrated wastewater tubes with 560 μL of AVL buffer containing carrier RNA. During the AW2 wash, the spin column was centrifuged three times, first for three minutes at full speed, and the next two spins for one minute each at full speed and with open lids. For each spin the old collection tube was replaced with a new collection tube. After the third spin, the spin columns were placed in microfuge tubes and incubated with open lids for 15 minutes at room temperature to allow any remaining ethanol to evaporate. For the elution of RNA, 60 μL of nuclease-free water was added to the membrane, incubated at room temperature for 1.5 minutes, and spun at 6000 g for 2 minutes. The extracted RNA was stored on ice briefly until it was used for digital PCR.

The digital PCRs were performed as twoplex assays with TaqMan hydrolysis probes, on the QIAcutyOne 2plex (Qiagen) platform. QIAcuty One-Step Viral RT-PCR Kit (Qiagen) was used to quantify the viral load. The duplex reactions were 40 μL and contained 24 μL of the purified RNA, 1X One-Step Viral RT-PCR Master Mix, 1X Multiplex Reverse Transcription Mix, SARS-CoV-2 and BCoV forward and reverse primers at 0.4 μM and probes at 0.2 μM. SARS-CoV-2 nCOV_N1 RUO primers and probe and BCoV primers (NOC43-1 and NOC43-2) were purchased from IDT. BCoV probe (NOC43-p) labeled with HEX fluorophore and OQA quencher was purchased from Sigma-Aldrich.

The QIAcuity was programmed to 50 °C for 40 minutes for reverse transcription, 95 °C for 2 minutes for initial heat inactivation, and 40 cycles of denaturation at 95 °C for 5 seconds and annealing/extension at 55 °C for 30 seconds.

##### 3. Flow-mediated mass balance correction

Proximity to dormitories was prioritized for placement of wastewater samplers. In a few cases, there were other dormitories or academic buildings that contributed sewage upstream of specific dormitories (Appendix Figure 3D). In these cases, additional samplers were placed upstream, and the background SARS-CoV-2 concentration for samples from upstream sites was subtracted from concentrations obtained from downstream sites, using a flow-mediated mass balance based on building-level potable water consumption to account for dilution.

##### 4. Comparison of viral titers and weekly case counts

We conducted two analyses to assess the relationship between viral titer and weekly case count. We first compared each individual wastewater sample against its corresponding weekly case count, across all collection sites with the exception of Site 5 which collected effluent from isolated positive cases. We calculated the Spearman correlation coefficient and its associated p-value.

Second, we calculated each hall’s average wastewater viral titer (*i.e.*, the average of available samples from Sunday through Saturday) and each hall’s total case count for each week. If a hall had no wastewater samples collected in a given week, it was removed. We then proceeded in a hall-wise fashion to determine the sign of the slope of the viral titer and case count (*i.e.*, to assess whether titer and case count rose or fell together from one week to the next). We created a contingency table of the sign of the slope of viral titer *vs.* the sign of the slope of case counts, and evaluated its significance via Fisher’s exact test.

##### 5. Viral sequencing analysis

During the 6 epi-weeks from Sunday, Feb. 9 through Mar. 20, 2021, viral RNA in aliquots of excess extracted wastewater from 42 samples was sequenced via the same ARTIC v3 procedure described below for clinical samples. Samples were sequenced in three batches. The final batch of nine samples was sequenced with technical replicates obtained by splitting the cDNA produced from the RNA template prior to library construction.

We inspected sequence data for the presence of regional blindspots in the genome distinct to wastewater as a sample type. To assess whether specific regions of the genome were more susceptible to degradation in wastewater rather than in clinical samples, we normalized read depth per base for each sample and plotted the distribution of depth across all wastewater samples, alongside a corresponding plot of depth from all clinical samples (Appendix Figure 10BC). We compared the median normalized depth per amplicon between wastewater and clinical samples by calculating the Pearson correlation (Appendix Figure 10E). Next, we compared amplicon read depth and Shannon entropy within the primer regions of wastewater sequences by calculating the Pearson correlation (Appendix Figure 10D). We used entropy data from a CDC-curated Nextstrain analysis focused on data from Colorado as of August 2021^8^.

##### 6. Development of quality controls for identifying SNVs in wastewater

We evaluated three quality control filters to remove spurious SNVs identified in wastewater: minimum allele frequency (AF), minimum read depth (DP), and presence in each of two replicates from the same cDNA source (Reps) (Figure 5E). For both AF and DP, we independently toggled their threshold from absolute minimum (AF=0, DP=0) to absolute maximum (AF=1, DP=29903). Since replicates were only available for nine of the forty-two wastewater samples, analyses were limited to those nine samples for consistent comparisons across the three quality control filters.

We investigated which quality control mechanisms identified the greatest number of wastewater SNVs present in any Colorado clinical sample. For AF and DP thresholds, sensitivity and specificity were defined as follows:

WW = set of SNVs in wastewater samples

CO = set of SNVs in Colorado clinical samples

$$Sensitivity of AF threshold x = \frac{cardinality( WW \cap CO \cap\{SNVs with AF \geq x\} )}{cardinality(WW \cap CO)}$$

$$Sensitivity of DP threshold x = \frac{cardinality( WW \cap CO \cap\{SNVs with DP \geq x\} )}{cardinality(WW \cap CO)}$$

$$Specificity of AF threshold x = \frac{cardinality( (WW - (WW\cap CO)) \cap\{SNVs with AF < x\} )}{cardinality( (WW - (WW\cap CO)) )}$$

$$Specificity of DP threshold x = \frac{cardinality( (WW - (WW\cap CO)) \cap\{SNVs with DP < x\} )}{cardinality( (WW - (WW\cap CO)) )}$$

For the Reps filter, sensitivity and specificity were calculated for each of the nine samples, rather than for the entire subset of samples. Sensitivity and specificity were defined as follows:

WW_X,union_ = set of SNVs found in either replicate of sample X

WW_X,intersection_ = set of SNVs found in both replicates of sample X

$$Sensitivity for sample X = \frac{cardinality( {WW}_{X,intersection} \cap CO )}{cardinality( {WW}_{X,union} \cap CO )}$$

$$Specificity for sample X = \frac{cardinality({WW}_{X,union} - {WW}_{X,intersection} - (({WW}_{X,union} - {WW}_{X,intersection}) \cap CO) )}{cardinality({WW}_{X,union} - ({WW}_{X,union} \cap CO) )}$$

##### 7. Analysis of expected number of unique SNVs contributed by additional samples

We predicted the number of unique SNVs present within a given number of CMU wastewater or clinical samples (Appendix Figure 11BC). For clinical samples, we bootstrapped 100 times over each possible subset size (*i.e.*, from 1 sample to all samples) to curate a set of clinical samples. We calculated the total number of unique consensus-level SNVs across each set of *n* samples, then found the average number of unique consensus-level SNVs that we could expect *n* samples to contribute.

For wastewater samples, we also bootstrapped 100 times over each possible subset size (*i.e.*, from 1 sample to all 42 samples) to curate a unique set of wastewater samples. We then calculated 1) the average number of SNVs across sets of *n* samples, and 2) the average number of SNVs of AF greater than or equal to 25% across sets of *n* samples. We repeated this process with the wastewater samples with technical replicates, again bootstrapping 100 times over each possible subset size (i.e. from 1 sample to all 9 samples) to calculate the average number of replicate-confirmed SNVs that we could expect from a set of *n* samples.

Finally, for all of the above metrics, we calculated the smoothed first derivative (*i.e.*, change in SNV count as a function of the number of samples) using a window size of 5 (Appendix Figure 11DE).

##### 8. Lineage identification in wastewater

To implement lineage detection across our sequenced wastewater samples, we called SNVs using LoFreq with default parameters. We estimated the relative abundance of constituent lineages using Freyja v1.3.4 ^9^. We limited analyses to samples with at least 30% genome coverage, and lineages that were detected with 95% confidence (per Freyja’s built-in bootstrapping capabilities, 5000 replicates) with at least 3% abundance. Lineages were assigned using freyja with a global UShER tree downloaded on March 14, 2022.

#### Clinical genomic analysis

##### 1. Sequencing and viral genome assembly

Members of the CMU community underwent diagnostic testing for SARS-CoV-2 infection using either saliva or nasal specimens, collected in response to random surveillance testing and reflexive testing. Residual material was only available for saliva specimens, accounting for a fraction of known cases during the 2020–2021 school year (Figure 2A, Appendix Figure 2B).

Saliva samples were collected from members of the campus community and sent to Warrior Diagnostics, Inc., for clinical diagnostic RT-qPCR testing. Excess material from specimens found to be positive for SARS-CoV-2 was inactivated and sent to the Broad Institute of MIT and Harvard for viral genomic sequencing. In initial sequencing rounds, samples were treated with 5 uL each of proteinase K; we determined that excluding this step did not negatively impact sequencing quality, and did not include it in later sequencing. Total RNA was extracted from the samples using the Thermo Fisher MagMAX Viral RNA Isolation kit. Concentration of viral RNA was determined through RT-qPCR with primers and probes targeting the SARS-CoV-2 N gene. Illumina sequencing libraries were prepared from tiled amplicons amplified using the ARTIC v3 primer set ^10–12^. The libraries were pooled and sequenced on Illumina NovaSeq and NextSeq instruments.

Using viral-ngs v2.1.28.0 pipelines running on Terra (https://app.terra.bio), reads from sequenced pools were demultiplexed, filtered to remove adapter sequences and contaminant sequences, depleted of reads mapping to the human genome, and assembled by alignment to the reference sequence NC_045512.2. All software used is publicly available on Dockstore (https://dockstore.org/organizations/BroadInstitute/collections/pgs) and GitHub (https://github.com/broadinstitute/viral-pipelines).

A total of 184 samples (of 278 received) from clinical diagnostic tests were successfully sequenced to yield viral genomes with median assembly length of 29827 bases (Appendix Figure 2C). Assembled viral genomes with at least 24000 unambiguous bases and successful annotation were deposited in NCBI GenBank as part of Bioprojects PRJNA715749 or PRJNA622837; accessions are listed in Appendix Table 5.

##### 2. Lineage assignment

Lineage designations were assigned to viral genomes using Pango v4.0.6 with pango-data v1.9 ^13,14^.

##### 3. Phylogenetic analysis

CMU genomes were aligned to the reference sequence NC_045512.2 using MAFFT v7.471 using the "--addfragments" and "--keeplength" arguments to ensure the resulting alignment would be in the same coordinate space as the reference sequence. It is of note that these parameters, widely used in the alignment of SARS-CoV-2 genomes, can produce alignments which omit insertions. This limitation was deemed acceptable for this study due to the rarity of known insertions in the SARS-CoV-2 genome at the time of sampling.

Using the Nexstrain augur pipeline, a maximum likelihood (ML) tree was created via IQ-Tree using a GTR mutation model, as well as a time-resolved tree via TreeTime, both rooted to the ancestral reference genome, NC_045512.2 ^15–17^. A filter was specified for TreeTime to exclude outlier sequences >4 interquartile distances from the root-to-tip *vs.* time (*i.e.*, molecular clock mean mutation rate) regression. Internal tree nodes were assigned dates based on their marginally most likely dates. CMU samples were placed in the context of viral genomes fromstate, national, and global datasets, weighted toward those collected in the US Mountain West states and those of short genetic distance from viral genomes from CMU samples. The contextual genomes were obtained as part of the open dataset of pre-aligned sequences curated by Nextstrain^18^. Default augur quality thresholds were applied to input sequences, which include only those sequences >27 kb in length.

To identify introductions to the campus community, ancestral state reconstruction was performed using TreeTime to produce a binary value indicating whether each viral genome or internal ancestral tree node was university-associated or not-associated. Following assignment of state for this value to all tree nodes, a state change from not-associated to university-associated descendant cases was considered a putative introduction. Sub-trees for each introduction event were extracted and plotted using the *baltic* Python library^19^, for those where the confidence of the inferred state was >0.8.

The number of intermediate hosts in a cluster noted to span semesters was estimated using TransPhylo, and generation time distribution parameters reported previously (shape=3.63, scale=1.408) ^20^.

##### 4. Overdispersion analyses

The distribution of offspring per cluster was calculated as the total number of individuals in the cluster minus one (being the introduction case itself). A negative binomial distribution was fit to the data using ‘*fitdistplus*’ in R 4.1.2.

The university conducted contact tracing for positive cases, who reported individuals that they had been in contact with for more than 15 minutes at less than 6 feet within the prior 48 hours of the earliest of their positive test date or their symptom onset date. From these investigations, the unique number of contacts reported per positive case were counted and a negative binomial distribution was fit to the data.

Using the previously described pairwise wifi proximity dataset, the interactions of positive users across the entire year (with an individual of any other test status) were extracted from the data. The interactions were then filtered to only include those that occurred during the 48 hours prior to the earlier date of either positive test or symptom onset. The total number of unique interactions in this period was calculated for each positive user, and a negative binomial distribution fit to the resulting data.

##### 5. Transmission network reconstruction

To reconstruct transmission networks, we first excluded sequences with more than 7% ambiguous bases (>2093 N). The remaining sequences were aligned to the reference genome NC_045512.2 using MAFFT v7.471 with the parameters "--addfragments" and "--keeplength". Following alignment, positions identified as prone to sequencing error or homoplasy were masked with ambiguous bases using the positions previously documented^21^. The 5' and 3' untranslated regions of the genome were also masked over the reference sequence positions 1-265 and 29558-29903. Following alignment and masking, any sequences with >10% ambiguity or >7% gaps across the genome were excluded.

Three forms of contact data were included in the transmission network model: 1) contact tracing data from university tracing efforts; and contacts assumed from shared proximity to Wi-Fi access points for individuals in contact within 2) 2 days or 3) 10 days of both case dates. We also developed models with 1) solely genomic and 2) solely contact tracing data for comparison. Case dates were the earlier of the date of symptom onset, when known, and the date of diagnostic test.

The probability of direct transmission between cases bearing B.1.429.1-lineage virus was estimated from case dates, viral genomes, and contact data using outbreaker2 with parameters previously described^22^ and a single chain of 40,000 iterations (of which the first 10% were discarded). Visualizations include all transmission events with a probability greater than or equal to 25%. For the three networks generated with a combination of genomic and contact data, we compared clusters of 2 or more individuals *via* the Jaccard distance.

#### Functional characterization of non-synonymous changes at position 677 of the Spike glycoprotein

##### 1. Lentivirus production

24 hrs prior to transfection, 6 × 10^5^ HEK-293T cells were plated per well in 6-well plates. All transfections used 2.49 µg plasmid DNA with 6.25 µL TransIT LT1 transfection reagent (Mirus, Madison, WI) in 250 µL Opti-MEM (Gibco). Single-cycle HIV-1 vectors pseudotyped with the indicated SARS-CoV-2 Spike constructs were produced by transfection of HIV-1 pNL4-3ΔenvΔvpr luciferase reporter plasmid (pNL4-3.Luc.R-E-; NIH AIDS Reagent Program, Division of AIDS, NIAID, NIH: from Dr. Nathaniel Landau; ARP Cat #3418) with the indicated Spike expression plasmid, at a ratio of 4:1.

##### 2. Lentivirus infectivity assays

16 hours prior to transduction, HEK-293T cells stably expressing ACE2/TMPRRS2 as previously described ^23^ were plated at 3 x 10^4^ per well. Cells were incubated in virus-containing media for 16 hours at 37 °C after which fresh media was added to cells. 48 hours after transduction, cells were assessed for luciferase activity using the Promega Steady-Glo system (Promega Madison, WI).

##### 3. Western blot analysis

Tissue culture media and cell lysate were collected 60 hours after transfection to produce lentivectors. Supernatant containing Spike pseudotyped particles was layered on a 20% sucrose cushion in PBS and spun at 110,000 x g at 4 °C for 2 hrs. The pellet was washed once with ice-cold PBS and resuspended in 15 uL of 2x SDS gel loading buffer. After removal of supernatant, transfected cells were lysed in 300 uL 2x SDS-PAGE loading buffer. Protein preps were boiled for 5 minutes and then separated by SDS-PAGE on a 10-20% Tris-Gycine gel (BioRad). Proteins were electro-transferred from gels to nitrocellulose membranes, which were blocked for an hour with Licor Blocking Buffer and detected with the indicated antibodies.

##### 4. Cell fusion assay

ACE2/TMPRSS2 expressing cells were prepared by transfecting 293T cells with pcDNA3.1- ACE2 and pcDNA3.1-TMPRSS2 along with pscALPs LgBit. Spike expressing cells were prepared by transfecting 293T cells with pcDNA3.1- constructs expressing the specified codon optimized SARS-CoV-2 Spike proteins, in addition to the pscALPs HiBit-FLuc fusion expression vector. 24 hours after transfection, ACE2/TMPRSS2 and Spike expressing cells were lifted from plates with TrypLE and plated together in a 1:1 ratio for a total of 40,000 cells in 96-well white-walled tissue culture plates. Promega Endurazine substrate was added to cells according to the manufacturer’s protocol 1 hour after plating and fusion was analyzed 4 hours later. Fusion signal of HiBit-LgBit interaction was normalized to Fluc signal to control for transfection efficiency. Background fusion was determined by using 293T cells transfected with pscALPs HiBit-Fluc alone with control pcDNA3.1- plasmid without Spike. Plasmids are available from *addgene* (https://www.addgene.org/Jeremy_Luban/).

8.	auspice <https://nextstrain.org/groups/spheres/ncov/colorado>.

9.	Freyja: Depth-weighted De-Mixing (Github).

10.	Quick, J., Grubaugh, N.D., Pullan, S.T., Claro, I.M., Smith, A.D., Gangavarapu, K., Oliveira, G., Robles-Sikisaka, R., Rogers, T.F., Beutler, N.A., et al. (2017). Multiplex PCR method for MinION and Illumina sequencing of Zika and other virus genomes directly from clinical samples. Nat. Protoc. *12*, 1261–1276.

11.	Betteridge, E., Park, N., James, K., Durham, J., and Quick, J. COVID-19 ARTIC v3 Illumina library construction and sequencing protocol - short amplicons (275bp) v1. protocols.io.

12.	Tyson, J.R., James, P., Stoddart, D., Sparks, N., Wickenhagen, A., Hall, G., Choi, J.H., Lapointe, H., Kamelian, K., Smith, A.D., et al. (2020). Improvements to the ARTIC multiplex PCR method for SARS-CoV-2 genome sequencing using nanopore. bioRxivorg.

13.	O’Toole, Á., Scher, E., Underwood, A., Jackson, B., Hill, V., McCrone, J.T., Colquhoun, R., Ruis, C., Abu-Dahab, K., Taylor, B., et al. (2021). Assignment of epidemiological lineages in an emerging pandemic using the pangolin tool. Virus Evol *7*, veab064.

14.	Rambaut, A., Holmes, E.C., O’Toole, Á., Hill, V., McCrone, J.T., Ruis, C., du Plessis, L., and Pybus, O.G. (2020). A dynamic nomenclature proposal for SARS-CoV-2 lineages to assist genomic epidemiology. Nat Microbiol *5*, 1403–1407.

15.	Hadfield, J., Megill, C., Bell, S.M., Huddleston, J., Potter, B., Callender, C., Sagulenko, P., Bedford, T., and Neher, R.A. (2018). Nextstrain: real-time tracking of pathogen evolution. Bioinformatics *34*, 4121–4123.

16.	Sagulenko, P., Puller, V., and Neher, R.A. (2018). TreeTime: Maximum-likelihood phylodynamic analysis. Virus Evol *4*, vex042.

17.	Minh, B.Q., Schmidt, H.A., Chernomor, O., Schrempf, D., Woodhams, M.D., von Haeseler, A., and Lanfear, R. (2020). IQ-TREE 2: New Models and Efficient Methods for Phylogenetic Inference in the Genomic Era. Mol. Biol. Evol. *37*, 1530–1534.

18.	Overview of remote nCoV files (intermediate build assets) — SARS-CoV-2 Workflow documentation <https://docs.nextstrain.org/projects/ncov/en/latest/reference/remote_inputs.html?highlight=open>.

19.	Dudas, G. baltic: baltic - backronymed adaptable lightweight tree import code for molecular phylogeny manipulation, analysis and visualisation. Development is back on the evogytis/baltic branch (i.e. here) (Github).

20.	Zhang, J., Litvinova, M., Wang, W., Wang, Y., Deng, X., Chen, X., Li, M., Zheng, W., Yi, L., Chen, X., et al. (2020). Evolving epidemiology and transmission dynamics of coronavirus disease 2019 outside Hubei province, China: a descriptive and modelling study. Lancet Infect. Dis. *20*, 793–802.

21.	ProblematicSites_SARS-CoV2 (Github).

22.	Siddle, K.J., Krasilnikova, L.A., Moreno, G.K., Schaffner, S.F., Vostok, J., Fitzgerald, N.A., Lemieux, J.E., Barkas, N., Loreth, C., Specht, I., et al. (2022). Transmission from vaccinated individuals in a large SARS-CoV-2 Delta variant outbreak. Cell *185*, 485–492.e10.

23.	Yurkovetskiy, L., Wang, X., Pascal, K.E., Tomkins-Tinch, C., Nyalile, T.P., Wang, Y., Baum, A., Diehl, W.E., Dauphin, A., Carbone, C., et al. (2020). Structural and Functional Analysis of the D614G SARS-CoV-2 Spike Protein Variant. Cell *183*, 739–751.e8.
